# Systematic comparison of phenome-wide admixture mapping and genome-wide association in a diverse biobank

**DOI:** 10.1101/2024.11.18.24317494

**Authors:** Sinead Cullina, Ruhollah Shemirani, Samira Asgari, Eimear E. Kenny

**Affiliations:** Institute for Genomic Health, Icahn School of Medicine at Mount Sinai, New York, NY; Department of Genetics and Genomic Sciences, Icahn School of Medicine at Mount Sinai, New York, NY, USA; Division of Genomic Medicine, Department of Medicine, Icahn School of Medicine at Mount Sinai, New York, NY, USA; Division of General Internal Medicine, Department of Medicine, Icahn School of Medicine At Mount Sinai, New York, NY, USA

## Abstract

Biobank-scale association studies that include Hispanic/Latino(a) (HL) and African American (AA) populations remain underrepresented, limiting the discovery of disease associated genetic factors in these groups. We present here a systematic comparison of phenome-wide admixture mapping (AM) and genome-wide association (GWAS) using data from the diverse Bio*Me* biobank in New York City. Our analysis highlights 77 genome-wide significant AM signals, 48 of which were not detected by GWAS, emphasizing the complementary nature of these two approaches. AM-tagged variants show significantly higher minor allele frequency and population differentiation (Fst) while GWAS demonstrated higher odds ratios, underscoring the distinct genetic architecture identified by each method. This study offers a comprehensive phenome-wide AM resource, demonstrating its utility in uncovering novel genetic associations in underrepresented populations, particularly for variants missed by traditional GWAS approaches.

## Introduction

The primary motivation of large genetic studies is to characterize the contribution of genetic variation to complex human phenotypes. It is well established that progress in genomic research has been unequal, with historical biases toward populations of European ancestry ^1–4^. Although genetic differences between populations account for a small fraction of overall genetic diversity, there are often differences in the prevalence of health outcomes based on ancestry background as a result of both genetic and environmental factors ^5^. Even if the causal disease variants are not population differentiated, tag variants and thus genome-wide association studies (GWAS), often do not port well across populations due to differences in linkage disequilibrium (LD) patterns ^6–8^. For these reasons, the Eurocentric focus of genetic research has led to a disparity in our understanding of the genetic drivers of health and disease in diverse global populations ^3,4,9^. Numerous groups have responded to calls for diversification including All of US, PAGE and eMERGE consortium among others ^10,11^. As a result, genetic data from two major underrepresented US populations; African Americans (AA) and Hispanic Latinos (HL) are increasingly available to researchers. This presents an opportunity to use methods specifically designed to leverage genetic diversity, such as admixture mapping (AM), which can enhance the discovery of novel genomic insights. Evaluating such approaches is particularly critical given the rapidly growing representation of HL and AA populations in the US, allowing for more accurate and equitable health outcomes through genomic research ^12,13^.

AM is a genetic association method for mapping risk loci in recently admixed populations ^14^. HL and AA populations are considered recently admixed with ancestry contributions from two or more continents as a result of historical events including the transatlantic slave trade and colonization of the Americas ^15,16^. HL have varying genetic contributions from populations with African, European and Native American continental ancestry ^17–19^. AA have major genetic contributions from populations with African and European continental ancestry ^16^. In AM, cases are tested in a genome-wide scan for enrichment of local ancestry haplotypes from one of the ancestral populations compared to controls ^20,21^. The conceptualization of AM as a method dates back many decades but it wasn’t until the advent of ancestry informative markers (AIMs) that AM became functionally possible ^14,22,23^. The development of local ancestry calling algorithms such as RFMix and GNOMIX along with dense genotyping data offers greater resolution of ancestral haplotypes and more sophisticated implementations of AM ^24,25^. AM has been used considerably less than GWAS, however it has contributed to major discoveries including the 22q12 locus and end stage renal disease in AA ^26,27^, 8q24 locus and prostate cancer in AA ^28^ and 13q33.3 locus and Alzheimer’s disease in HL ^29^.

Both GWAS and AM are designed to detect different types of casual variation. AM is most effective when the causal allele or phenotype differs in frequency between ancestral populations ^14^. On the other hand, GWAS is generally more effective for detecting common variants with small to moderate effects, though it requires large sample sizes to do so ^7,30^. One key difference between the two methods lies in the number of tests performed. AM involves fewer effective tests because it assesses longer local ancestry blocks, which result from relatively recent admixture events. In recently admixed populations, these local ancestry tracts are longer and have undergone less recombination, compared to the shorter LD haplotypic blocks tested in GWAS ^31–33^. As a result, AM has a smaller multiple test burden and requires smaller sample sizes compared to GWAS ^33^. Although both methods have been used to complement each other in some studies ^31,34,35^, a comprehensive comparison of their discovery potential across a wide range of phenotypes is lacking.

In this study we compare AM and GWAS in the diverse Bio*Me* health system biobank using a phenome-wide association study (PheWAS) framework (**Figure 1**). The PheWAS framework was developed to facilitate phenome-wide association testing across hundreds of disease phenotypes available in health systems electronic health record (EHR) data. We develop a well-calibrated and optimized phenome-wide AM resource, and demonstrate its utility in uncovering novel genetic associations in admixed populations. We also perform fine-mapping of associated peaks, and compare tag-variants for GWAS and AM loci across various metrics, offering an evaluation of the types of results each method is effective to detect. As genetic research increasingly moves towards the inclusion of diverse, admixed populations in line with the changing global demographic landscape ^36^, it is vital that researchers are primed with the resources to understand the efficacy of different association testing methods.

**Figure 1:**
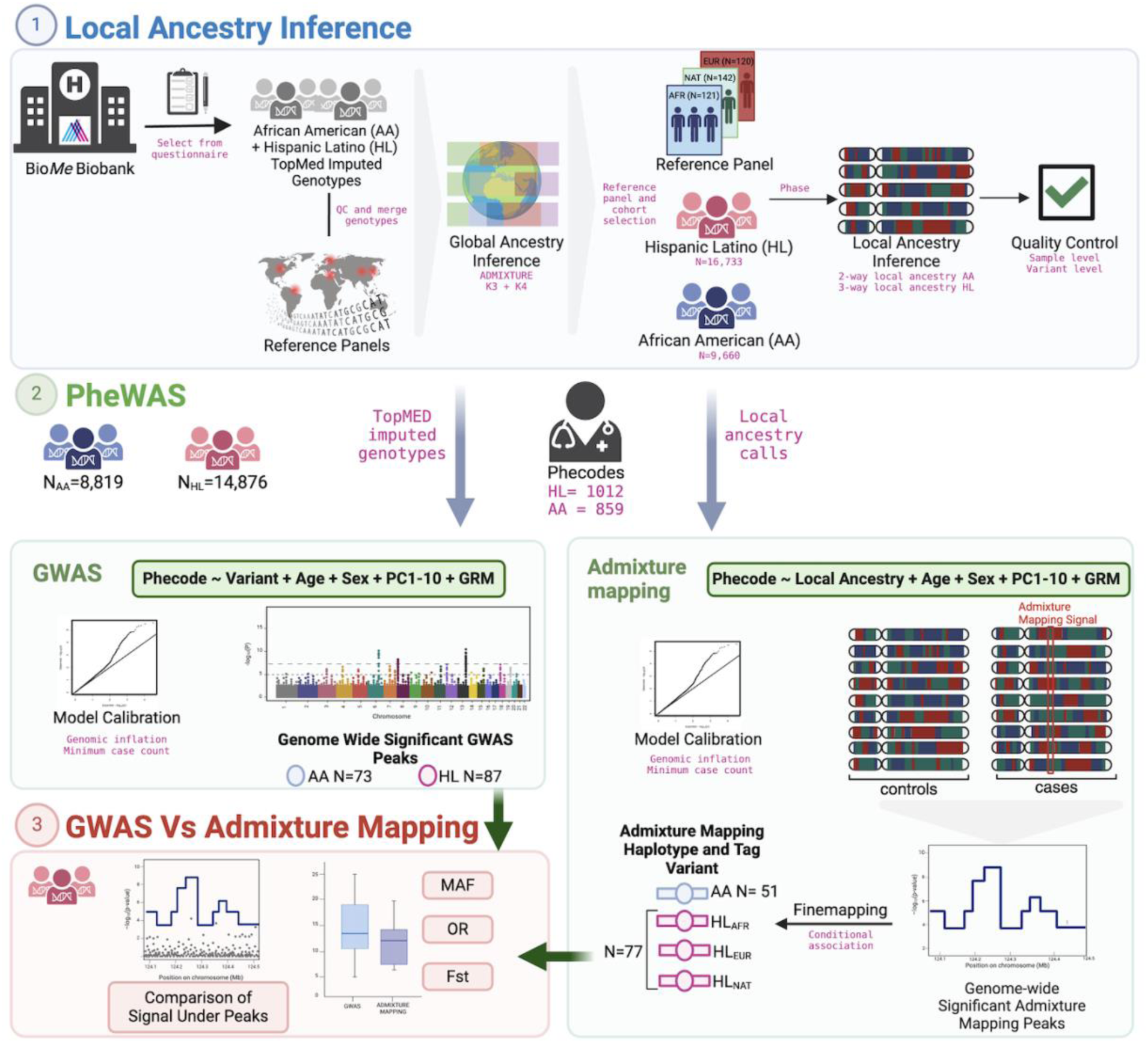
Study design for systematic comparison of admixture mapping and genome-wide association at a phenome-wide scale. Overview of workflow showing strategy for local ancestry inference, methods for genome-wide association and admixture mapping across the medical phenome, and metrics used for evaluation. HL, Hispanic Latino; AA, African American; GWAS, genome-wide association; PheWAS, phenome-wide association

## Results

### Overview of study design

First we describe a framework to do a direct comparison of GWAS and AM approaches across hundreds of phenotypes, a detailed overview is described in **Supplemental Figure 1**. Leveraging data from the Bio*Me* Biobank program at the Icahn School of Medicine at Mount Sinai, participants with genotype data passing quality control (QC) were selected for imputation using the TOPMed reference panel (see methods). So that we could perform both AM and GWAS analysis on these individuals, we selected participants with Hispanic/Latino (H/L) and African-American (AA) self-reported identity. Participants were surveyed about their heritage, Hispanic Latin American identity, and country of birth (see **Supplemental Figure 2**), with responses summarized in **Supplemental Table 1**. A summary of sample sizes and demographics are provided in **Table 1**.

**Table 1:** Demographic information for HL and AA cohorts used in association testing with “All BioMe” representing BioMe participants with completed questionnaire data.

|  | Hispanic Latino |  | African American |  | All BioMe |  |
| --- | --- | --- | --- | --- | --- | --- |
| **Sample Size** | 17,150 |  | 10,160 |  | 53,449 |  |
| **Age (Median years, IQR)** | 62 (26) |  | 62 (23) |  | 61 (27) |  |
| **Sex, Female (%)** | 10,773 (62.8%) |  | 6,218 (61.2%) |  | 30,853 (58.0%) |  |
| **Genotype Chip (N GSA, %)** | 10,600 (61.8%) |  | 7,082 (69.7%) |  | 31,140 (58.3%) |  |
| **BMI (Median, IQR) N** | 28.46 (7.71) | 16,478 | 29.06 (9.48) | 9,720 | 27.18 (7.90) | 50,812 |
| **Ever Smoked (%) N** | 34.9 | 9,531 | 40.2 | 5,138 | 35.8 | 30,636 |

### Robust pipeline for admixture mapping

Most local ancestry (LA) inference methods ^24,25,37^ are designed to work with a specific number of source reference populations. Therefore we first estimated individual-level genetic similarity with reference panels to filter for complex ancestry in some HL and AA individuals. The dataset was then combined with genome sequencing data for three reference panels. After filtering and merging, the combined AA, HL, and reference panels comprised N=31,771 individuals and n=186,697 variants (**Supplemental Figure 3A**). Using the ADMIXTURE software, AA individuals with greater than 5% genetic similarity with Native American reference panels at K=3 were removed (N=356; ∼3.6%). HL individuals with greater than 5% genetic similarity with East Asian reference panels at K=4 were removed (N=271; ∼1.6%), This resulted in sample sizes of N=16,733 HL and N=9,660 AA for LA inference. In HL, the median proportion of genetic similarity with European reference populations at an individual-level was (58.9%, 95% CI =58.6%-59.2%), African (19.8%, 95% CI = 19.5%-20.3%) and Native American (12.8%, 95% CI = 12.7%-12.9%). In AA, the mean proportion of genetic similarity with European reference populations at an individual-level (12.5%, 95% CI= 12.2%-12.7%) and African (86.6%, 95% CI=86.4%-86.9%; **Supplemental Figure 3B**). Finally, to select reference panels for LAI, we aimed for a comparable panel size for each source population to help the algorithm more accurately differentiate between genetic signatures of each source population. There were only 154 samples across all three reference panels with ≥ 90% enrichment for the ADMIXTURE component seen in the Native American reference populations at K=4. Therefore, we also selected individuals that were ≥90% enriched for the ADMIXTURE component seen in the European reference populations at K=4, but downsampled to only used a subset who were the 1KGP Utah residents CEPH with Northern and Western European ancestry (CEU) population (N=178) and likewise for the ≥ 90% African and 1KGP Yoruba in Ibadan, Nigeria (YRI) population (N=178).

To run the local ancestry inference, we extracted the HL and AA and reference samples from the merged Bio*Me* + reference panel dataset (N = 26,903, n=479,586), and further removed 1st and 2nd degree relateds followed by phasing with Eagle v2.4. Final Sample sizes were AA (N=9022) and HL (N=15,149), YRI (N=121), CEU (N=120), Native American (N=142) reference panels and population descriptors are detailed in **Supplemental Tables 2 and 3**. GNOMIX was then used to call local ancestry with different window sizes for specific chromosomes, achieving high model accuracy. Quality control included comparing local ancestry proportions with global ancestry estimates and removing outlier samples and regions with enriched ancestry components (+/− 3SD from median ancestry proportion) or centromeric regions. Three-way LA was inferred in the HL group using NAT (Native American), EUR (CEU), and AFR (YRI) reference panels. Two-way local admixture was inferred in AA using EUR and AFR reference panels. We observed a high correlation of 99-100% between global ancestry proportions and the summed length of LA haplotypes for a given ancestry as a proportion of the genome (see **Supplemental Figure 4**).

### Admixture Mapping PheWAS

HL and AA were tested separately in a phenome-wide association study (PheWAS) framework. International Classification of Disease (ICD) codes (ICD-9 and ICD-10 from 2005-2023) were extracted from the EHR of Bio*Me* participants. ICD codes were mapped to phecodes (V1.2) which are used to categorize medical conditions and phenotypes. Cases were defined as having at least two instances of an ICD code for a given phecode; controls had none. Participants with one instance were considered “NA”. Permutations were performed to assess the robustness of the test statistic at varying case counts (see methods, **Supplemental Figures 5-8**), and demonstrated that the stability of test statistics was similar across all case counts tested. Therefore the lowest N=30 case count tested was set as the minimum case count for downstream AM. This resulted in 1012 phecodes (81 female specific and 17 male specific) tested in HL (N=14,876) and 859 (68 female specific and 10 male specific) phecodes tested in AA (N=8,819).

For sex-specific phecodes association testing was performed in the appropriate single sex cohort (N_Male_HL_=5,641, N_Female_HL_=9,235, N_Male-AA_ =3,491, N_Female_AA_=5,328). AM was performed using SAIGE which employs a generalized linear mixed model (GLMM) along with a genetic relatedness matrix (GRM) to account for test statistic inflation and cryptic relatedness. A pruned set of genotyped SNPs were used to construct a GRM in HL and AA cohorts separately (see methods). LA calls passing quality control were converted into VCF format. For AM in the HL cohort three separate VCFs were used, each with calls for an individual ancestry component e.g. for the AFR VCF all calls were recorded as either 0/0=no AFR ancestry, 0/1 AFR heterozygous, 1/1=AFR homozygous. Because the AA cohort has two-way admixture, only one association test is necessary for AM. The local ancestry calls in AA VCF files represent EUR ancestry status at each site, for example “1/1” indicates homozygous for EUR ancestry. The frequency distribution of local ancestry calls used in AA and HL PheWAS are shown in **Supplemental Figure 9**. A genome-wide significance threshold was defined using the STEAM software, and was estimated as *p<*1.60×10^−05^ in AA and *p<*4.88×10^−06^ in HL similar to previous estimates by other groups ^33^. We define a Bonferroni study-wide significance threshold of *p<*1.86×10^−08^ in AA and *p<*4.82×10^−09^ in HL by dividing the genome-wide significance threshold in each ancestry by the number of phecodes tested in that cohort.

The top position of each AM peak was defined by the first position with the most significant *p*-value for a given association, and start and stop positions for the peak interval are defined as the positions at which the *p*-value signal drops below one order of magnitude from the peak. Peaks were plotted and boundaries manually inspected (see example **Supplemental Figure 10**). Permutations, as described previously, were also used to test for any evidence of systematic inflation in the AM PheWAS and to derive lambda value thresholds. Permutations were carried out at four case count thresholds with randomized assignment of cases. We did not observe any systematic inflation of the permutation test statistics with median genomic inflation factor (lambda)=1.01 in AA and lambda=1.02 across each local ancestry background tested in HL respectively (**Supplemental Figures 11-14**). For quality control of PheWAS results lambda value thresholds were established based on ncase=30 permutation results of the corresponding ancestry background tested. AM PheWAS Phecodes with lambda values +/− 3 SD from the median were excluded (AA=3, HL_NAT_=2, HL_EUR_=5, HL_AFR_=7). A summary of thresholds are described in **Supplemental Table 4**). The median lambda value across all HL phecodes tested was 1.03 and 1.03 in AA. Of the 1012 phecodes tested in HL we identified 77 (7.6%) genome-wide significant AM intervals and of the 859 phecodes tested in AA 51 genome-wide significant intervals were identified (5.9%) (**Figure 2**). Genome-wide significant AM peaks ranged in size from 0.07-11.99Mb (median=1.47Mb) in HL and 0.055-13.79 Mb (median=2.99Mb) in AA. Odds ratios (OR) for genome-wide significant AM associations range from 1.18-4.95 (median=1.67) in HL and from 1.28-3.86 (median=1.97) in AA. Three AM peaks on the EUR ancestry background in HL: 282.5 chr11p15.4, 288.1 chr1q23.2 and 290 chr18q23 contained genome-wide significant signal in the same genomic region on the AFR ancestry background with OR in opposite directions (**Supplemental Figures 15-17**). These associations were collapsed such that we report only the association with the positive OR. All genome-wide significant AM results are reported in **Supplemental Table S1 and S2**.

**Figure 2:**
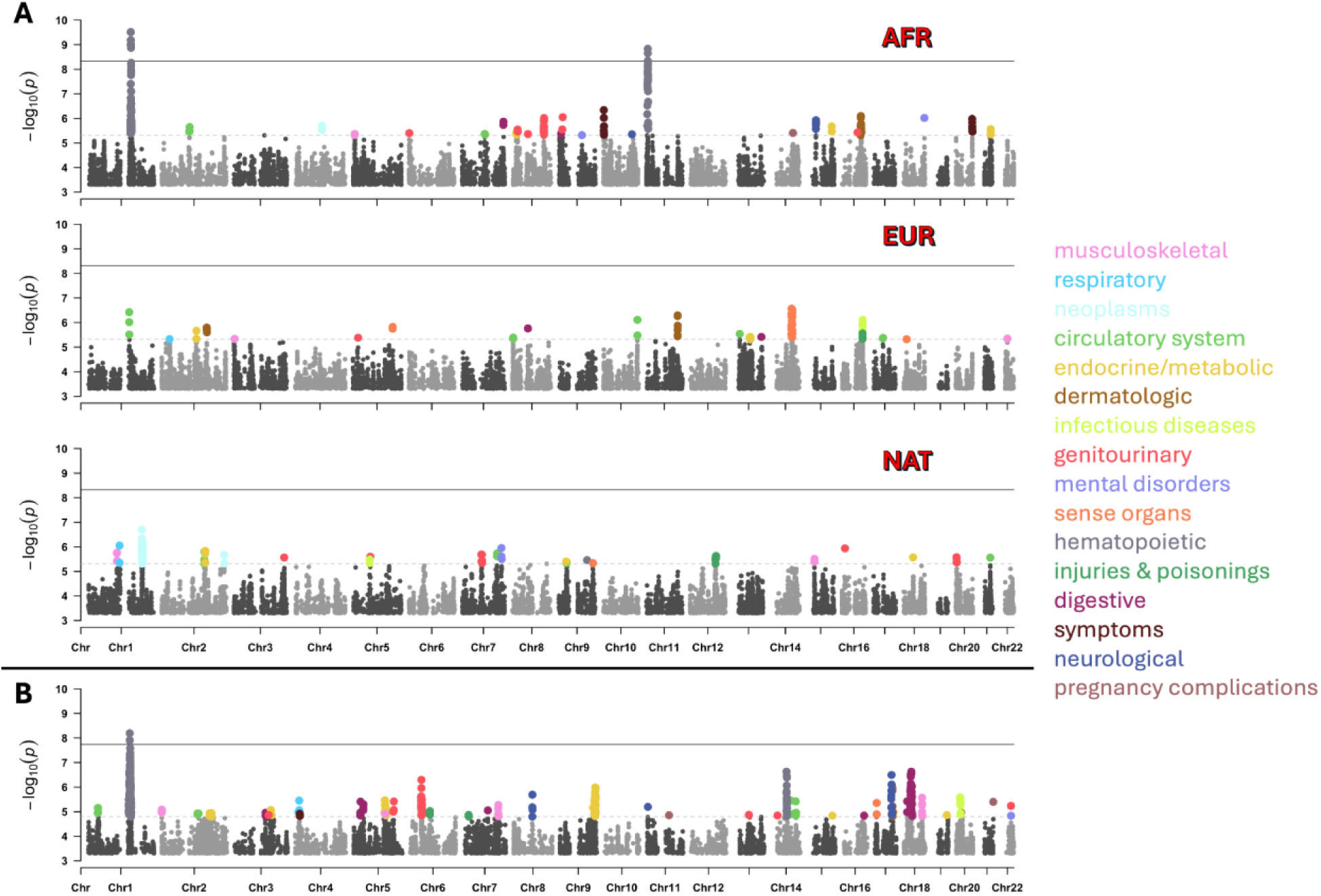
Manhattan plots of admixture mapping PheWAS results in BioMe. Plotting associations with p<5×10^−4^. A) HL association results with each ancestry background represented by a separate Manhattan plot. B) AA association results. Dots represent association signal p-values with peaks passing genome-wide significance colored by phecode categorical labels. The genome-wide significance threshold is indicated by the grey dotted horizontal line, while the study-wide significance threshold is marked by the grey solid horizontal line.

Three AM associations reach study-wide significance (**Table 2**), all three peaks are in regions with established association to phenotypes that have been previously reported. The association between phecode 288.2 “Elevated white blood cell count” and a EUR haplotype at 1q23.1 (Chromosome 1 156.832-159.011Mb GRCh38, *p*<6.41×10^−09^, OR=2.93, 95% CI=2.04-4.2, **Supplemental Figure 18**) in AA replicates known AM signal in the region reported by Nalls et al.^38^ at chromosome 1 158.67-159.57Mb (GRCh38). This association signal was later linked to the *ACKR1* gene, also called the *DARC* gene, at 159.20Mb^38,39^. We saw a similar association in the same region at 1q23.2 between phecode 288.1 “Decreased white blood cell count” and an AFR haplotype in HL (Chromosome 1 158.907-159.795Mb, *p*<3.07×10^−10^, OR=2.19, 95% CI=1.75-2.8, **Supplemental Figure 19**). We note that the duffy null allele (rs2814778) in the *ACKR1* gene, has previously been shown to account for a high degree of variability of white blood cell and neutrophil counts in African-Americans ^39^ and Hispanic Latinos ^40^, and is known to have arisen to high frequency in malaria-endemic regions in Africa and other areas of the world, as this allele also provides resistance to Plasmodium vivax malaria ^41,42^. Of note, the *DARC* locus was not discovered using GWAS approach in the same individuals, though GWAS association signal is observed just below genome-wide significance at the AM tag variant (chr1:159338749:G:A, GWAS p-value = 4.373915×10^−07^, OR=1.88). In addition, a study-wide significant association between phecode 282.5 “Sickle cell anemia” and an African ancestry haplotype in HL (11:4.71-6.00Mb, *p*<1.47×10^−09^, OR=4.95, **Supplemental Figure 20**) overlapping several genes including the *HBB* locus, the primary causal gene for sickle cell anemia, with pathogenic variants most common in African populations. An enrichment of African ancestry at this 11p15.4 locus has previously been reported in a Brazilian Hispanic sickle cell cohort ^43^. Finally, several putatively novel loci emerged, which did not reach study-wide significance, but did achieve genome-wide significance. For example an AM signal located at chr14q31.1 in which an AFR ancestry haplotype is associated with “Missed abortion/Hydatidiform mole” (p<3.92×10^−6^, OR(95%CI)=3.11(1.92-5.04)) in HL females. Fine-mapping implicated variant chr14:80425295:CATAA:C (hg38) within an intronic region of the *DIO2* gene. *DIO2* is highly expressed in first trimester placental villi where it is responsible for thyroid hormone regulation of the fetus ^44,45^. *DIO2* has been identified as being downregulated in the placental villi of those experiencing early recurrent miscarriage ^46^. Characterization of *DIO2* variants and pregnancy loss risk in a population genetics context has not been reported. A full list of the genome-wide significant AM loci is given in **Supplemental Table S1 and S2**.

**Table 2:**
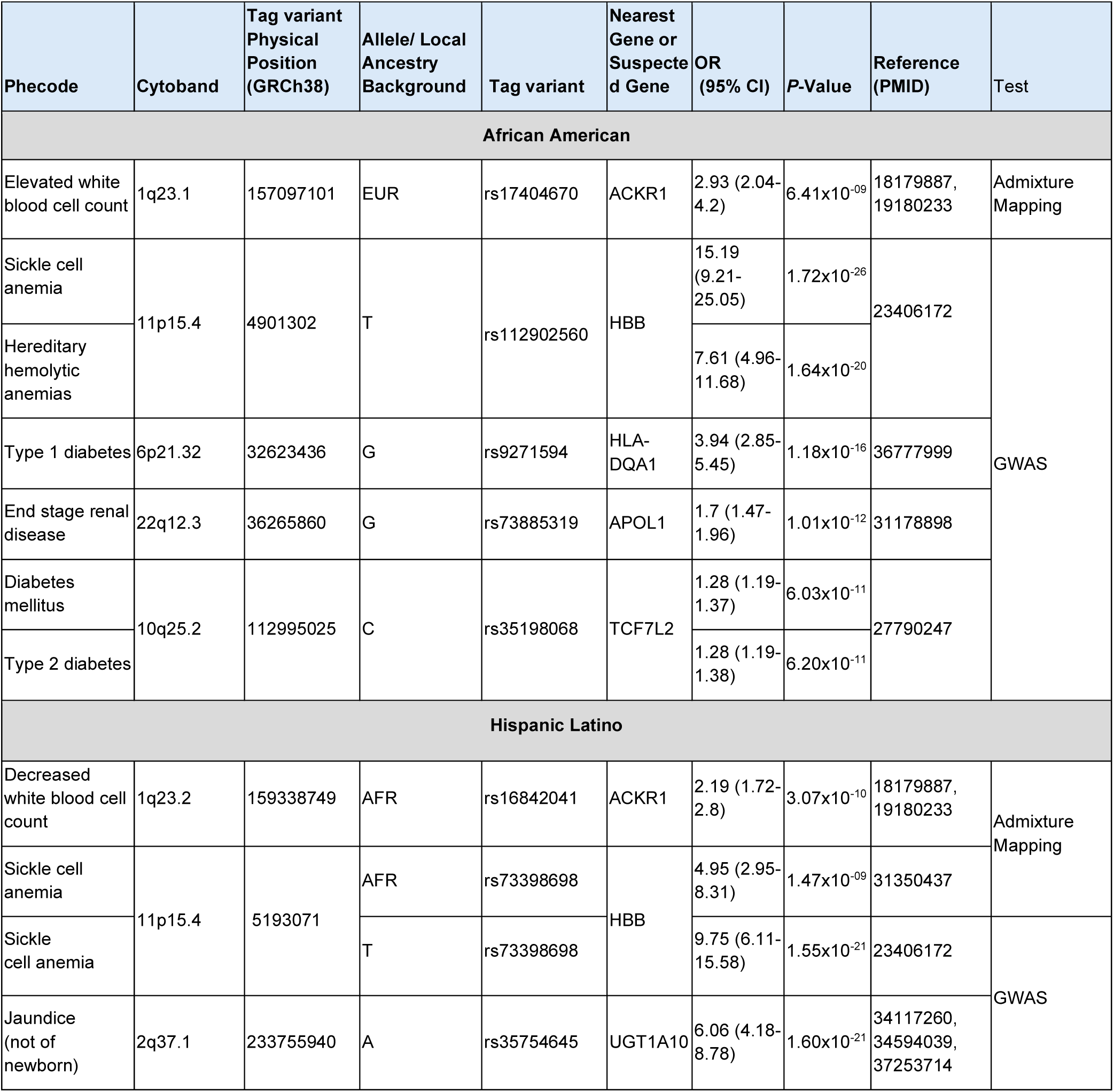
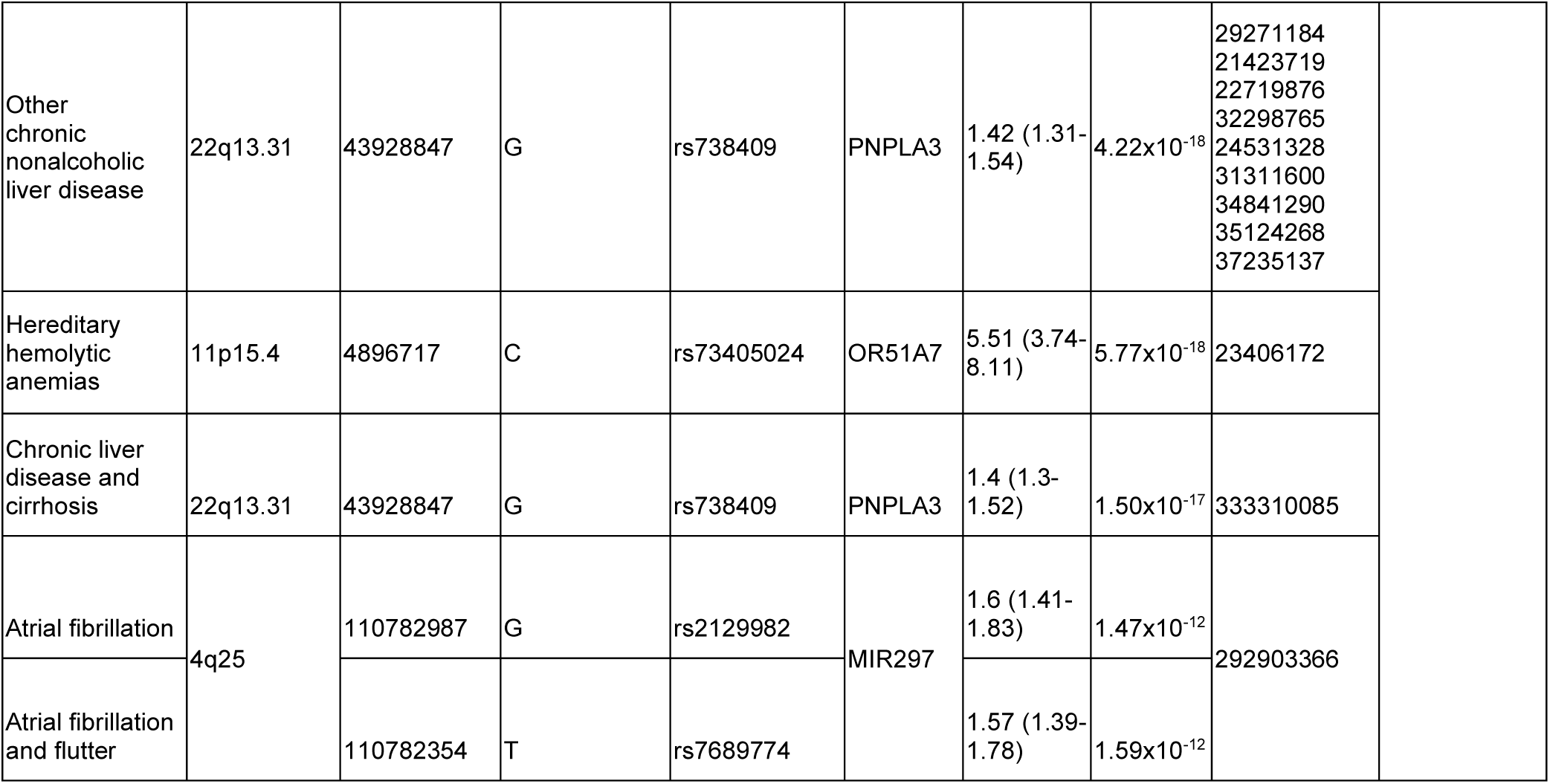
Study-wide significant GWAS and admixture mapping results in BioMe Hispanic Latino (HL) and African American (AA) cohorts.

### Genome-Wide Association PheWAS

GWAS was performed using the same cohorts and phecode definitions used in AM (N_AA_=8,819 and N_HL_=14,876). TOPMed imputed genotypes >1% and passing QC (n_AA_=14,002,950 and n_HL_=11,674,448) were tested for association with each phecode, the allele frequency distributions of variants included in AA and HL GWAS are shown in **Supplemental Figure 21**. HARVESTER package was used for GWAS peak detection and a threshold of p<5×10^−8^ was used to define genome-wide significance. We observed inflation of odds ratios (ORs) at low case counts, particularly when the allele count in cases was <40 (**Supplemental Figure 22**). Therefore, post-hoc we filtered results to remove variant GWAS associations for phecodes in which allele count in cases was <40. We also examined the lambda value for all GWAS associations, and removed phecodes with lambda >1.15 (Phecodes_HL_=49, Phecodes_AA_=0). Lambda values for GWAS passing these post-hoc filters ranged from 0.971-1.13 in HL (median=1.010, mean=1.015) and 0.983-1.15 in AA (median=1.020, mean=1.028). The number of phecodes with in HL was 880, with 13 being male-specific and 67 female-specific. In AA, the number of phecodes was 764, with 9 being male-specific and 55 female-specific. 87 and 73 genome-wide significant peaks were detected in HL and AA, respectively, see **Supplemental Figure 23** for Manhattan plots. Risk OR values of GWAS results passing QC ranged from 1.28-15.19 (median=2.13) in AA and from 1.17-9.75 (median=1.93) in HL. Study-wide significance threshold was defined by dividing the genome-wide significance threshold (5×10^−08^) by the number of phecodes tested in each cohort and was set at p<5.68×10^−11^ for HL and p<6.54×10^−11^ for AA GWAS. Six associations pass study-wide significance in AA and 7 in HL (Table 2, **Supplemental Figures 24-36**). Except for one, all the study-wide significant loci were in a 1MB window of a previously reported association to the same condition and have been reported elsewhere according to the GWAS Catalog (**Table 2**). The exception is the locus at chr2:233755940:ATC:A (rs35754645), associated with phecode 573.5 “Jaundice (not of newborn)” in HL (p<1.6×10^−21^, OR (95% CI)=6.06 (4.18-8.78), MAF_HL_=36.3%, MAF Admixed American gnomAD = 32.5%) which maps to an intronic region of *UGT1A* gene locus. Numerous GWAS have identified variants in *UGT1A* genes to be associated with bilirubin levels in European, East-Asian, and Native American populations^47^. A GWAS including over 9000 individuals of European, African American and Hispanic Ancestry identified rs35754645 as being highly associated with bilirubin levels.^48^ Moreover, *UGT1A1* has been highlighted as a “major locus” influencing bilirubin levels. In a GWAS of African Americans, the variant rs887829 located in the core promoter region of the *UGT1A1* gene was found to explain 12.4% of the variance in serum total bilirubin ^47^. Therefore we speculate that high serum bilirubin levels may underlie the association between rs35754645 and phecode 573.5, as Jaundice is a defining feature of hyperbilirubinemia. Among the genome-wide significant findings, there were several interesting regions, for example between rs13287767 (chr9:130193034:C:G) and phecode 418.1 “Precordial pain” p<2.36×10-8, OR(95%CI)=2.58(1.85-3.6) in HL. This is a compelling signal as the variant is located within an intron of the *NCS1* gene which is known to be involved in regulation of Ca2+ mediated signaling in the heart ^49,50^ All genome-wide significant GWAS results are summarized in **Supplemental Tables S3 and S4**.

To assess the robustness of GWAS results in both AA and HL, we used the phenotype-genotype reference map or PGRM, a curated list of phecode-variant GWAS catalog results developed by Bastarache et. al.^51^. We evaluated replication rates against GWAS studies with reported PGRM statistics: BioVU Biobank: African genetic ancestry (BioVU_AFR_, N=12,142), BioVU Biobank: European genetic ancestry (BioVU_EUR_, N=62,777), Michigan Genomics Initiative biobank (MGI, N=51,393), UK BioBank (UKBB, N=407,202), BioBank Japan (BBJ, N=179,726). Because GWAS findings sometimes do not transfer well across different ancestry backgrounds, PGRM associations are categorized according to 1KGP continental groups. AA GWAS results were compared with “AFR” ancestry PGRM associations (N=22) and the HL GWAS results were compared with “AMR” ancestry PGRM associations (N=19). A summary of results can be found in **Supplemental Table 5.** Replication power was calculated using case rates and allele frequencies in Bio*Me* HL and AA GWAS and the lower OR confidence interval reported in the PGRM to account for winner’s curse ^52^. Of the PGRM AMR GWAS associations, we had >80% power to replicate 16/19 (84%) of the PGRM associations in HL. Of the PGRM AFR GWAS associations, we had >80% power to replicate only 7/22 (31%) of the PGRM associations in AA.

A PGRM association was considered replicated in our cohort if it had a p-value less than 0.05 and an OR with the same direction of effect as reported in the PGRM. Of the 16 PGRM associations we were powered to replicate in HL, 11/16 (69%) associations replicated. Of the 7 PGRM associations we were powered to replicate in AA, only 3/7 (43%) replicated. The reduced power and replication rates in AA may be explained by the relatively low sample sizes for GWAS analysis. Overall these results indicate a lack of power for GWAS in our AA cohort thus we limit the comparison of AM and GWAS results in subsequent analysis to our HL cohort only.

### Systematic comparison of AM and GWAS

We next performed a systematic comparison of AM and GWAS associated loci in the HL cohort. We first investigated the proportion of AM peaks that also contain GWAS statistical signal and the proportion of GWAS peaks that contain AM statistical signal. GWAS signal under AM peaks were compared according to the rubric described in **Supplemental Figure 37**. A threshold of nominal significance is defined as 0.05 divided by the number of GWAS variants tested within the admixture mapping peak. Results are reported in **Figure 3A**. We then compared the AM signal under GWAS peaks using the same rubric, this time defining a 1Mb window around GWAS tag variants to test for AM association signal. Nominal significance was defined as 0.05 divided by the number of admixture mapping haplotypes within each 1Mb GWAS window. Results are reported in **Figure 3B**. We observe that the majority of AM (62.3%, 48/77) peaks contain no nominal GWAS signal in the interval. Additionally, the majority of GWAS peaks (73.6%, 64/87) contain no nominal AM association signal. Under identical sample size and case definitions, AM and GWAS identify different association signals. We next compared the odds ratio (OR) of the top GWAS SNP and the top AM haplotypes. We excluded large effect AM or GWAS peaks that were +/−3SD from the mean, which included GWAS associations for “Sickle cell anemia” (chr11:5193071:C:T, OR=9.75), Jaundice (not of newborn) (chr2:233755940:ATC:A, OR=6.06) and the AM association for “Sickle cell anemia” (11:5225282:T:G, OR=4.95). The median value of OR in AM was 1.34 (sd=0.91) and in GWAS was 1.71 (sd=0.77). OR distributions are significantly higher in GWAS compared to AM in HL (Wilcoxon rank sum test; *p*<2.88×10^−04^_;_ **Figure 4A**). The significant difference in OR between GWAS and AM remains when comparing the lower 95% confidence interval OR to account for winners’ curse (Wilcoxon rank sum test; *p*<1.51×10^−05^; **Supplemental Figure 38**).

**Figure 3:**
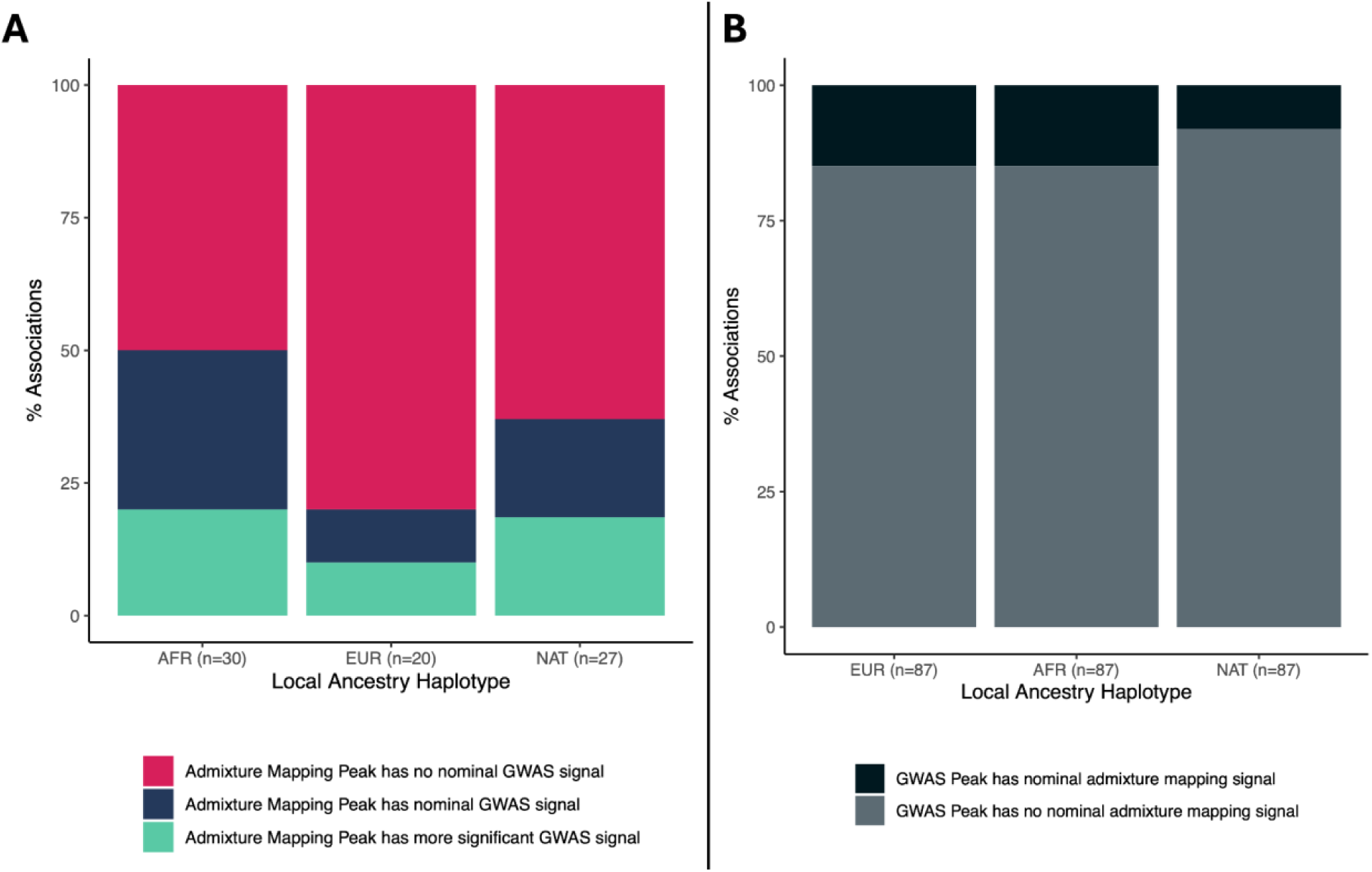
Comparison of association signal at GWAS and admixture mapping genome-wide significant peaks. A) Summary of nominal GWAS signal at genome-wide significant admixture mapping peaks. Nominal significance is defined as 0.05 divided by the number of imputed variants falling within the admixture mapping peak. B) Summary of nominal admixture mapping signal within a 1Mb around each genome-wide significant GWAS peak. Nominal significance is defined as 0.05 divided by the number of admixture mapping haplotypes falling within the 1Mb window.

**Figure 4:**
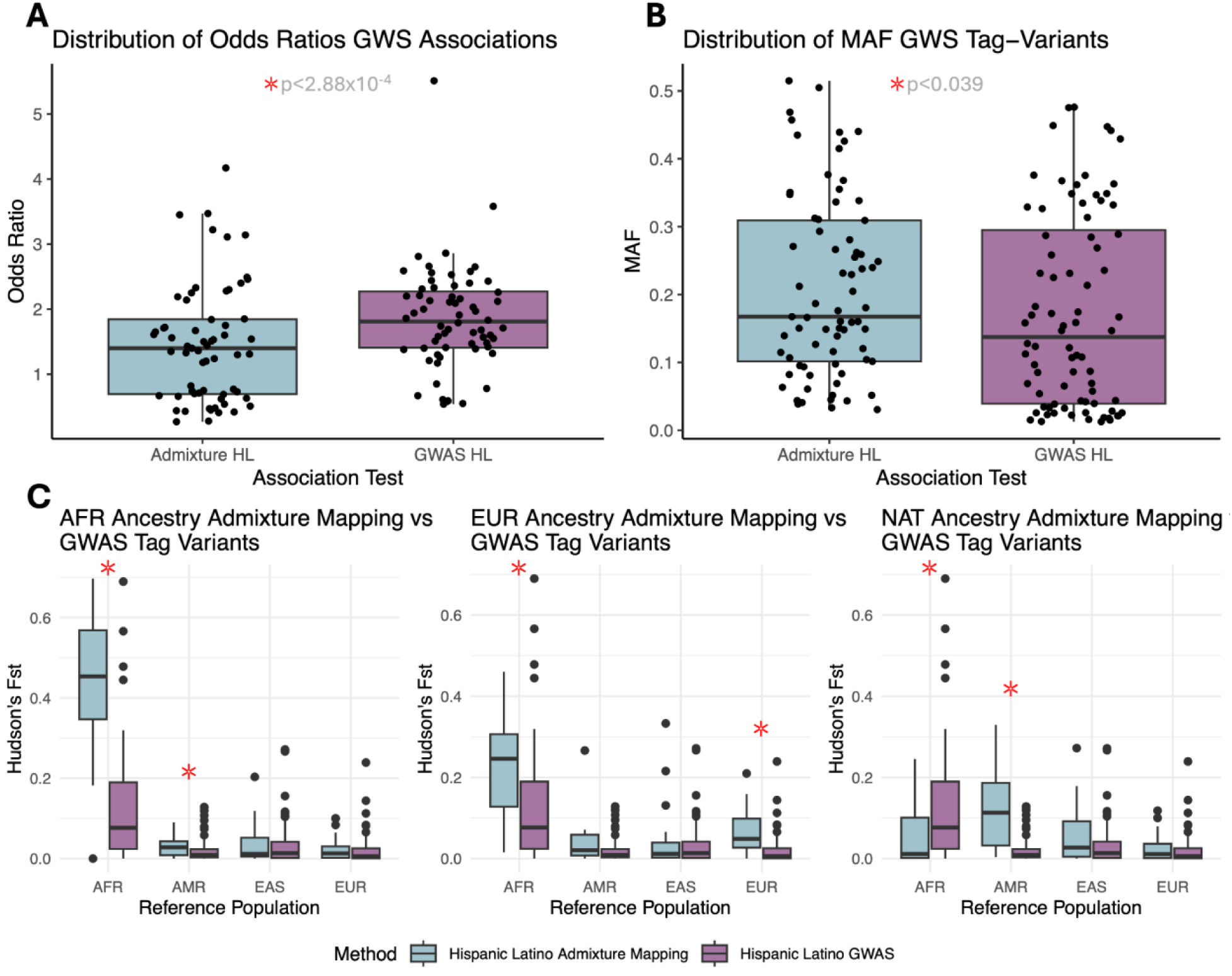
Comparison of genome-wide significant GWAS vs admixture mapping (AM) associations by. **A)** Odds Ratio (OR) of association signal in AM vs GWAS. OR reported are the OR of AM peak and the OR of the GWAS peak **B)** Minor Allele Frequencies (MAF) of fine-mapped tag variant in AM and GWAS tag variant **C)** Fst Values of fine-mapped tag variants in AM and GWAS tag variant. Fst values with significantly different distributions are marked with red asterisk (p value <= 0.05). Note for GWAS the tag variant is the same as the GWAS peak.

To compare AM and GWAS associations at a variant level, we next fine-mapped the AM peaks. Because AM peaks can span regions several Mb in length, we used conditional association testing to fine-map. Within each AM peak, we applied the original AM model, this time conditioning on the TOPMed imputed genotypes at each SNP in the interval iteratively, and the “tag variant” is defined as the SNP that attenuates the AM association signal the most. Of the HL AM associations, 29.9% (23/77) have a tag variant falling within a protein-coding gene, and tag variant and gene mapping information is available in **Supplemental Table S1 and S2**. The top SNP at the GWAS peak was considered the GWAS tag variant for this analysis. The MAF of the AM and GWAS tag variants were compared (**Figure 4B)**. MAF distributions are significantly higher in AM compared to GWAS in HL (Wilcoxon rank sum test, *p*<0.039), the median value of tag variant MAF in AM was 0.17 (sd=0.13) and in GWAS 0.14 (sd=0.14). Finally, we compared variant level Fst values to measure the degree of variant differentiation. Fst is a fixation index with high values indicating the variant is highly differentiated/drifted between the two populations tested ^53^. We estimated variant level Fst values of AM and GWAS tag variants using the 1KGP+HGDP continental populations, calculating Fst values in each population compared to the Central South Asian (CSA) population ^54^. We find that variants identified using AM are significantly more population differentiated compared to GWAS tag variants (**Figure 4C**). For example, tag variants from HL NAT AM tracks (N=25) have significantly higher Fst values compared to GWAS tag variants (n=80) when comparing CSA populations to populations of the Americas (AMR) (*p*<1.41×10^−07^). Similarly HL EUR AM tag variants (n=20) and HL AFR AM tag variants (n=28) have significantly higher Fst values compared to GWAS tag variants when comparing CSA populations to European (EUR) and African (AFR) populations (*p*<4.28×10^−05^ and (*p*<1.38×10^−10^ respectively).

## Discussion

We conducted a systematic comparison of phenome-wide admixture mapping (AM) and genome-wide association studies (GWAS) to investigate the genetic architecture of complex traits in admixed populations. Utilizing genomic data from the Bio*Me* Biobank, we focused on participants self-identified as Hispanic/Latino (HL) and African-American (AA). Our comprehensive pipeline for local ancestry inference (LAI) ensured high-quality ancestry assignments, enabling well-calibrated and accurate AM across hundreds of conditions. We replicated previously known AM and GWAS loci at study-wide significance, demonstrating the robustness of our pipeline. Our analysis revealed that the majority of AM signals were distinct from GWAS signals, with 62% of AM peaks showing no significant GWAS associations. We also demonstrated that AM tag variants showed higher minor allele frequency and population differentiation compared to GWAS tag variants, underscoring the potential of AM to identify variants tied to processes of local selection and adaptation. This work emphasizes the complementary nature of AM and GWAS in elucidating the genetic basis of complex traits in diverse and admixed populations, suggesting different evolutionary mechanisms at play.

The finding that AM and GWAS often pick up different disease associated signals is in line with previous work by Galanter et. al. who, in the context of Asthma, demonstrated that AM and GWAS identified different risk variants in a Hispanic Latino cohort ^31^. One explanation is that AM is able to specifically test for associations based on local ancestry (e.g., African or European tracts within an admixed genome), which directly tests for population-differentiated genetic effects. As variants under historical selection are more likely to be poorly imputed or filtered out via standard GWAS QC for violating Hardy Weinberg equilibrium, AM could provide an orthologous means to detect these signals. On the other hand, the longer AM linkage disequilibrium (LD) blocks make it more challenging to fine-map causal loci at AM peaks. Multiple variants in the same AM LD block may be more common in one ancestral population, making it harder to pinpoint the variant responsible. This may help account for the lower odds ratio in AM vs GWAS tag variants, as AM tag-variants have poorer resolution for the underlying causal variant. One notable example of this was the association of the *HBB* locus with ‘Sickle Cell Anemia’ in HL. Both AM and GWAS approaches pinpointed the rs73398698 tag variant, although the OR was two-fold higher for the GWAS association.

Our study represents a novel phenome-wide application of AM, supported by a robust analytic pipeline for local ancestry inference. The application of this pipeline to phenome-wide AM identified significant associations in 7.6% of tested conditions in HL and 5.9% in AA cohort. Anticipated findings include study-wide significant associations at loci, such as the ACKR1/DARC locus linked to white blood cell counts in both HL and AA. Of note, signal at this locus was below genome-wide significance in the GWAS. On the other hand, some anticipated findings were not seen, for example, variants in the ACKR1 and HBB genes which are known to exhibit highly population-differentiated allele frequencies due to historical selective pressures ^38,39,55,56^. Although a significant GWAS signal at the HBB locus was identified, the AM association signal at the HBB locus in AA did not reach genome-wide significance. This is perhaps due to the overall high proportion of AFR ancestry in the cohort, which may have diluted the signal. Specifically, the proportion of African local ancestry at the HBB locus was 84% in controls and 92% in cases for AA and 25% in controls and 78% in cases for HL. Among the significant AM associations that did not reach study-wide significance, several had interesting biological etiology. However, further validation and replication in independent cohorts are needed to confirm their significance and broader applicability.

Despite the significant findings of the study, several limitations warrant consideration. One limitation of this study is sample size, particularly in the AA, which reduced power to detect associations and limited our ability to compare AA AM and GWAS results. Potential biases, such as population stratification and cryptic relatedness, also pose challenges to the accuracy of LAI and AM. To address these issues, further research should focus on increasing sample size, independently validating findings in other diverse and admixed cohorts and biobanks, and improving the precision of LAI pipelines. Another limitation is the formulation of the test when there are more than two ancestries. In HL, there are three homologous and three heterozygous diplotypes, necessitating the performance of three-way pairwise comparisons which is statistically inefficient. It is also notable in our study that we had to exclude 4% AA and 2% HL individuals as admixture was too complex for current pipelines. Methodological advancements in AM, such as incorporating more comprehensive reference panels, resolving complex LAI, and refining statistical models, are essential for capturing genetic associations more accurately. Studies such as the All of US Research Program will increase sample size and diversity of admixed populations, however interpreting and replicating results may remain challenging due to the complex interplay of multiple sub-group contributions. For example, the HL in Bio*Me* is primarily composed of individuals of Puerto Rican and Dominican heritage, and findings may not replicate in other HL groups with different sub-group composition due to the high population differentiation of AM tag variants.

In conclusion, many phenotypes show population-differentiated effects, but whether these effects reflect genetic or environmental factors can be difficult to elucidate. For the proportion of phenotypic variance that is genetic, AM is a powerful approach for providing insights into genetic diversity influenced by these historical selective pressures. The higher population differentiation and unique association signals captured by AM emphasize its importance in genetic research, particularly in diverse populations that are often underrepresented in traditional GWAS. This study’s findings underscore the need for integrating AM with GWAS to achieve a more comprehensive understanding of genetic influences on health and disease. The methodological advancements and insights gained from this research have the potential to significantly impact future genetic studies and enhance clinical applications, ultimately advancing precision medicine tailored to the genetic backgrounds of diverse populations.

## Methods

### Study population

The Bio*Me* biobank is a Electronic Health Record-linked biorepository that has been enrolling patients in the Mount Sinai Health System in New York City since 2007. There are currently approximately 70,000 participants enrolled in Bio*Me,* under an Institutional Review Board (IRB)-approved study protocol and consent. Participants consent to provide whole-blood derived DNA and plasma samples, which are banked for broad future research use, Participants also complete an extensive questionnaire to answer questions on demographic information, personal and family health history, and social and lifestyle information as has been previously described ^57–59^. Bio*Me* participants represent the broad diversity of New York City, with 65% participants representing minority populations in the United States.

### Genotype data quality control

Bio*Me* participants were genotyped using the Illumina Infinium Global Diversity Array (GDA; number of participants, N=23,430; number of variants, n=1,833,111) or Infinium Global Screening Array (GSA; N=32,595; n=635,623). Quality control (QC) of GDA samples included removal of duplicates and participants with call rate <95% and with mismatch between self-report and genetic sex (N=576). Participants who fell outside +/− 3 standard deviations (SD) of the heterozygosity rate per self-report population group and/or overall were removed (N=276). Duplicated positions (n=553) and variants with genotyping rate <95% were removed (n=12,140). Variants that deviated from Hardy-Weinberg equilibrium (HWE; p<1×10^−08^) within each self-reported population group were also removed (n=119,156). For QC of GSA data samples were stratified by population group and those with a call rate of <95% or heterozygosity rate +/−6 SD of the per ancestry-specific mean were removed (N=684). Duplicate samples and samples with sex discrepancies were also removed (N=206). Variants that had a call rate of <95% and sites that deviated HWE of p<1×10^−05^ African Americans and European American or p<1×10^−13^ in Hispanic/Latinos were also removed (n=11,503). Participants genotyped on the GDA who were also genotyped on the GSA panel were removed to avoid duplicates (N=287). Post QC, the following participants and variants were taken forward for analysis; GDA (N=22,299; n=1,820,156) and GSA (N=31,705; n=604,869).

### TOPMed imputation

Imputation using the TOPMed reference panel freeze 5 was performed using the Michigan Imputation Server. The GDA genotype data was lifted to build GRCh38 prior to imputation to match GSA. GSA and GDA QC’d genotype data was imputed separately and sites with R2 values <0.7 were removed. Of note, a A 10Mb region on chromosome 19 is missing from position 40000000-50000000 and thus is not included in subsequent analysis having failed imputation due to a low reference overlap. These imputed datasets were then combined post imputation using the eMERGE consortium framework described in ^60^ and the final imputed dataset comprised N=53,449 participants and n=60,491,206 variants.

### Ascertainment of self-reported heritage information and strategy to define population groups

Of the Bio*Me* participants in the combined imputation dataset, we extracted information from a multiple choice question about their heritage asked at enrollment. Participants could choose one or multiple of the options. The survey question and the choice of options are detailed in **Supplemental Figure 2**. Participants were also asked whether they self-reported as ‘Hispanic Latin American’, and could choose yes or no. Finally, participants were given the option to self-report their country of birth. Participants’ responses were summarized, and if participants selected ‘Hispanic Latin America’ and one or multiple options from the heritage question, they were assigned to a population group with the population label Hispanic/Latino (HL) (N=17,150). If a participant selected ‘African-American/African’ or ‘African-American/African’ and one or multiple other options from the heritage question, they were assigned AA (N=10,160). The answer to the country of birth question was not used to assign population groups, but is given to provide contextual information about the participants in the study. Groups of individuals who answered survey questions with ≤20 participants were collapsed to protect confidentiality.

### Global genetic similarity with reference panels

Global ancestry proportions were calculated using the imputed dataset. Samples from previously published reference panels were merged with the imputed dataset to identify samples from reference panels to be used as reference samples for local ancestry inference (LAI; see Local Ancestry Inference section). This included two publicly available genome sequencing (GS) reference panels; The 26 populations of the 1000 Genomes Project (1KGP; N=3202, n=75,310,370) and the 54 populations of the Human Genome Diversity Project (HGDP; N=929, n=75,310,370)^61,62^. In addition, a GS data was also available for an unpublished reference panel, comprising a subset of samples from the Women’s Health Initiative (WHI) (N=235,n= 33,590,559) and Stanford Global Reference Panel (SGRP) (N=78, n= 14,421,461), as well as a small cohort of Bio*Me* participants (N=307, n=48,189,246) sequenced as part of the Population Architecture using Genomics and Epidemiology (PAGE) Study (n=519,648) ^63–65^. Firstly, the imputed dataset (N=53,449, n=60,491,206) was filtered to keep individuals assigned to HL and AA groups as described above and retain sites that were genotyped on the GDA panel (N=27,310, n=938,671), and these positions were also extracted from the reference panels. The datasets were merged using PLINKv1.90 ^66^ (N=32,061, n=479,586). Downsampling to the GDA sites was performed to enhance computational efficiency with the panel developed specifically to capture global genetic diversity. PLINKv2.00 ^67^ was used to filter variants with genotyping rate <95%, MAF < 1% as well as duplicate samples and samples with missingness >5% (N=291 individuals and n=12,666 variants were removed). Variants in regions that are under recent selection were removed, namely human leukocyte antigen region (chr6:28,510,120-33,480,577,hg38), lactase gene (chr2:134,242,429-136,242,430), an inversion on chromosome 8 (chr8:6,142,478-16,142,491), a region of extended LD on chromosome 17 (chr17:41,843,748-46,922,634), EDAR (chr2:108,383,544-109,383,544), SLC2A5 (chr15:47,707,803-48,707,803), TRBV9 (chr7:142,391,891-142,392,412) (n= 11,398 variants removed). Palindromic sites were removed and LD pruning was performed using the PLINKv2.00 flag ––indep-pairwise 50 5 0.2 (n=268,825 variants in total removed). Following these filtering steps, the combined AA and HL and reference panels comprised N=31,771 participants and n=186,697 variants. This dataset was used to infer individual-level genetic similarity between the AA and HL to the reference populations using ADMIXTURE software v1.22^68^. Principal components (PCs) were also calculated using this dataset for addition as covariates in downstream association testing using PLINKv2.00 –pca command. Samples with AA participants with >5% genetic similarity with ancestry components other than European or African reference panels at K=3 were removed (N=356). HL participants with >5% genetic similarity with East Asian (based on Chinese Dai in Xishuangbanna, China) genetic ancestry at K=4 were excluded (N=271). This resulted in sample sizes of N=16,733 HL and N=9,660 AA for LA inference. Confidence intervals around median global ancestry proportions were calculated using the “boot” R package, with N bootstrap replicates=1000 ^69^.

### Selecting reference panels for local ancestry inference

The following strategy was used to select reference populations for LAI. Samples with ≥ 90% enrichment for the ADMIXTURE component seen in the Native American reference population at K4 were selected (N=154). The same process was applied to samples with ≥ 90% enrichment for ADMIXTURE components seen in European and African reference panels at K4. Because balanced representation from each source population is recommended for local ancestry inference, the European and African reference panels were downsampled to include only CEU and YRI samples respectively. This resulted in reference panel sample sizes of Native American=154, CEU=178 and YRI=178. The selected reference panels and AA and HL populations post QC based on ADMIXTURE components were extracted from the merged dataset pre ADMIXTURE protocol (n=479,586). PLINKv2.00 ––king-cutoff 0.125 was used to remove one of each pair of second degree relatives from the reference panel, AA and HL (N= 2,348).

### Local ancestry inference

Eagle v2.4 was used to phase the reference panel, AA and HL participants (N=24,554 and n=479,586) using the hapmapII genetic map and default parameters. Variants with missingness >10% were removed (n=1,026)^70^. Finally, local ancestry was called using GNOMIX^24,25^. Phased genotype calls (n=478,560) were then split into reference (CEU, YRI and Native American reference panel) for training the GNOMIX model and query VCF files (N_HL_=15,149 and N_AA_=9,022). GNOMIX was used using default parameters defined in the GNOMIX config file for array data, with window size of 0.2cM, to call two-way local ancestry in AA, CEU and YRI reference panels were used to infer EUR and AFR ancestry tracks respectively. Three-way local ancestry was inferred in HL, inferring EUR and AFR ancestry tracks, along with the Native American reference panel used to infer NAT ancestry tracks. Chromosomes 7, 13 and 19 needed adjustment of window size to 0.25cM in order to successfully run without error. GNOMIX estimated the average trained model accuracy to be 99.27% for AA and 98.05% for HL. QC of local ancestry calls involved comparing the proportion of the genome assigned to a specific local ancestry component with global ancestry estimates. To do this in AA, the physical distances of local ancestry calls for a given ancestry component (e.g., all AFR local ancestry haplotypes) were summed and then divided by the total physical distance. This proportion was compared to the YRI ancestry component for that individual at K=3 of the global ancestry estimates, similarly for European local ancestry calls and the CEU ancestry component. The same process was applied to AFR, EUR and NAT haplotypes in HL individuals, using K4 global ancestry components. Overall correlation between global ancestry and local ancestry for all HL and AA LAI calls was 1. 2 samples deviated from global ancestry proportions greater than 5% in the HL cohort. 58 deviated from global ancestry proportions greater than 5% in the AA cohort (max difference 7%). These outlier samples were removed from further analysis. The density of each ancestry component along the genome was plotted and regions with enrichment of one ancestry component +/− 4SD of the median were removed from LAI calls. Centromere positions were downloaded from UCSC Genome Browser (https://genome.ucsc.edu/cgi-bin/hgTables;Group: Mapping and Sequencing, Track: Centromeres, Table Centromeres) and LAI calls within centromeric regions were removed.

### Extracting phecodes from the Electronic Health Record

International Classification of Diseases 9th Revision (ICD-9) and ICD-10 codes were extracted from the Mount Sinai Health System electronic health records (EHR) records for Bio*Me* participants on Oct 4 2021. ICD codes were converted to phecodes (v1.2) using the PheWAS R package using the createPhenotypes() function with min.code.count=2 and add.phecode.exclusions=F). Cases and controls were encoded as 0 and 1 respectively and samples with one instance of an ICD code were assigned “NA”.

### Permutations to determine minimum number of cases needed and lambda value thresholds for admixture mapping PheWAS

The stability of association test statistics and genomic inflation of the AM PheWAS was empirically tested using permutations at different minimum number of case counts. AM using the HL (N=8,819) and AA (N=14,876) cohorts was performed with randomized case/control assignments at four different N case thresholds: 30, 40, 50 and 100. In HL each local ancestry population was tested, i.e NAT, EUR and AFR local ancestry haplotypes tested separately. For each permutation (p=100) at a given case count, the *p*-value at the 95th percentile was plotted. For calculation of genomic inflation (lambda) values in both AA and HL AM permutations, a single p-value from each tested AM haplotype was used.

### Admixture mapping PheWAS

GNOMIX outputs an individual’s LAI (0 = NAT, 1= EUR, 2= AFR) at each variant in the input VCF. LAI calls were converted into VCF genotype style calls for association testing using SAIGEv1.1.6.2^71^. Separate VCF files were constructed for each LAI component, for example, HL LAI calls were split into three separate VCF files corresponding EUR, AFR and NAT LAI haplotypes. All variant calls assigned to a given LAI component e.g. NAT were encoded as 1 and all variant calls assigned a different LAI component (i.e. EUR and AFR) were assigned 0, thus a given individual at a given variant was assigned an “LAI genotype” of either 0/0, 0/1 or 1/1. These VCF files are referred to subsequently as “LAI VCFs”. A single LAI VCF was used for AA individuals with EUR LAI encoded as “1” and AFR LA encoded as “0”. In AAs, as admixture is two-way, a single association test per phecode was performed and association results were reported in terms of EUR LAI. Three separate sets of AM was performed in HL due to three-way admixture: NAT vs non-NAT, EUR vs non-EUR, AFR vs non-AFR. The association results for each set of AM in HL is reported in terms of the test ancestry e.g. in NAT vs non-NAT admixture mapping OR are reported in terms of NAT LAI haplotypes.

Association testing was performed in AA and HL cohorts separately. SAIGE was used to construct a genetic relatedness matrix (GRM) using the same genotype calls as were used as input for LAI. Sites were pruned using PLINKv2.00 —indep-pairwise 100 5 0.1 and a MAF of 1% was applied (n_HL_=171,935 and n_AA_= 235,249). The null logistic mixed models were then fit separately for each phecode using the full GRM with phecode case status as the binary phenotype along with the covariates *AGE + SEX + Genotype Chip + PC 1-10* (N_HL_=14,876 and N_AA_=8,819). The parameters –skipVarianceRatioEstimation=FALSE ––IsOverwriteVarianceRatioFile=TRUE –– isCovariateOffset=TRUE were used for fitting the null model. LAI VCF files were used in SAIGE step 2 for association testing according to the model: *phecode ∼ Local Ancestry at SNP X (0/0 or 0/1 or 1/1) + Age + Sex+ Genotype Chip + PC1-10* with the parameters —minMAF=0 –– minMAC=0.5 –LOCO=TRUE ––is_Firth_beta=TRUE ––pCutoffforFirth=0.05.

Only phecodes with ≥ 30 cases were tested (Phecodes_AA_=859 and Phecodes_HL_=1,012). For male and female specific phecodes (for AA, Phecodes_female specific_=68 and Phecodes_male specific_=10; for HL = 81 Phecodes_female specific_ and 17 Phecodes_male specific_) only individuals of the corresponding sex were included for testing and sex was removed as a covariate in the null model. Lambda values thresholds were defined based on lambda value distributions of the ncase=30 AM permutations in HL and AA. The lower and upper lambda cutoffs for the AM PheWAS were defined as ±3 standard deviations from the median permutation lambda. The STEAM^33^ R package was used to derive a genome-wide significance threshold for admixture mapping in HL and AA cohorts. QCd LAI calls were used to derive the significance threshold using the get_thresh_simstat() function with nreps = 10000. One SNP per 0.2 Cm window was used to estimate the correlation of local ancestry across loci. This correlation file was used to calculate the number of generations since the admixture (g) using the get_g() function (AA g =9, HL g = 10). Genome-wide significance significance thresholds of *p*<1.598×10^−05^ and *p*<4.876×10^−06^ were calculated for AA and HL, respectively. A Bonferroni study-wide *p*-value threshold defined as the genome-wide threshold divided by the number of phecodes tested was *p<*1.86×10^−08^ and *p*<4.82×10^−09^ for AA and HL, respectively.

AM results were processed using in-house R scripts. We examined regions that exceeded genome-wide significance to define the boundaries of the peaks. The SNP at the start of the ancestral haplotype with the lowest admixture mapping *p*-value was considered the “index LAI SNP” for a given phecode association. The range of each admixture-phenotype genome-wide significant peak was defined by the inclusion of all variants upstream or downstream of the index LA SNP as long as their *p*-values were within one order of magnitude of the index LA SNP. In HL three peaks had genome-wide significant AM signal at the same position on two local ancestry backgrounds with opposite directions of effect: phecode 282.5 chr11p15.4, phecode 288.1 chr1q23.2 and phecode 290 chr18q23. These association peaks were collapsed and we report only the association with a risk OR.

### Genome-wide association testing

GWAS was performed for each phecode tested in the Admixture mapping PheWAS (Phecodes_AA_=859, Phecodes_HL_=1,012). The full set of merged TOPMed imputed in the combined dataset were included in each GWAS. Sites were filtered using PLINKv2.00 ^67^ to only include variants with a MAF >1%, genotyping rate of >95% and sample missingness rate <5%. Single-variant association tests were carried out using SAIGE in HL (N=14,876, n=11,674,448) and AA (N=8,819, n=14,002,950). Each null model was fit using the same pruned genotype calls used for construction of the full GRM in the Admixture mapping PheWAS (n_HL_=171,935, n_AA_=235,249). The model for association testing was: Phecode ∼ Genotype SNP X + Age + Sex + Genotype Chip + PCs 1-10 + GRM. Results were scanned for GWAS peaks using the Manhattan Harvester V0.1 software^72^ using default parameters –peak-limit=5, –dots=5. Manhattan Harvester results were filtered to retain peaks with a p<5×10^−8^. The variant with the lowest p-value was defined as the “tag variant” for each GWAS peak. An additional post hoc variant-level filter of allele count in cases <40 and a phecode-level filter to remove phecodes where the QQplot had lambda>1.15 were applied. The final number of phecodes tested were (Phecodes_AA_=764, Phecodes_HL_=880).

### Conditional Association Testing

Conditional association testing was used to fine-map genome-wide significant admixture peaks to identify likely tag variants driving the admixture mapping signal in both AA and HL. The model used was similar to the original admixture mapping analysis however the LAI calls at the index LAI SNP remained fixed as a covariate. Genotype calls for imputed TOPMed sites falling within the admixture peak were iteratively included as a covariate in the model for conditional analysis, testing each variant separately, using the same discovery cohorts. The GRM for the regression was fit with the same full GRM using genotype calls as described previously. Conditional analysis at each gws admixture peak was performed using the model: *Phecode ∼ Local Ancestry at index LA SNP* + *Age + Sex +* Genotype Chip + *PCs1-10 + TEST SNP GENOTYPE X + full GRM*. Results are compared across all tested SNPs within a given admixture mapping peak. The “tag variant” defined as the variant that, when included as a covariate, increases the *p*-value of the index LA SNP by the greatest amount.

### Estimating Power for Replication using the PGRM

The phenotype-genotype reference map (PGRM)^51^ defines variant-phecode associations curated from the GWAS-catalog that can be used for determining replication rates in a cohort. We used this resource to assess power for replication in the AA and HL GWAS. Associations reported in the PGRM are assigned to 1000 Genomes superpopulations ancestry groupings based on the discovery study cohort. The PGRM was downloaded and associations with admixed American (AMR) or African (AFR) ancestry groupings were retained to determine replication rates in the HL and AA GWAS, respectively. We restricted our comparison to PGRM associations where the phecode had n_cases_ ≥ 40 in the corresponding HL or AA GWAS. We calculated power to replicate each phecode matched PGRM association in the corresponding HL and AA GWAS using the genpwr R package. To calculate power we used the number of samples (N), case rate, and MAF from the corresponding HL or AA GWAS. The odds ratio (OR) used for each power calculation took the lower 95% confidence interval of the OR reported in the matched PGRM association. A replication was defined as variant-phecode association with a p<0.05 and OR in the same direction as the original PGRM reported study. Replication statistics are described in ^51^. Briefly, %Power is the percentage of matched PGRM associations which we had >=80% power to replicate compared to the total number of matched PGRM associations tested. Overall replication rate (RR_All_) is defined as the overall proportion of matched PGRM associations replicated in our study. Powered replication rate (RR_Powered_) is the proportion of matched associations replicated in our study but only considering associations for which we had >=80% power (α=0.05) to replicate

### Variant Annotation

Nearest gene and transcript information of tag variants were annotated using HOMER ^73^. Cytoband information was annotated using the cytoBand.txt.gz file downloaded from https://hgdownload.soe.ucsc.edu/goldenPath/hg38/database/ ^74^. Annotation of rsIDs was applied using the Ensembl REST API (http://rest.ensembl.org).

### Fst Calculation

Fst values were calculated using the harmonized 1000 Genomes Project (1KGP) and Human Genome Diversity Project (HGDP) call set from Martin et. al.^54^. Relatedness of samples within the HGDP+1KGP cohort was estimated based on a LD pruned set of 171,628 variants with a MAF > 5%. One member of each pair of samples with kinship coefficient greater than 0.125 were removed using the ––king-cutoff 0.125 command in Plinkv2.00 (N= 686). The 5 largest harmonized population groups were used to estimate Fst values; AFR (Africa N= 772), AMR (Americas N= 396), CSA (Central South Asia N= 695), EAS (East Asia N= 740), EUR (Europe N= 678). Variant-level Fst values between each population group were calculated using Plinkv2.00^67^ with the command ––fst POP method=hudson report-variants ––pheno popgroup.pheno. For statistical analysis, we used variant Fst values between CSA and each of the other four population groups for each genome-wide significant variant in the HL AM and GWAS studies. All HL AM and GWAS tag variants were present in the HGDP+1KGP dataset for variant level Fst estimation. Fst values less than 0 were winsorized to 0.

### Plots

PheWAS Manhattan plots were made using the “CMplot” ^75^ R package. All additional supplemental Manhattan and quantile-quantile (qq) plots were made using the “qqman” R package^76^. The “topr” R package was used to make locus zoom style plots of AM results ^77^. All additional plots were made using “ggplot2” in R.

## Data and Code Availability

Pipeline scripts and summary statistics can be found at https://github.com/sinai-igh/admix_phewas/

## Data Availability

Pipeline scripts and summary statistics can be found at https://github.com/sinai-igh/admix_phewas/

## Acknowledgements

Preprocessing of Bio*Me* genotype data and imputation was performed by the Charles Bronfman Institute for Personalized Medicine genomics team. The authors are grateful to the patients and their families who contributed to this study.

## Funding

This work was supported in part through the computational and data resources and staff expertise provided by Scientific Computing and Data at the Icahn School of Medicine at Mount Sinai and supported by the Clinical and Translational Science Awards (CTSA) grant UL1TR004419 from the National Center for Advancing Translational Sciences. Research reported in this publication was also supported by the Office of Research Infrastructure of the National Institutes of Health under award number S10OD026880 and S10OD030463 and National Institute on Minority Health and Health Disparities award number R21MD019104. The content is solely the responsibility of the authors and does not necessarily represent the official views of the National Institutes of Health. The data in this paper were used in a dissertation as partial fulfillment of the requirements for a PhD degree at the Graduate School of Biomedical Sciences at Mount Sinai.

## Competing interests

EK received personal fees from Illumina, 23andMe, Allelica, and Regeneron Pharmaceuticals, received research funding from Allelica, and serves as a scientific advisory board member for Encompass Bio, Foresite Labs, and Galatea Bio.

## Contributions

EK, SC, SA and RS participated in study design. SC and RS performed bioinformatic and statistical analysis. All authors drafted the manuscript, provided critical edits and approved the final manuscript.

## Notes

### Author Declarations

This study was approved by the Icahn School of Medicine at Mount Sinais Institutional Review Board (Institutional Review Board 07 0529). All study participants provided written informed consent.

