## Supplementary Materials (Figures and Tables) for "Systematic comparison of phenome-wide admixture mapping and genome-wide association in a diverse biobank"

This file includes:

**Supplemental Figure 1:** Flowchart overview of methods pipeline with sample and variant sizes.
**Supplemental Figure 2:** Bio*Me* survey questions regarding participants' heritage asked at enrollment.
**Supplemental Figure 3**: Admixture plots illustrating K3 and K4 global ancestry components A) pre-quality control and B) post-quality control
**Supplemental Figure 4**: Correlation between global ancestry components and sum of physical distances of local ancestry calls
**Supplemental Figure 5:** Permutation p-value histograms per case count - African American
**Supplemental Figure 6.** Permutation p-value histograms per case count - Hispanic Latino (HL) AFR ancestry
**Supplemental Figure 7**: Permutation p-value histograms per case count - Hispanic Latino EUR ancestry
**Supplemental Figure 8**: Permutation p-value histograms per case count - Hispanic Latino NAT ancestry
**Supplemental Figure 9**: Minor allele frequency (MAF) distribution plots - admixture mapping
**Supplemental Figure 10**: Example of admixture mapping peak definition.
**Supplemental Figure 11:** Permutation lambda value histograms per case count - African American
**Supplemental Figure 12:** Permutation lambda value histograms per case count -Hispanic Latino AFR ancestry
**Supplemental Figure 13**: Permutation lambda value histograms per case count -Hispanic Latino EUR ancestry
**Supplemental Figure 14**: Permutation lambda value histograms per case count -Hispanic Latino NAT ancestry
**Supplemental Figure 15:** Manhattan plots, admixture mapping results Hispanic Latino phecode 282.5
**Supplemental Figure 16:** Manhattan plots, admixture mapping results Hispanic Latino phecode 288.1
**Supplemental Figure 17:** Manhattan plots, admixture mapping results Hispanic Latino phecode 290
**Supplemental Figure 18:** Region plot- admixture mapping and GWAS results Hispanic Latino phecode 288.2
**Supplemental Figure 19:** Region plot- admixture mapping and GWAS results Hispanic Latino phecode 288.1
**Supplemental Figure 20:** Region plot- admixture mapping and GWAS results African American phecode 282.5
**Supplemental Figure 21:** Minor allele frequency (MAF) distribution plots - GWAS
**Supplemental Figure 22**: Distribution of odds ratios (OR) across different values of allele counts in cases for genome wide significant GWAS associations
**Supplemental Figure 23:** Manhattan plots of GWAS phenome wide association study (PheWAS) results
**Supplemental Figure 24-30**: Manhattan and quantile quantile plots, study wide significant Hispanic Latino GWAS results

- Phecode 571 - chronic liver disease
- Phecode 282 - hereditary hemolytic anaemias

- Phecode 282.5 - sickle cell anaemia
- Phecode 427.21 - atrial fibrillation
- Phecode 427.2 - atrial fibrillation and flutter
- Phecode 571.5 - other chronic nonalcoholic liver disease
- Phecode 573.5 - jaundice (not of newborn)

**Supplemental Figure 31-36**: Manhattan and quantile quantile plots, study wide significant African American GWAS results

- Phecode 282 - hereditary hemolytic anaemias
- Phecode 282.5 - sickle cell anaemia
- Phecode 250.1 - type 1 diabetes
- Phecode 585.32 - end stage renal disease
- Phecode 250 - diabetes mellitus
- Phecode 250.2 - type 2 diabetes

**Supplemental Figure 37:** Decision tree for main text Figure 2A
**Supplemental Figure 38:** Boxplot showing the distribution of the lower 95% confidence interval of odds ratios (OR) from admixture mapping vs GWAS results.

**Supplemental Table 1:** Summary of Bio*Me* participants' responses to survey questions categorized by Hispanic Latino (HL) and African American (AA) cohorts.
**Supplemental Table 2:** Population descriptors and abbreviations of samples ascertained from reference panels.
**Supplemental Table 3:** Description of reference panel names, population labels and sample sizes selected for local ancestry inference
**Supplemental Table 4:** Summary of lambda value distributions from ncase=30 admixture mapping permutations used to establish thresholds for admixture mapping PheWAS results.
**Supplemental Table 5:** Phenotype-genotype reference map (PGRM) replication summary statistics

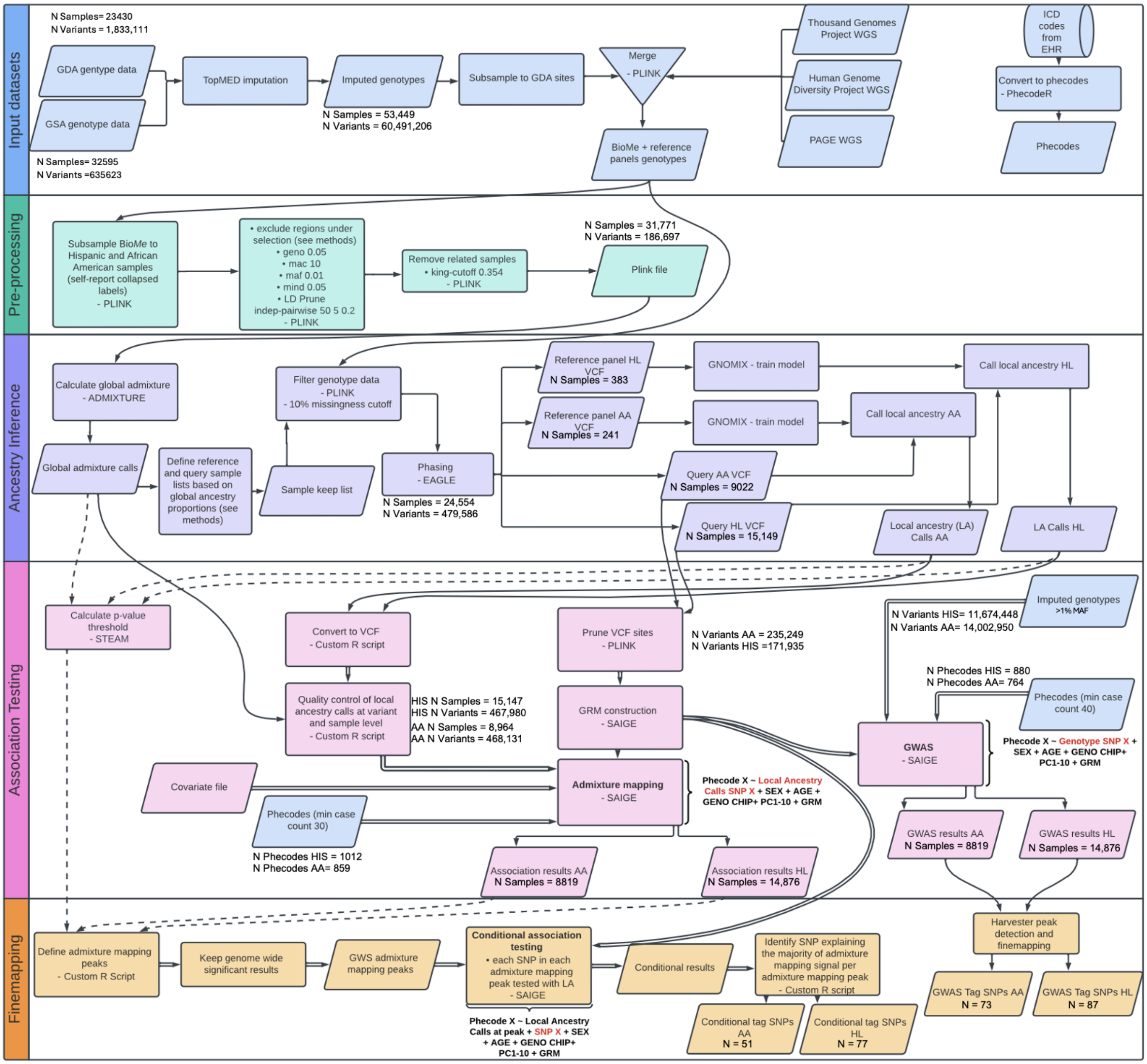

**Supplemental Figure 1:** Flowchart overview of methods pipeline with sample and variant sizes.

**A)**

**Which of the following best describes your heritage? (tick all that apply)**

| |
|---|
| American Indian, Native American or Alaskan Native |
| African-American / African |
| Caucasian / White |
| Mediterranean |
| East or Southeast Asian |
| South Asian/Indian |
| Jewish |
| Native Hawaiian or Other Pacific Islander |
| Other: please specify |

B) **Are you Hispanic/Latino?**

No 0 **GO TO QUESTION 4**

Yes 1 **GO TO QUESTION 5**

C) **In what country were you born?** ____________________________

**Supplemental Figure 2:** Bio*Me* survey questions regarding participants' heritage asked at enrollment.

| Answer Combination | Sample Size (N) | Percentage |
|---|---|---|
| **Hispanic/Latino HL (N= 17,150)** | | |
| **Heritage** | | |
| Hispanic-Latin-American | 15989 | 93.23 |
| Hispanic-Latin-American, African-American/African | 424 | 2.47 |
| Hispanic-Latin-American, Caucasian/White | 396 | 2.31 |
| Hispanic-Latin-American Mediterranean | 90 | 0.52 |
| Hispanic-Latin-American Other | 69 | 0.40 |

| | | |
|---|---|---|
| Hispanic-Latin-American, American-Indian/Native-American | 53 | 0.31 |
| Hispanic-Latin-American, East-or-Southeast-Asian | 48 | 0.28 |
| Other | 81 | 0.47 |
| **Country of Birth** | | |
| USA | 6972 | 40.65 |
| Puerto Rico | 3666 | 21.38 |
| Dominican Republic | 2795 | 16.30 |
| Ecuador | 834 | 4.86 |
| Colombia | 803 | 4.68 |
| Mexico | 535 | 3.12 |
| Peru | 249 | 1.45 |
| Honduras | 202 | 1.18 |
| Cuba | 199 | 1.16 |
| El Salvador | 123 | 0.72 |
| Guatemala | 119 | 0.69 |
| Venezuela | 111 | 0.65 |
| Brazil | 82 | 0.48 |
| Argentina | 81 | 0.47 |
| Panama | 69 | 0.40 |
| Chile | 62 | 0.36 |
| Nicaragua | 40 | 0.23 |
| Bolivia | 34 | 0.20 |
| Costa Rica | 33 | 0.19 |
| Spain | 23 | 0.13 |
| other | 117 | 0.68 |
| **African-American AA (N= 10,160)** | | |
| **Heritage** | | |
| African-American/African | 10024 | 98.66 |
| African-American/African Other | 101 | 0.99 |

| | | |
|---|---|---|
| African-American/African Mediterranean | 26 | 0.26 |
| Other | 9 | 0.09 |
| **Country of Birth** | | |
| USA | 8504 | 83.70 |
| Jamaica | 384 | 3.78 |
| Trinidad & Tobago | 155 | 1.53 |
| Haiti | 132 | 1.30 |
| Guyana | 106 | 1.04 |
| Barbados | 75 | 0.74 |
| Ghana | 74 | 0.73 |
| Senegal | 61 | 0.60 |
| Nigeria | 56 | 0.55 |
| Belize | 37 | 0.36 |
| Ivory Coast | 35 | 0.34 |
| Grenada | 34 | 0.33 |
| Antigua & Barbuda | 33 | 0.32 |
| Guinea | 32 | 0.31 |
| United Kingdom | 28 | 0.28 |
| Sierra Leone | 25 | 0.25 |
| St. Thomas | 24 | 0.24 |
| St. Lucia | 24 | 0.24 |
| Virgin Islands (U.S. or British) | 20 | 0.20 |
| St. Kitts & Nevis | 21 | 0.21 |
| Panama | 20 | 0.20 |
| Other | 280 | 2.76 |

**Supplemental Table 1:** Summary of Bio*Me* participants' responses to survey questions (refer to Supplemental Figure 2) categorized by HL and AA cohorts chosen for association testing.

| Population Descriptor | Abbreviation |
|---|---|
| **Thousand Genomes Project (1KGP)** | |
| African Caribbean in Barbados | ACB |
| African Ancestry in SW USA | ASW |
| Yoruba in Ibadan, Nigeria | YRI |
| Esan in Nigeria | ESN |
| Mende in Sierra Leone | MSL |
| Gambian in Western Division – Mandinka | GWD |
| Luhya in Webuye, Kenya | LWK |
| Bengali in Bangladesh | BEB |
| Sri Lankan Tamil in the UK | STU |
| Punjabi in Lahore, Pakistan | PJL |
| Indian Telugu in the U.K. | ITU |
| Gujarati Indians in Houston, Texas, USA | GIH |
| Chinese Dai in Xishuangbanna, China | CDX |
| Han Chinese in Beijing, China | CHB |
| Han Chinese South | CHS |
| Japanese in Tokyo, Japan | JPT |
| Kinh in Ho Chi Minh City, Vietnam | KHV |
| Utah residents (CEPH) with Northern and Western European ancestry | CEU |
| Toscani in Italia | TSI |
| Finnish in Finland | FIN |
| British From England and Scotland | GBR |
| Iberian Populations in Spain | IBS |
| Colombian in Medellín, Colombia | CLM |
| Puerto Rican in Puerto Rico | PUR |
| Peruvian in Lima Peru | PEL |

| | |
|---|---|
| Mexican Ancestry in Los Angeles CA USA | MXL |
| **Human Genome Diversity Project (HGDP)** | |
| Mbuti | |
| Biaka | |
| Mandenka | |
| Yoruba | |
| San | |
| Bantu South Africa | |
| Bantu Kenya | |
| Colombian | |
| Surui | |
| Maya | |
| Karitiana | |
| Pima | |
| Brahui | |
| Balochi | |
| Hazara | |
| Makrani | |
| Sindhi | |
| Pathan | |
| Kalash | |
| Burusho | |
| Uygur | |
| Bougainville | |
| PapuanSepik | |
| PapuanHighlands | |
| Druze | |
| Bedouin | |

| |
|---|
| Palestinian |
| Mozabite |
| French |
| Sardinian |
| Orcadian |
| Russian |
| BergamoItalian |
| Tuscan |
| Basque |
| Adygei |
| Cambodian |
| Japanese |
| Han |
| Yakut |
| Tujia |
| Yi |
| Miao |
| Oroqen |
| Daur |
| Mongolian |
| Hezhen |
| Xibo |
| NorthernHan |
| Dai |
| Lahu |
| She |
| Naxi |
| Tu |

| PAGE- WHI |
|---|
| Native American |
| **PAGE -STANFORD GRP** |
| Quechuan/Aymaran ancestry from Puno, Peru |

**Supplemental Table 2:** Population descriptors and abbreviations for samples ascertained from reference panels used for global ancestry inference.

| Reference Dataset | Abbreviation | European Local Ancestry | African Local Ancestry | Native American Local Ancestry |
|---|---|---|---|---|
| Thousand Genomes Project | 1KGP | Utah residents (CEPH) with Northern and Western European ancestry (CEU):**121** | Yoruba in Ibadan, Nigeria (YRI): **121** | Mexican Ancestry in Los Angeles, California (MXL): **2** |
| Thousand Genomes Project | 1KGP | | | Peruvian in Lima, Peru: **20** |
| Human Genome Diversity Project | HGDP | | | AMERICA: **45** |
| Population Architecture using Genomics and Epidemiology | PAGE WHI | | | PAGE WHI: **7** |
| Population Architecture using Genomics and Epidemiology | PAGE SGRP | | | PAGE SGRP: **68** |
| Total | | **120** | **121** | **142** |

**Supplemental Table 3:** Description of reference panel names, population labels and sample sizes selected to train GNOMIX local ancestry inference model.

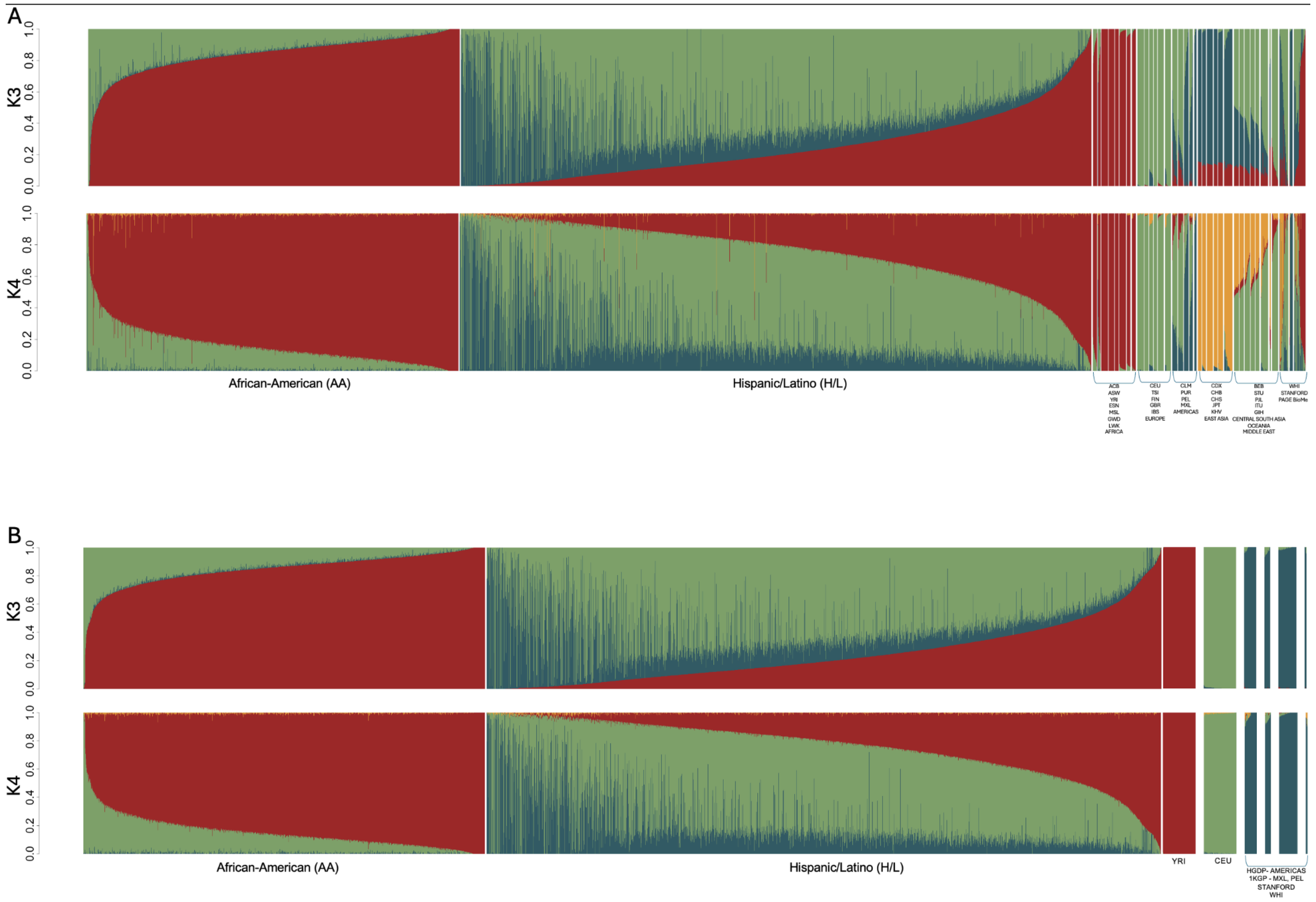

**Supplemental Figure 3**: Admixture plots illustrating K3 and K4 global ancestry components A) pre-quality control and B) post-quality control AA and HL Bio*Me* cohorts and reference panels

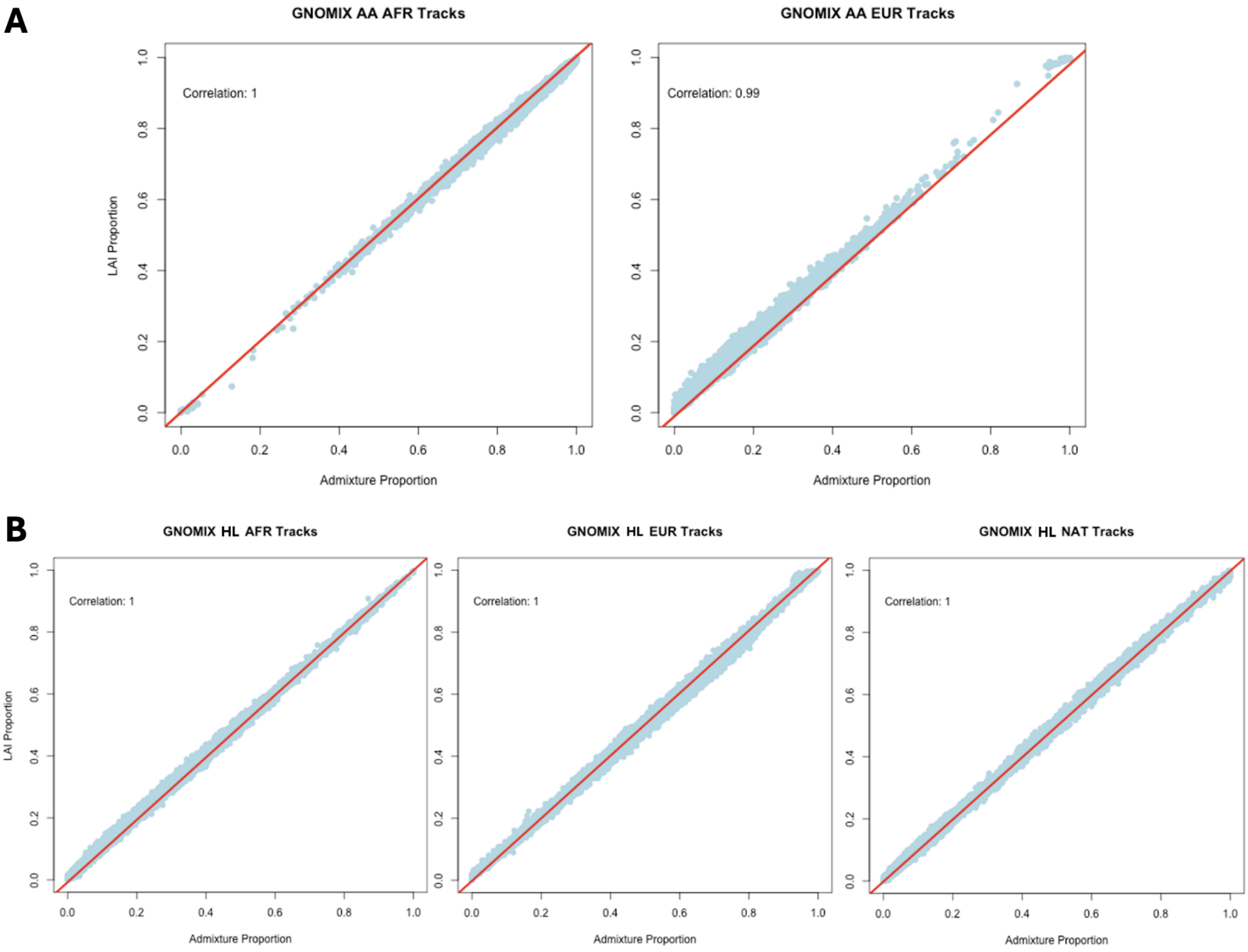

**Supplemental Figure 4:** Correlation between global ancestry admixture component proportion and the sum of physical distances of GNOMIX local ancestry calls for the corresponding ancestry component as a proportion of the sum of all local ancestry calls (LAI proportion) at: A) K3 admixture component in AA participants. B) K4 admixture component  in HL participants.

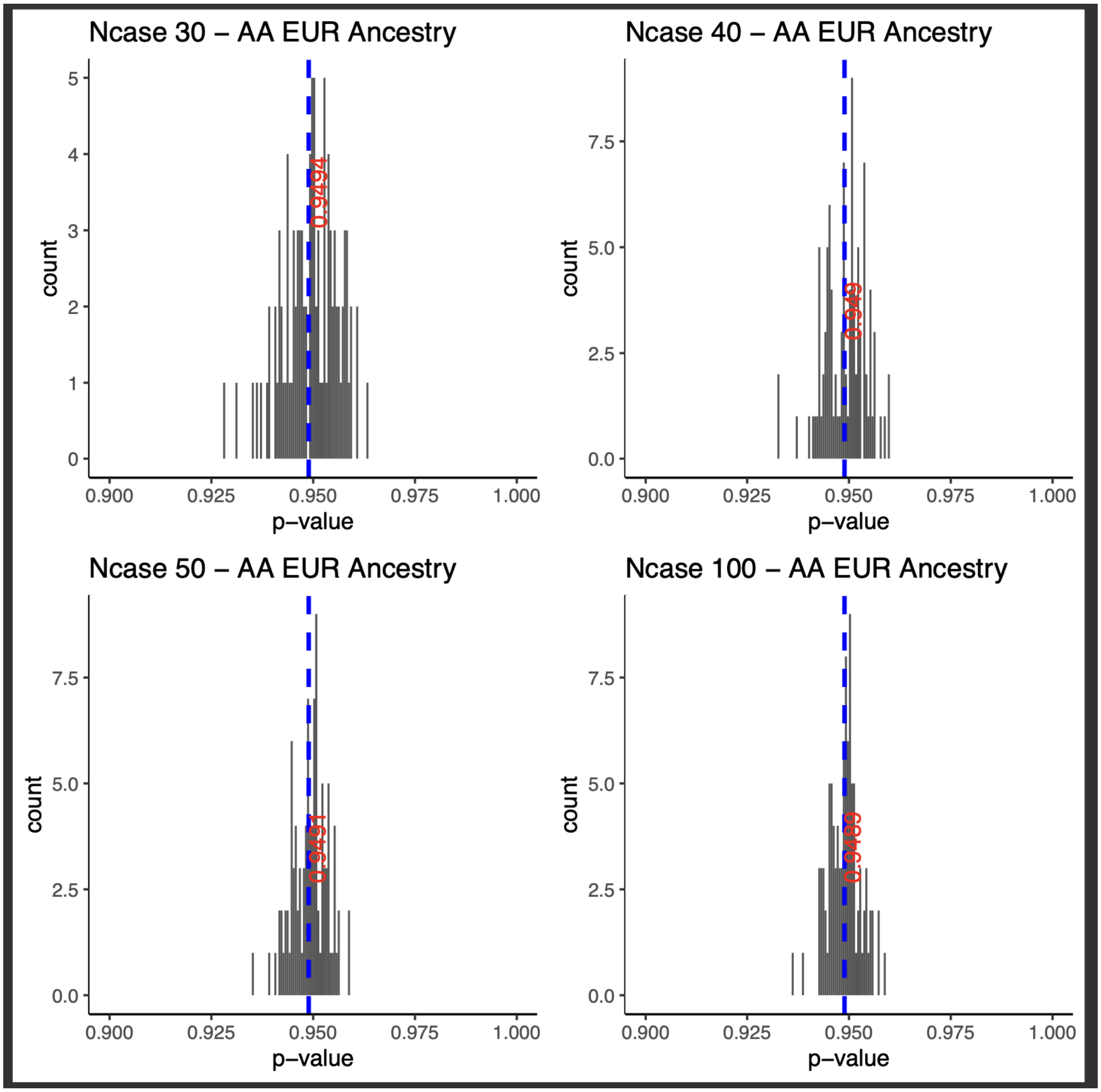

**Supplemental Figure 5:** Histogram plots showing the distribution of p-values at the 95th percentile per admixture mapping permutation in the African American (AA) cohort. Case-control assignments were randomized across four different thresholds of case counts: 30, 40, 50, and 100. Each threshold underwent 100 permutations. The mean p-value is represented by a blue dashed line, with the numerical mean value indicated in red

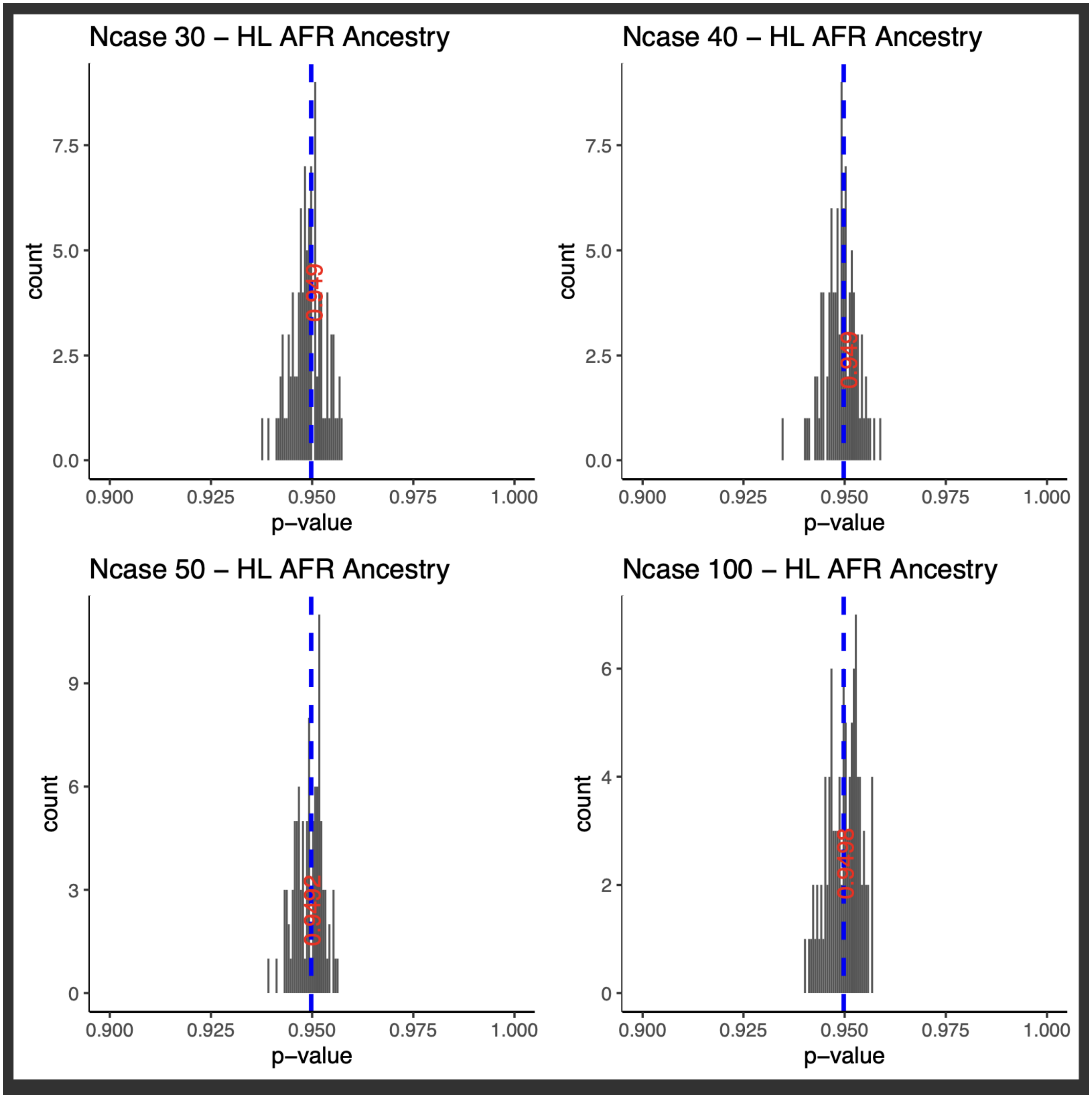

**Supplemental Figure 6:** Histogram plots showing the distribution of p-values at the 95th percentile per admixture mapping permutation in the Hispanic Latino (HL) cohort using African (AFR) local ancestry haplotype calls. Case-control assignments were randomized across four different thresholds of case counts: 30, 40, 50, and 100. Each threshold underwent 100 permutations. The mean p-value is represented by a blue dashed line, with the numerical mean value indicated in red.

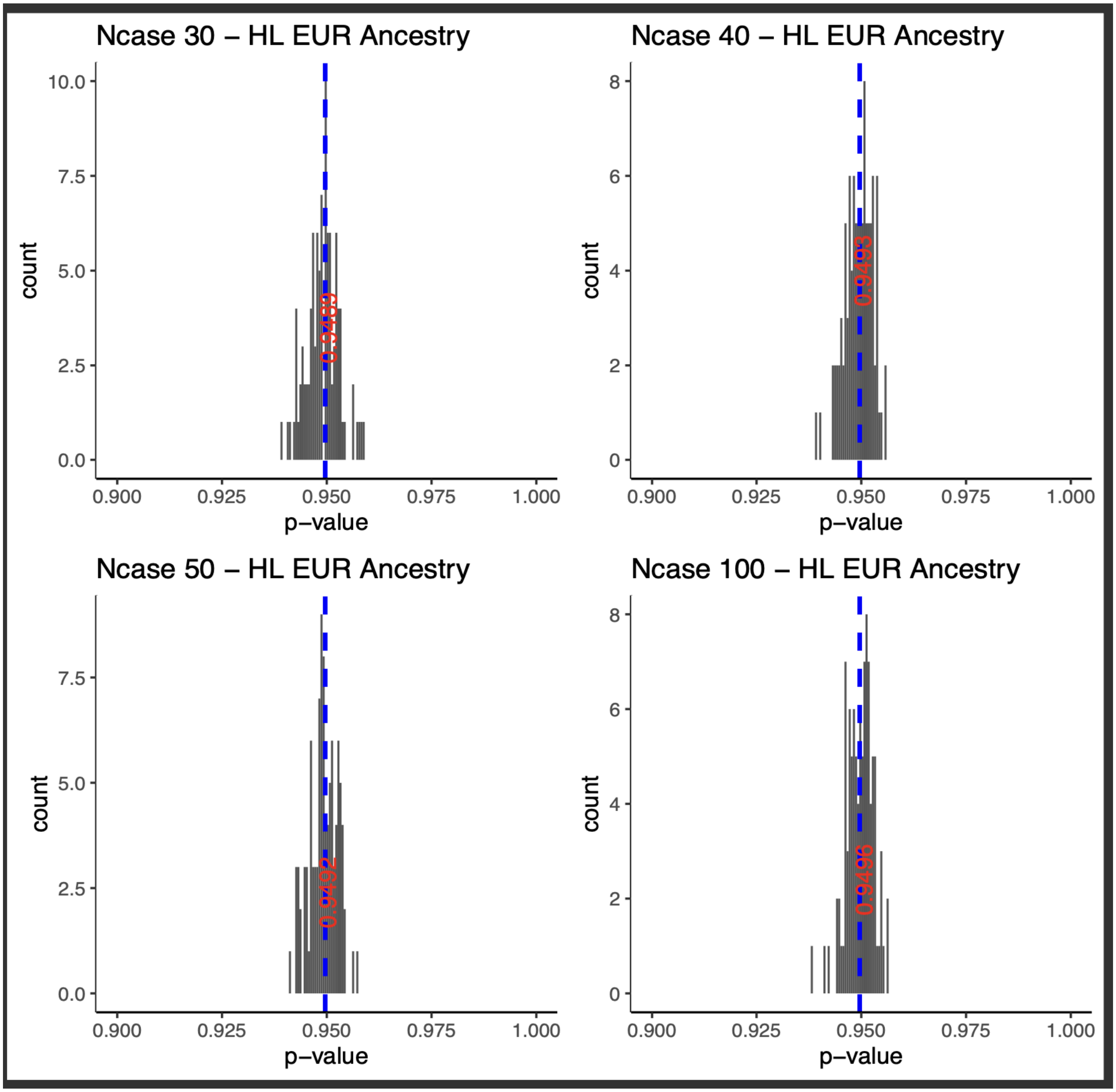

**Supplemental Figure 7:** Histogram plots showing the distribution of p-values at the 95th percentile per admixture mapping permutation in the HL cohort using European (EUR) local ancestry haplotype calls. Case-control assignments were randomized across four different thresholds of case counts: 30, 40, 50, and 100. Each threshold underwent 100 permutations. The mean p-value is represented by a blue dashed line, with the numerical mean value indicated in red

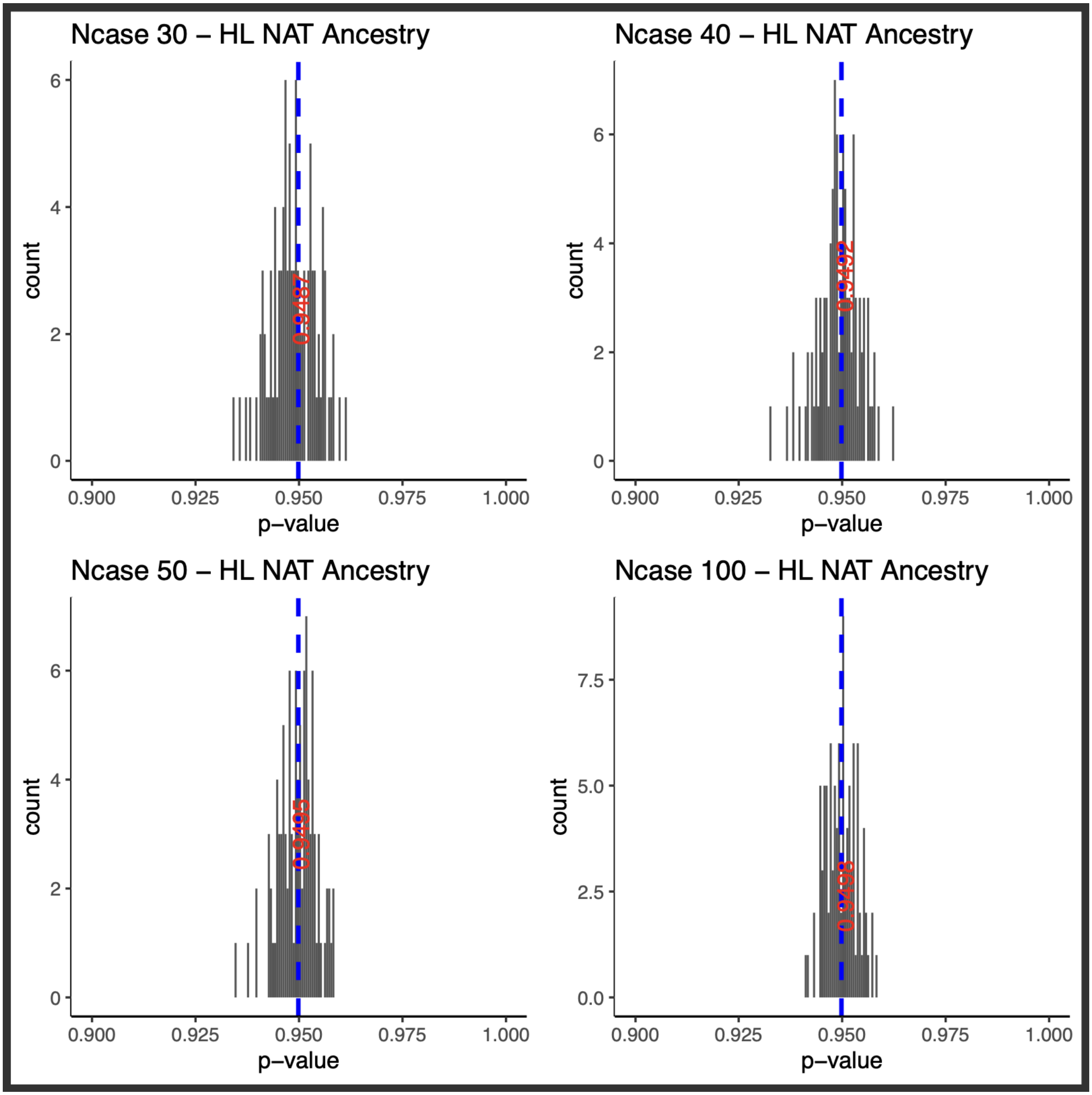

**Supplemental Figure 8**: Histogram plots showing the distribution of p-values at the 95th percentile per admixture mapping permutation in the Hispanic Latino (HL) cohort using Native American (NAT) local ancestry haplotype calls. Case-control assignments were randomized across four different thresholds of case counts: 30, 40, 50, and 100. Each threshold underwent 100 permutations. The mean p-value is represented by a blue dashed line, with the numerical mean value indicated in red.

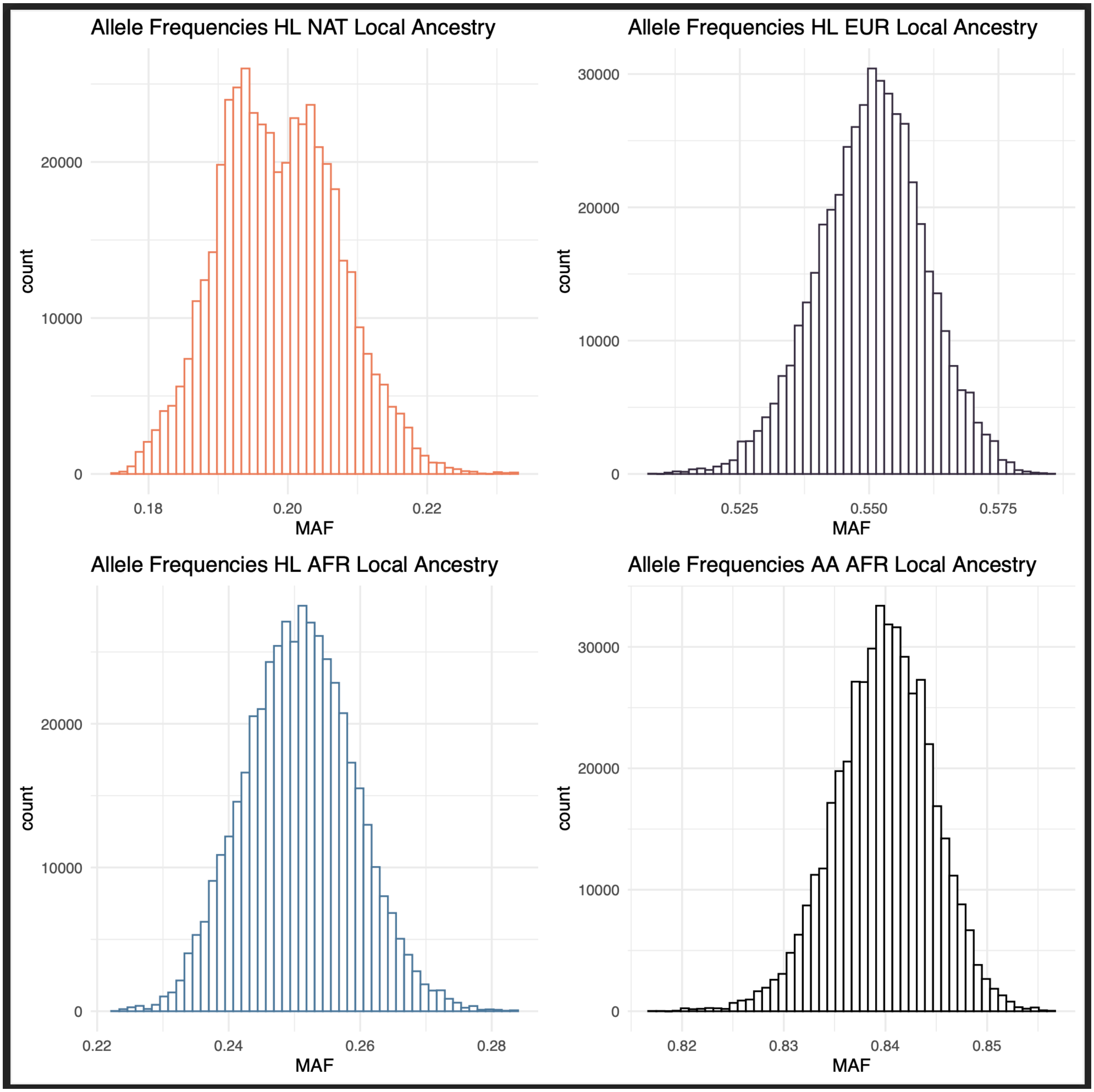

**Supplemental Figure 9:** Allele frequency distribution plots of local ancestry halplotypes tested in admixture mapping. The distribution of the allele frequency, which is essentially the frequency of the local ancestry haplotype is centered around the average proportion of each ancestry component in HL. In AA the allele frequency of African local ancestry haplotypes are plotted.

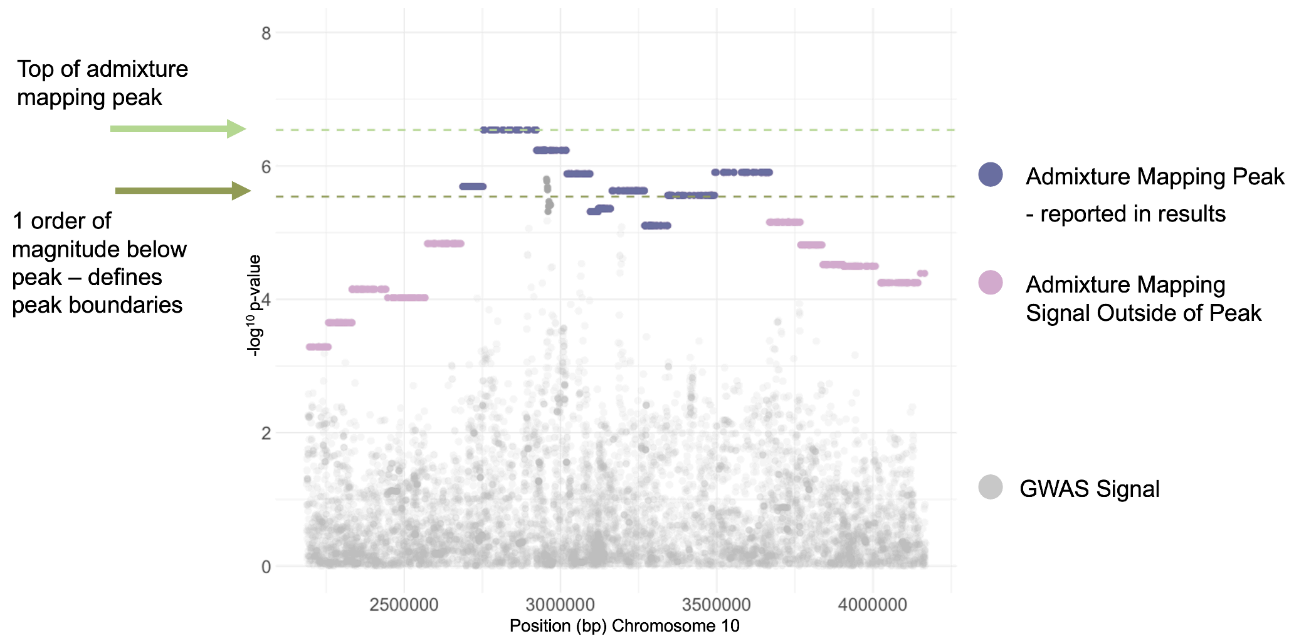

**Supplemental Figure 10**: Example of admixture mapping peak definition. Zoom in of admixture mapping peak for the top association in the AA cohort: phecode 288.2 “Elevated White Blood Cells” $AA^{EUR}$ chr1q23.1. Variant level GWAS signal is depicted by gray dots, admixture mapping peak association signal shown by purple dots. Admixture mapping signal outside of peak boundaries shown by pink dots. Top of admixture mapping signal is represented by dark green horizontal line. P-value one order of magnitude below the peak is depicted by light green horizontal line and is used to define peak boundaries.

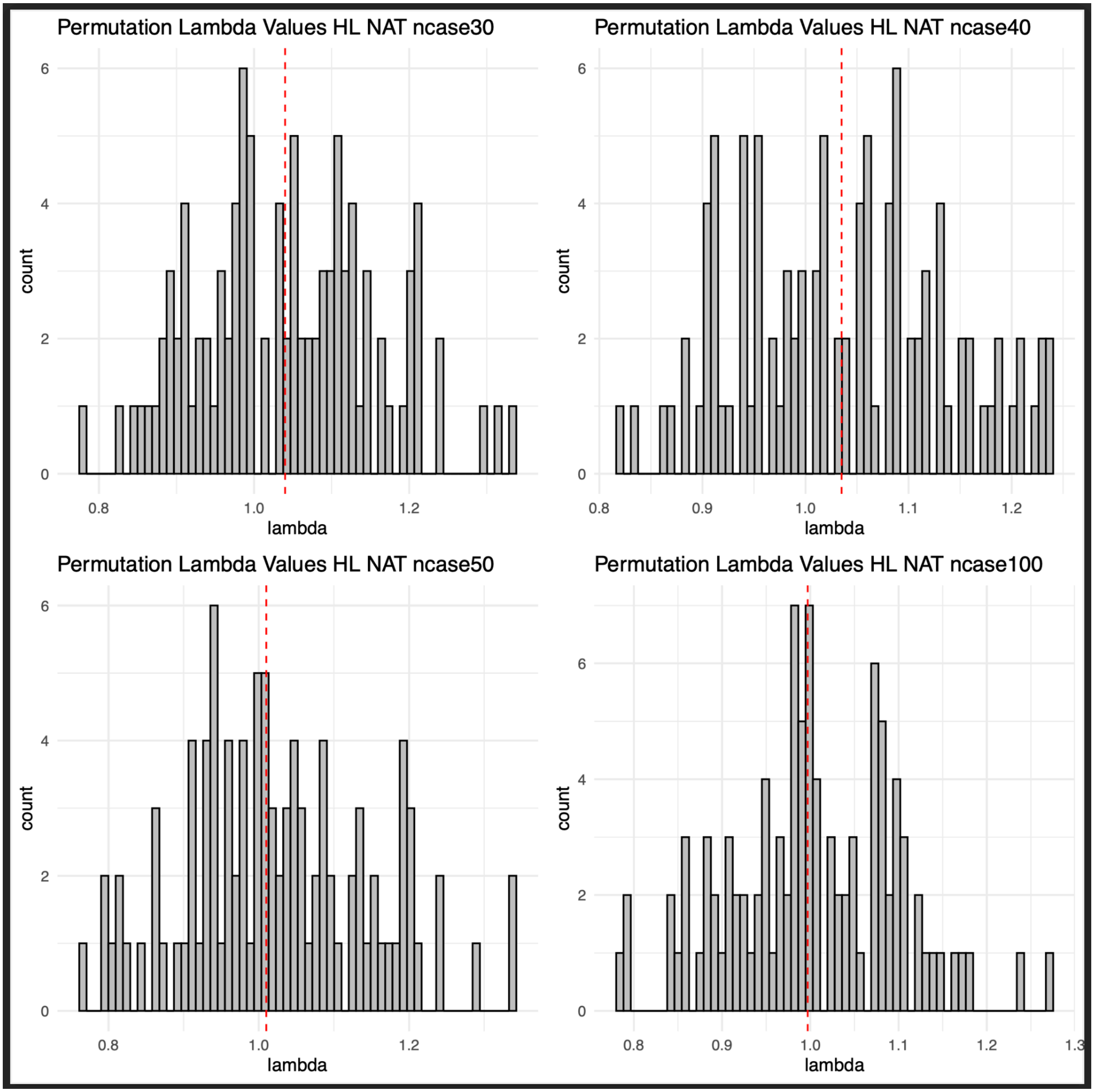

**Supplemental Figure 11:** Histogram plots showing the distribution of lambda values per admixture mapping permutation in the HL cohort using Native American (NAT) local ancestry haplotype calls. Case-control assignments were randomized across four different thresholds of case counts: 30, 40, 50, and 100. Each threshold underwent 100 permutations. The median lambda value is represented by a red dashed line.

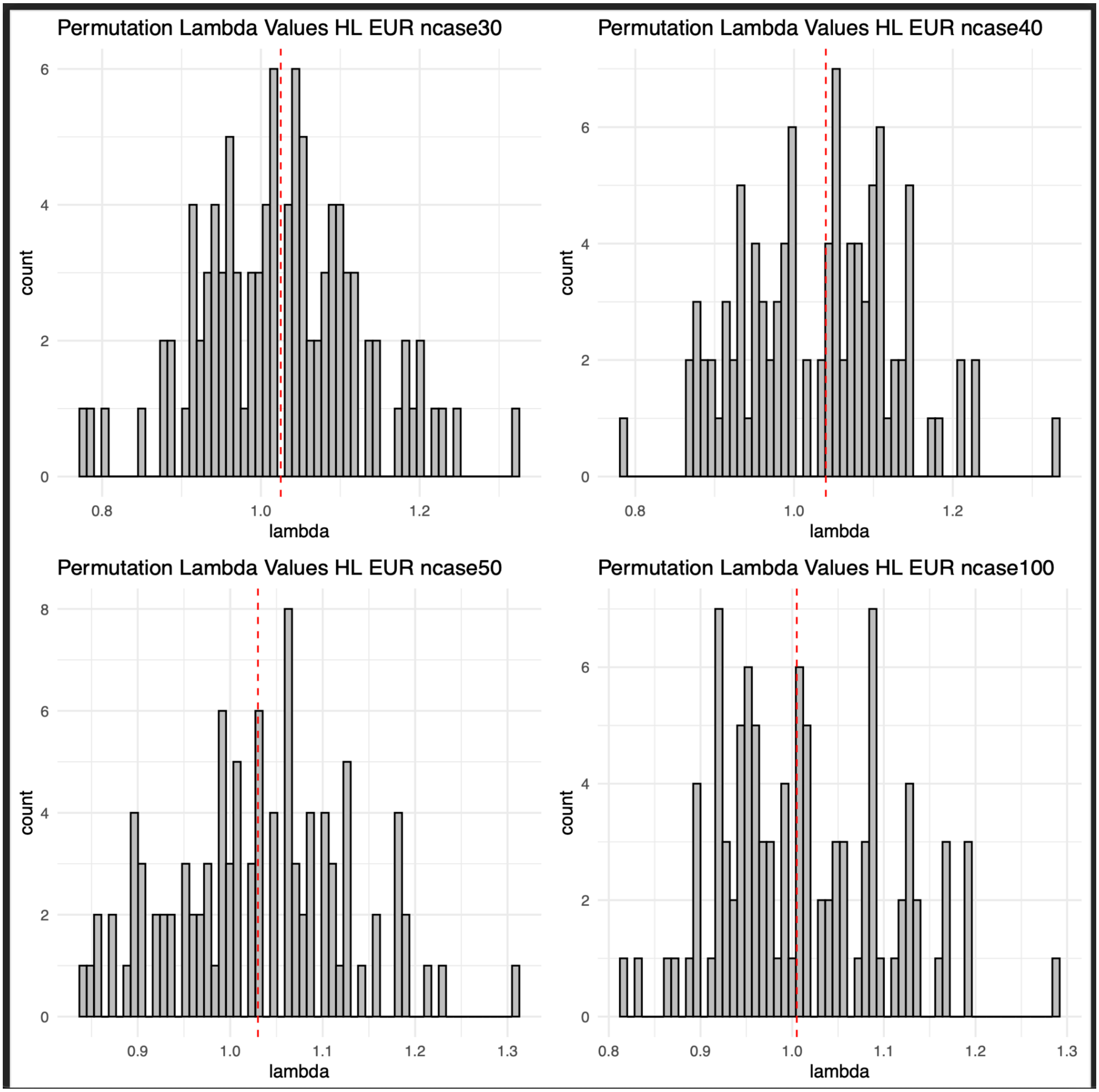

**Supplemental Figure 12:** Histogram plots showing the distribution of lambda values per admixture mapping permutation in the Hispanic Latino (HL) cohort using European (EUR) local ancestry haplotype calls. Case-control assignments were randomized across four different thresholds of case counts: 30, 40, 50, and 100. Each threshold underwent 100 permutations. The median lambda value is represented by a red dashed line.

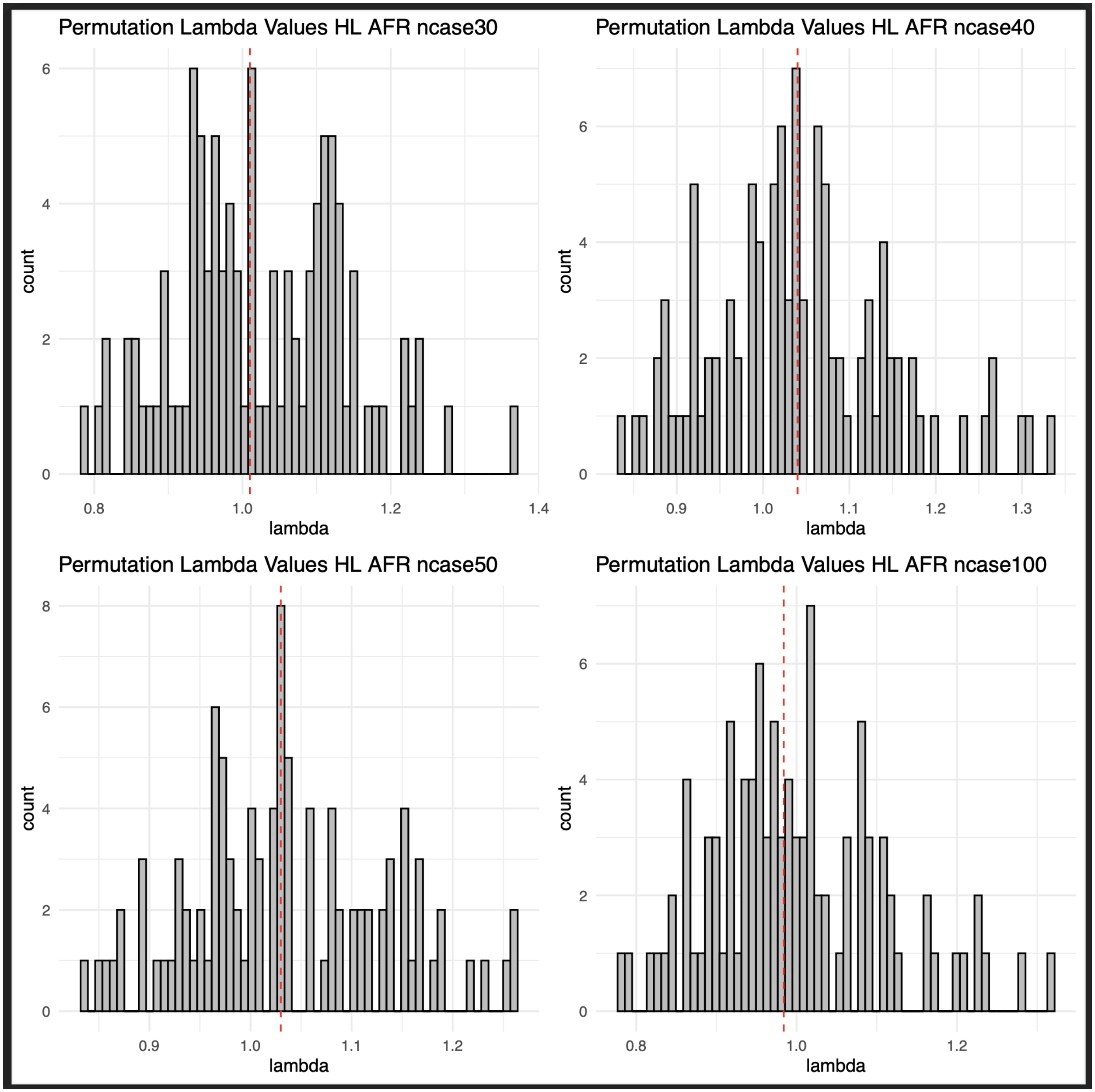

**Supplemental Figure 13:** Histogram plots showing the distribution of lambda values per admixture mapping permutation in the HL cohort using African (AFR) local ancestry haplotype calls. Case-control assignments were randomized across four different thresholds of case counts: 30, 40, 50, and 100. Each threshold underwent 100 permutations. The median lambda value is represented by a red dashed line.

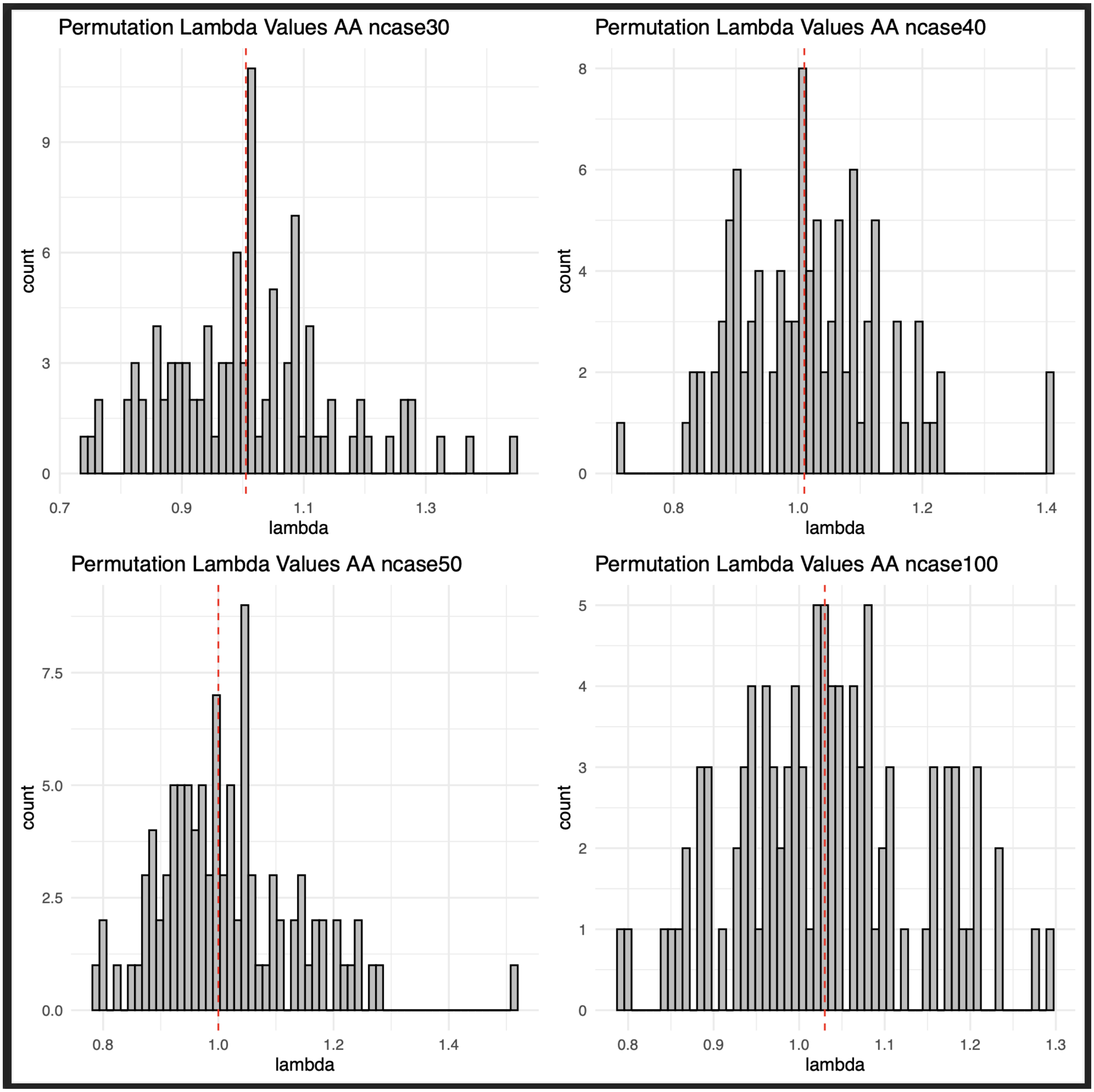

**Supplemental Figure 14:** Histogram plots showing the distribution of lambda values per admixture mapping permutation in the African American cohort. Case-control assignments were randomized across four different thresholds of case counts: 30, 40, 50, and 100. Each threshold underwent 100 permutations. The median lambda value is represented by a red dashed line.

| | permutations | | | GWS admixture PheWAS results | | |
|---|---|---|---|---|---|---|
| | median lambda | lower bound lambda | higher bound lambda | N associations removed | min lambda | max lambda |
| HL NAT | 1.04 | 0.69 | 1.39 | 0 | 0.91 | 1.23 |
| HL EUR | 1.03 | 0.72 | 1.33 | 0 | 0.86 | 1.20 |
| HL AFR | 1.01 | 0.67 | 1.35 | 1 | 0.81 | 1.38 |
| AA | 1.01 | 0.59 | 1.42 | 0 | 0.839 | 1.41 |

**Supplemental Table 4:** Summary of lambda value distributions from admixture mapping permutations used to establish thresholds for admixture mapping PheWAS results (grey). The lower and upper lambda cutoffs were defined as ±3 standard deviations from the median. The resulting phecodes removed from PheWAS results are summarized (blue). One association in the HL admixture PheWAS teseting AFR local ancestry tracks was excluded due to inflation (phecode 557.1, Celiac disease, lambda = 1.38).

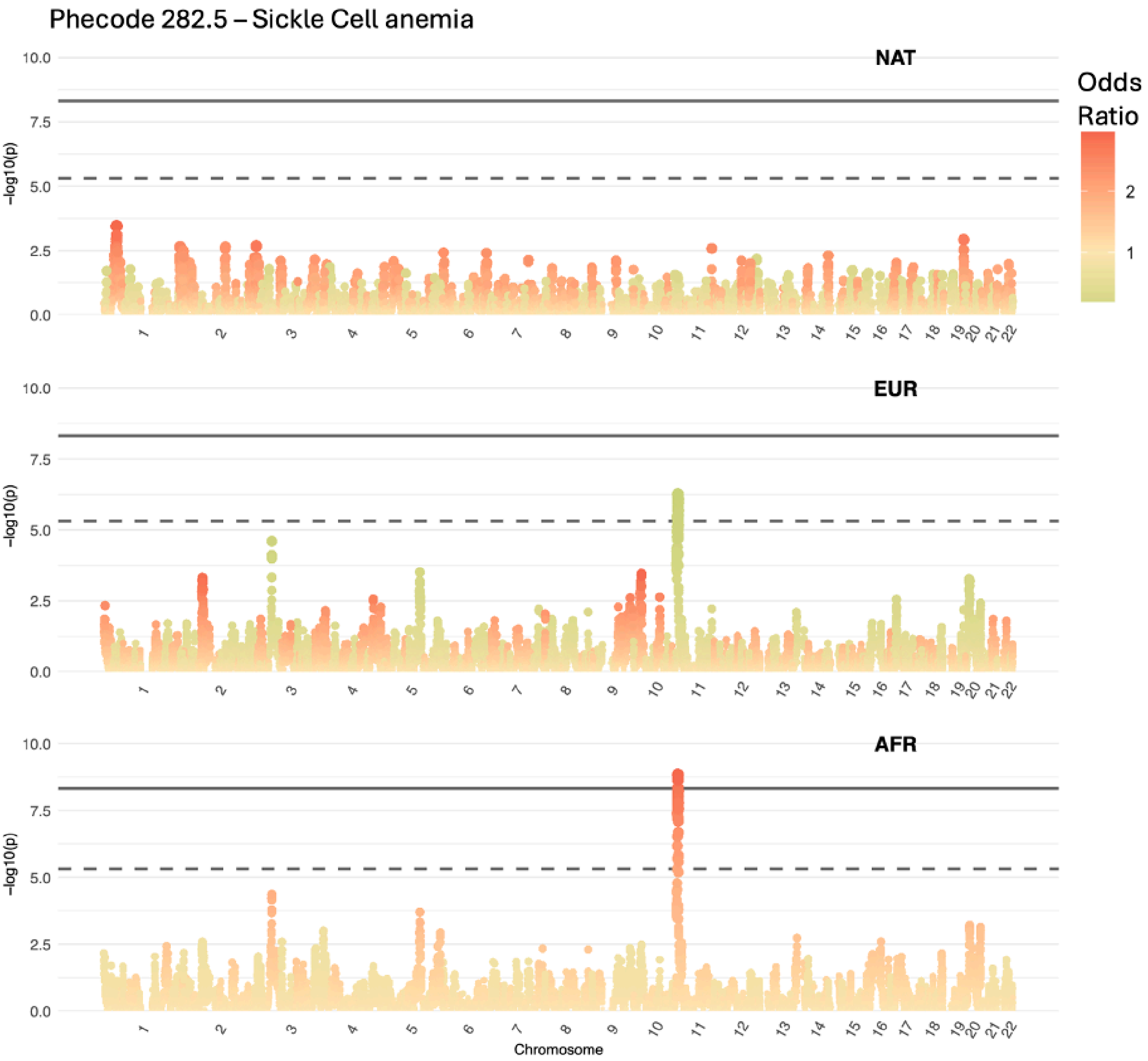

**Supplemental Figure 15**: Manhattan plots showing genome wide admixture mapping signal for phecode 282.5: "Sickle Cell Anaemia" in the Hispanic Latino cohort. Points are colored according to odds ratio. Each Manhattan plot corresponds to admixture mapping signal on a particular local ancestry background; Native American (NAT), European (EUR) and African (AFR). Genome wide association signal is observed on both the EUR and AFR backgrounds, with the AFR association signal being retained for reporting due to its risk odds ratio (OR).

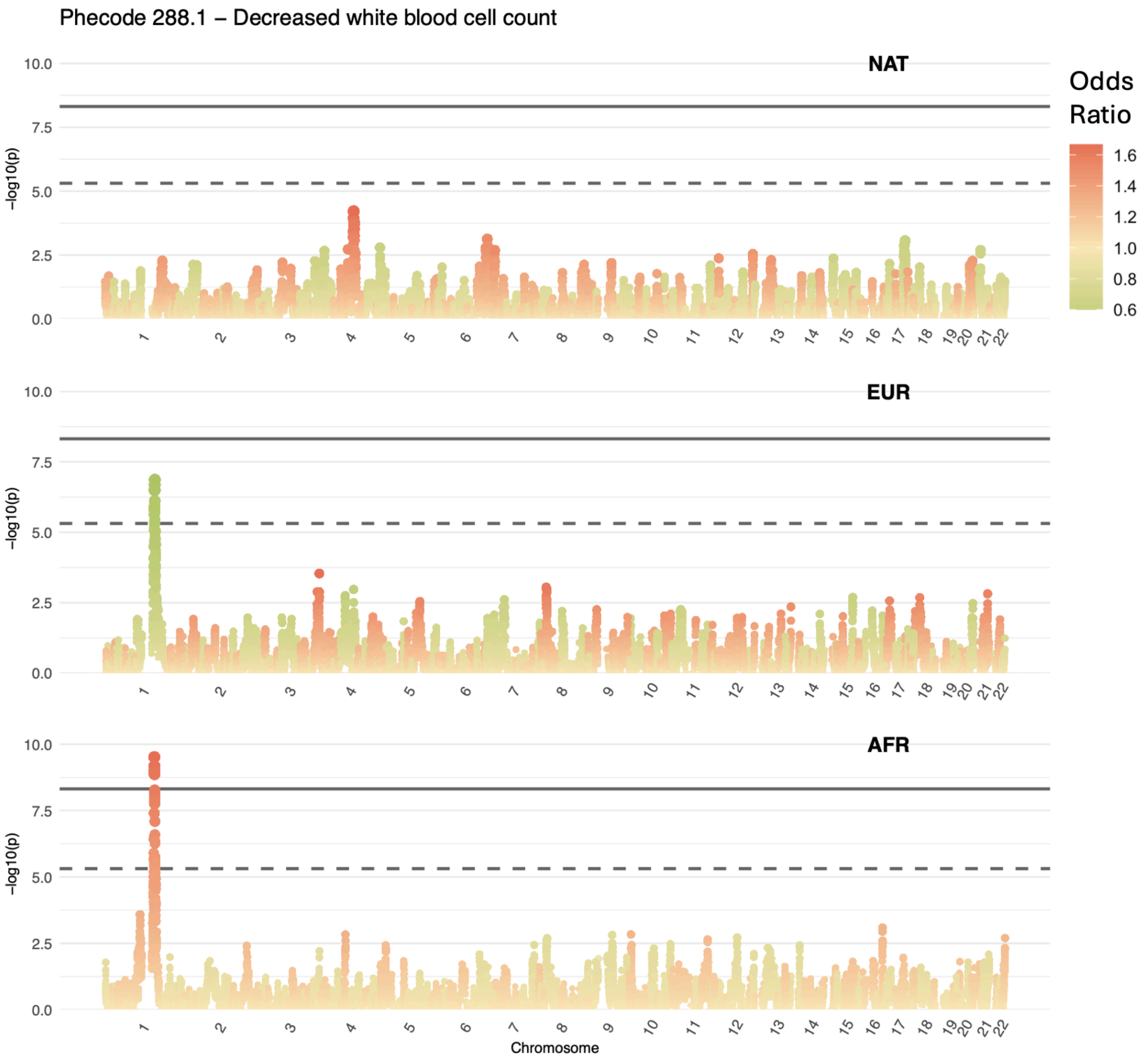

**Supplemental Figure 16:** Manhattan plots showing genome wide admixture mapping signal for phecode 288.1: “Decreased white blood cell count” in the HL cohort. Points are colored according to odds ratio. Each Manhattan plot corresponds to admixture mapping signal on a particular local ancestry background; Native American (NAT), European (EUR) and African (AFR). Genome wide association signal is observed on both the EUR and AFR backgrounds, with the AFR association signal being retained for reporting due to its risk odds ratio (OR).

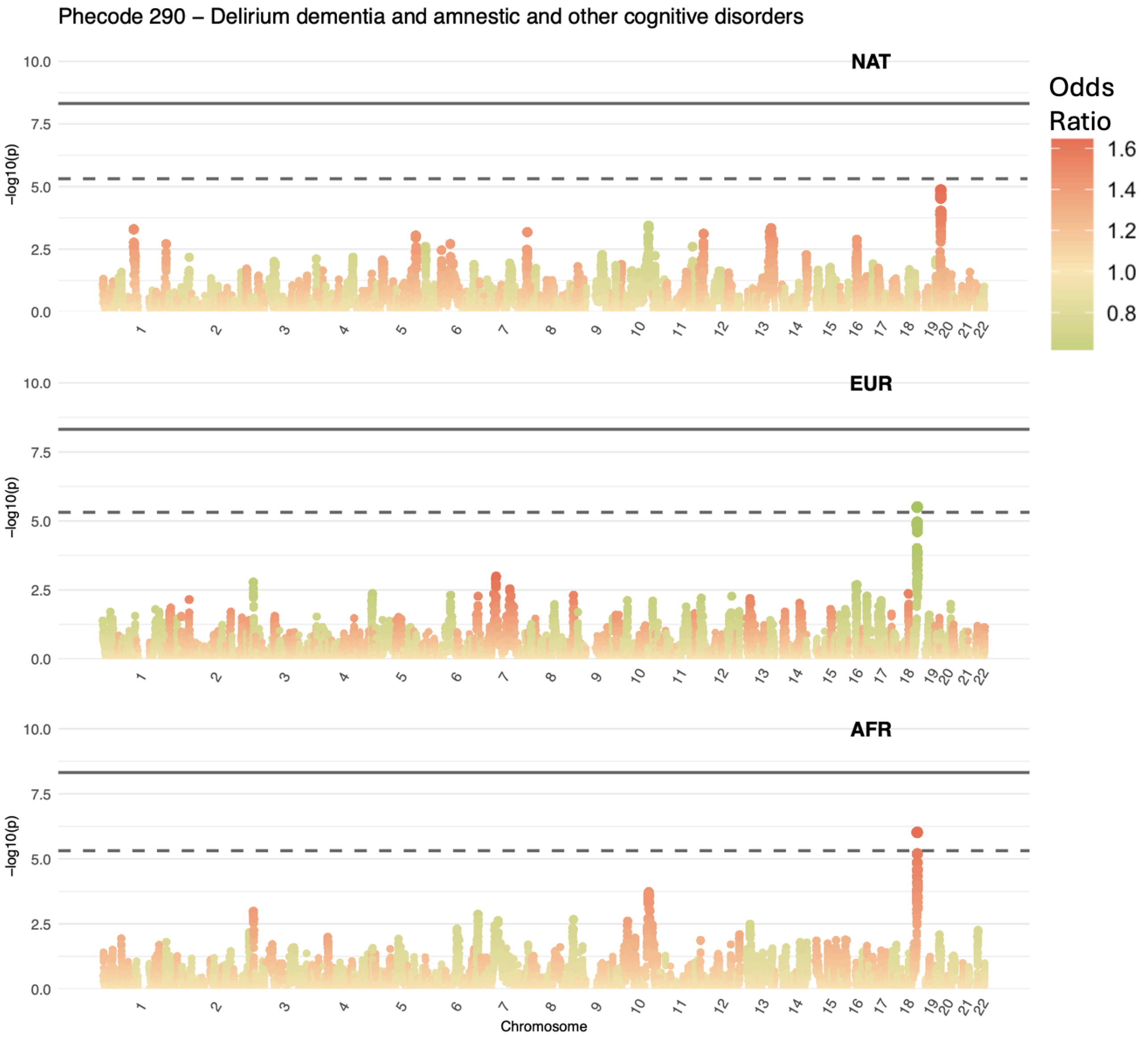

**Supplemental Figure 17**: Manhattan plots showing genome wide admixture mapping signal for phecode 290: "Delirium dementia and amnestic and other cognitive disorders" in the Hispanic Latino cohort. Points are colored according to odds ratio. Each Manhattan plot corresponds to admixture mapping signal on a particular local ancestry background; Native American (NAT), European (EUR) and African (AFR). Genome wide association signal is observed on both the EUR and AFR backgrounds, with the AFR association signal being retained for reporting due to its risk odds ratio (OR).

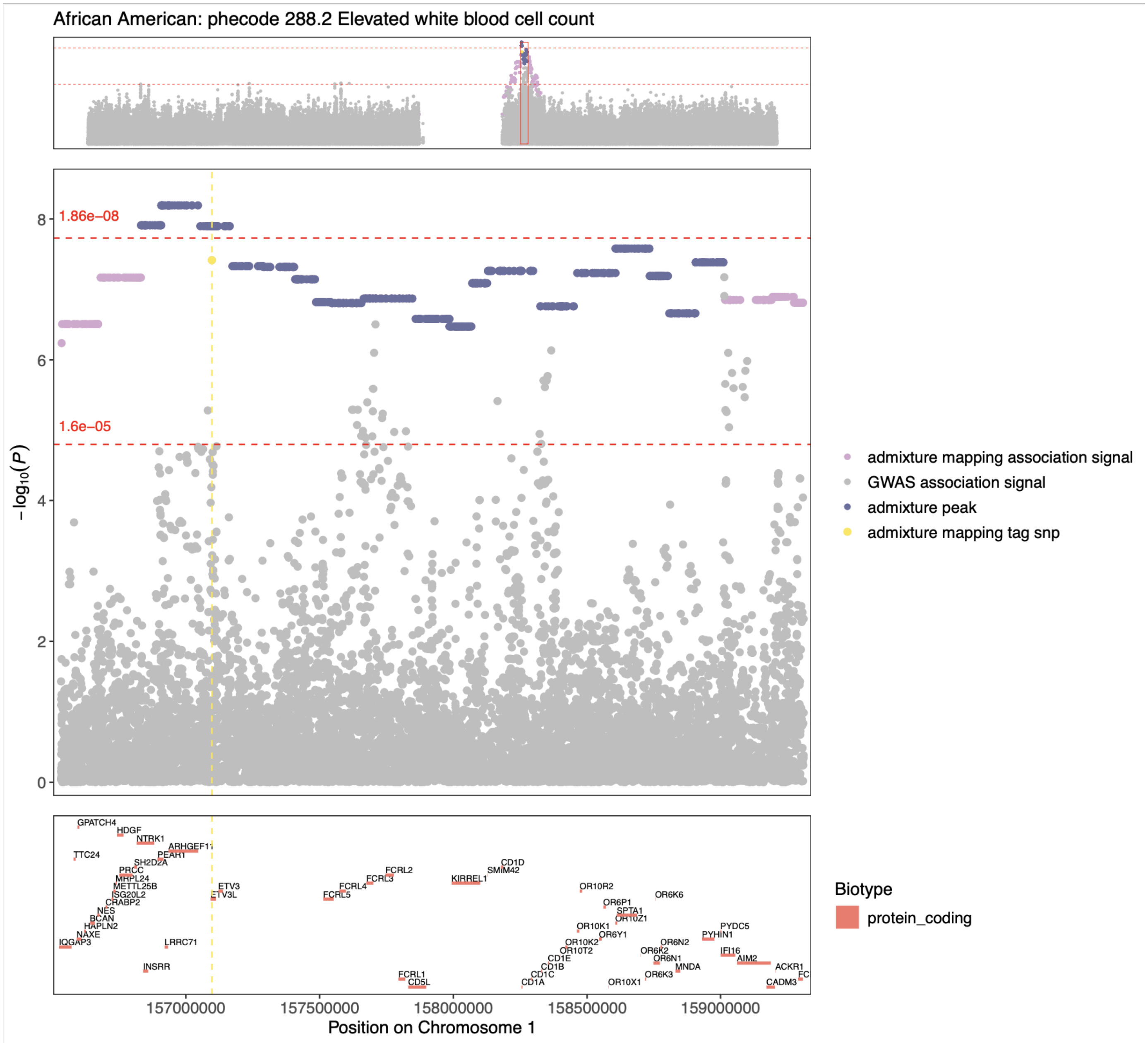

**Supplemental Figure 18** : Region plot illustrating admixture mapping and GWAS signals at a study-wide significant admixture mapping peak for phecode 288.2, "Elevated white blood cell count," in AA. Grey dots represent the GWAS signal, while purple dots indicate the admixture mapping peak. The top section shows a chromosome view, and the middle plot zooms in on the admixture peak. Red dashed horizontal lines represent genome wide and study wide significance thresholds. The yellow dashed line highlights the GWAS signal at the tag SNP identified using conditional association of the admixture mapping signal. Protein coding gene locations are depicted in the bottom section.

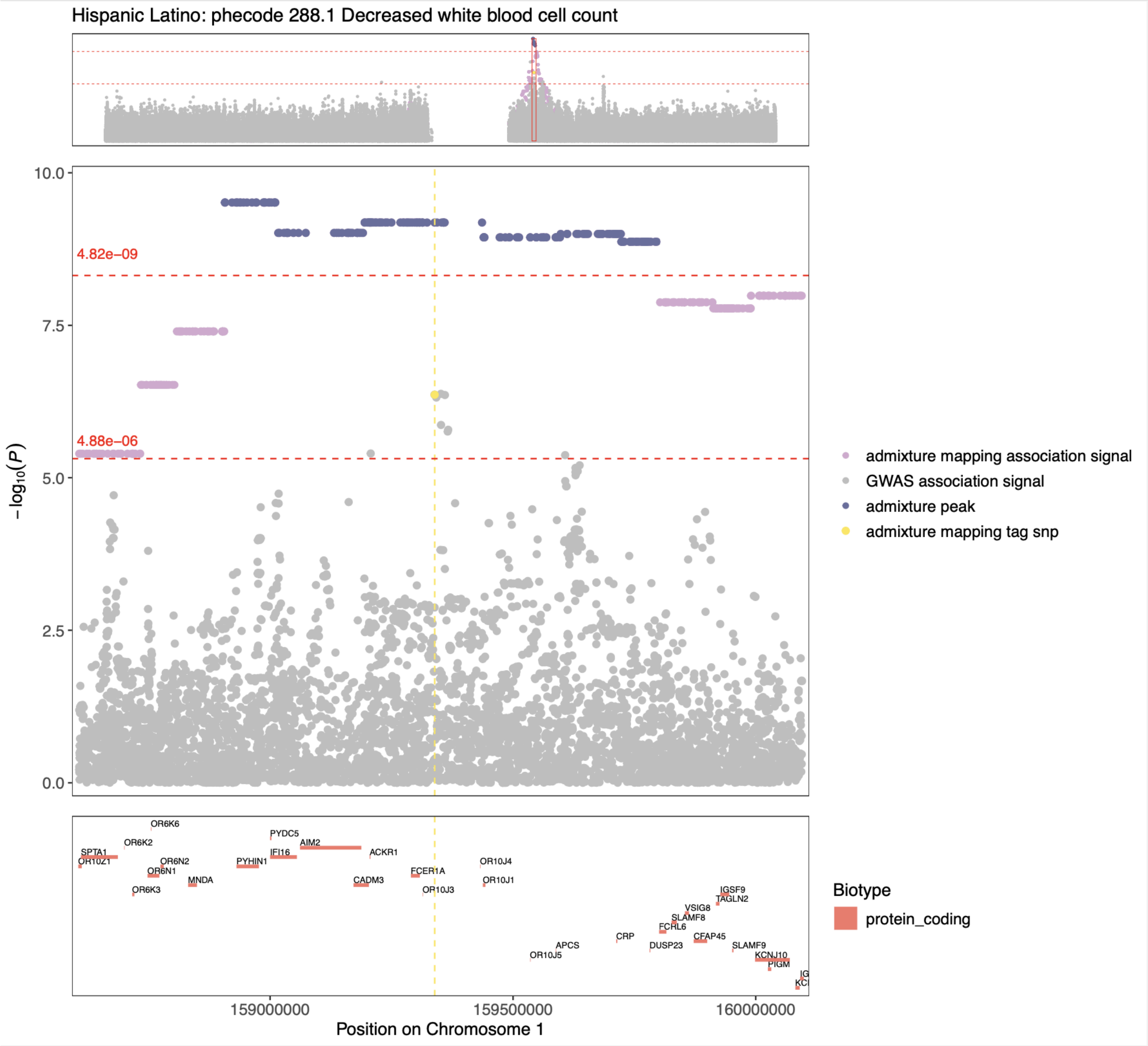

**Supplemental Figure 19:** Region plot illustrating admixture mapping and GWAS signals at a study-wide significant admixture mapping peak for phecode 288.1, "Decreased white blood cell count" in HL. Grey dots represent the GWAS signal, while purple dots indicate the admixture mapping peak. The top section shows a chromosome view, and the middle plot zooms in on the admixture peak. Red dashed horizontal lines represent genome-wide and study-wide significance thresholds. The yellow dashed line the GWAS signal at the tag SNP identified using conditional association of the admixture mapping signal. Protein-coding gene locations are depicted in the bottom section.

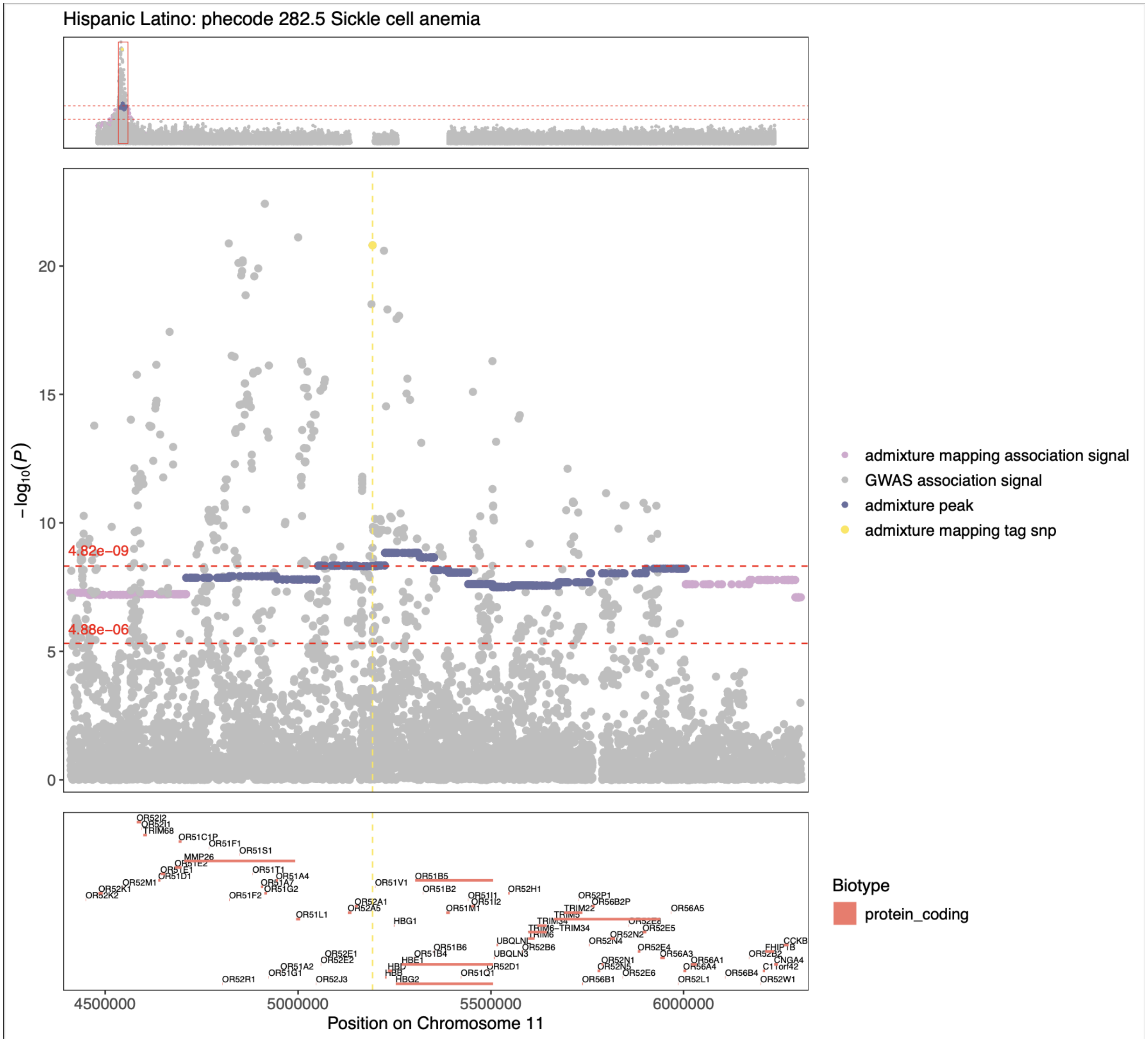

**Supplemental Figure 20**: Region plot illustrating admixture mapping and GWAS signals at a study-wide significant admixture mapping peak for phecode 282.5, "Sickle Cell Anemia" in Hispanic Latinos. Grey dots represent the GWAS signal, while purple dots indicate the admixture mapping peak. The top section shows a chromosome view, and the middle plot zooms in on the admixture peak. Red dashed horizontal lines represent genome-wide and study-wide significance thresholds. The yellow dashed line highlights the GWAS signal at the tag SNP identified using conditional association of the admixture mapping signal. Protein-coding gene locations are depicted in the bottom section.

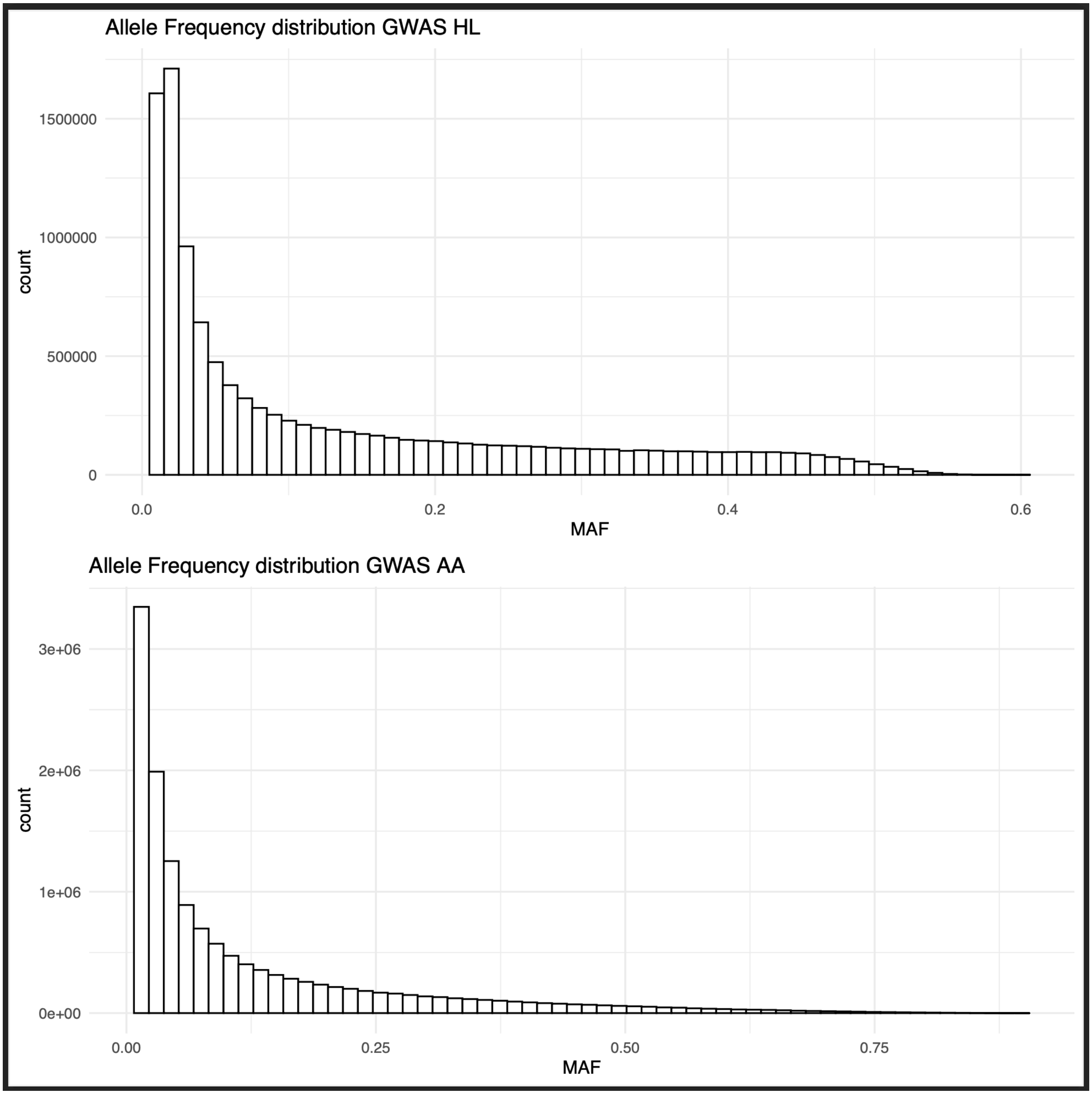

**Supplemental Figure 21**: Minor allele frequency spectrum of variants included in HL and AA GWAS post quality control.

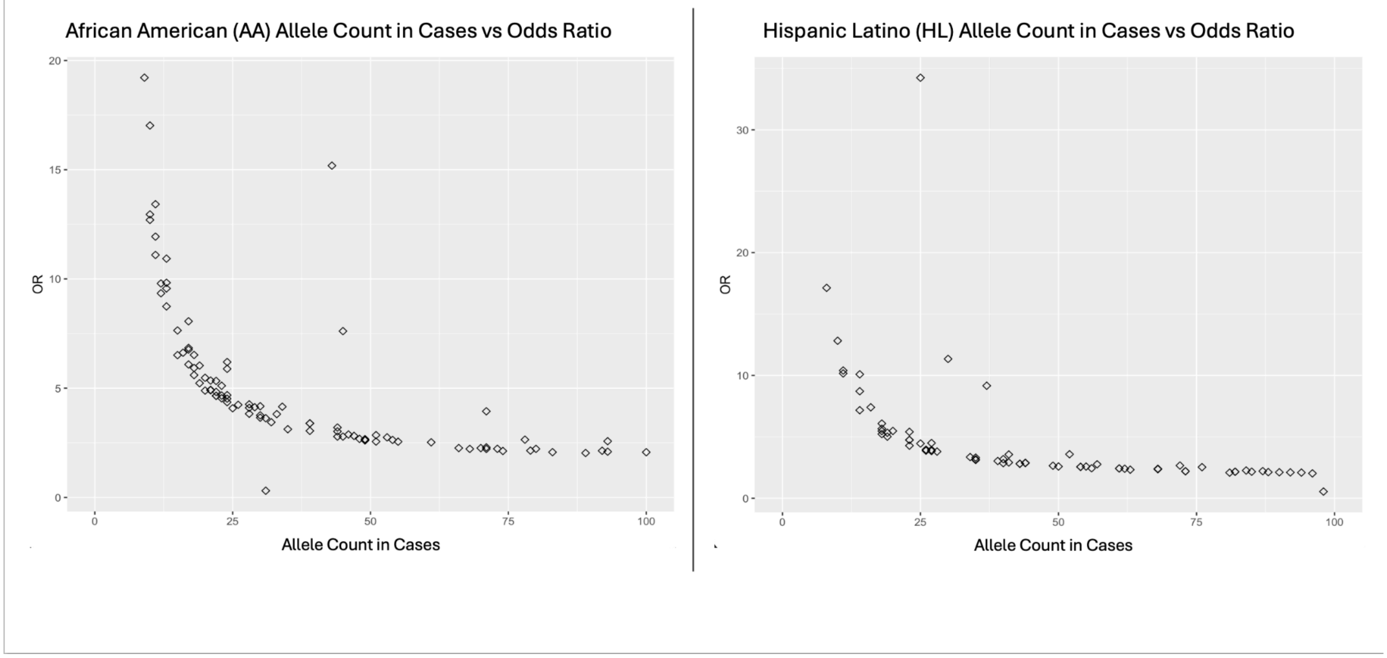

**Supplemental Figure 22:** Distribution of odds ratios (OR) across different values of allele counts in cases for GWAS associations in A) AA and B) HL cohorts. Each point represents the odds ratio of a single genome-wide significant association. The plots illustrate OR inflation as the allele count in cases goes below 40.

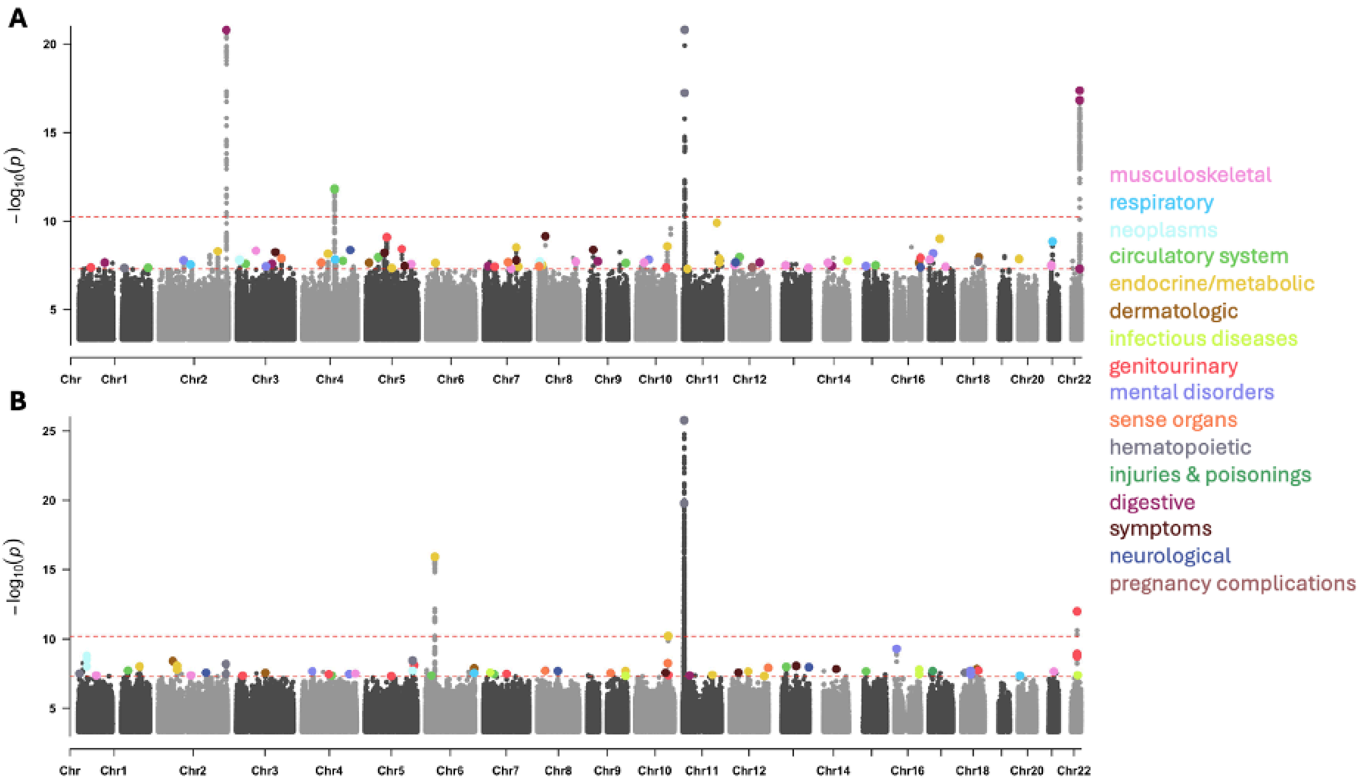

**Supplemental Figure 23:** Manhattan plots of GWAS phenome wide association study (PheWAS) results in Bio*Me*. Plotting associations with $p<5x10^{-4}$. A) HL cohort association results B) AA cohort association results. Dots represent association signal p-values with peaks passing genome wide significance colored by phecode categorical labels. Genome-wide and study-wide significance thresholds are indicated by the dashed red lines.

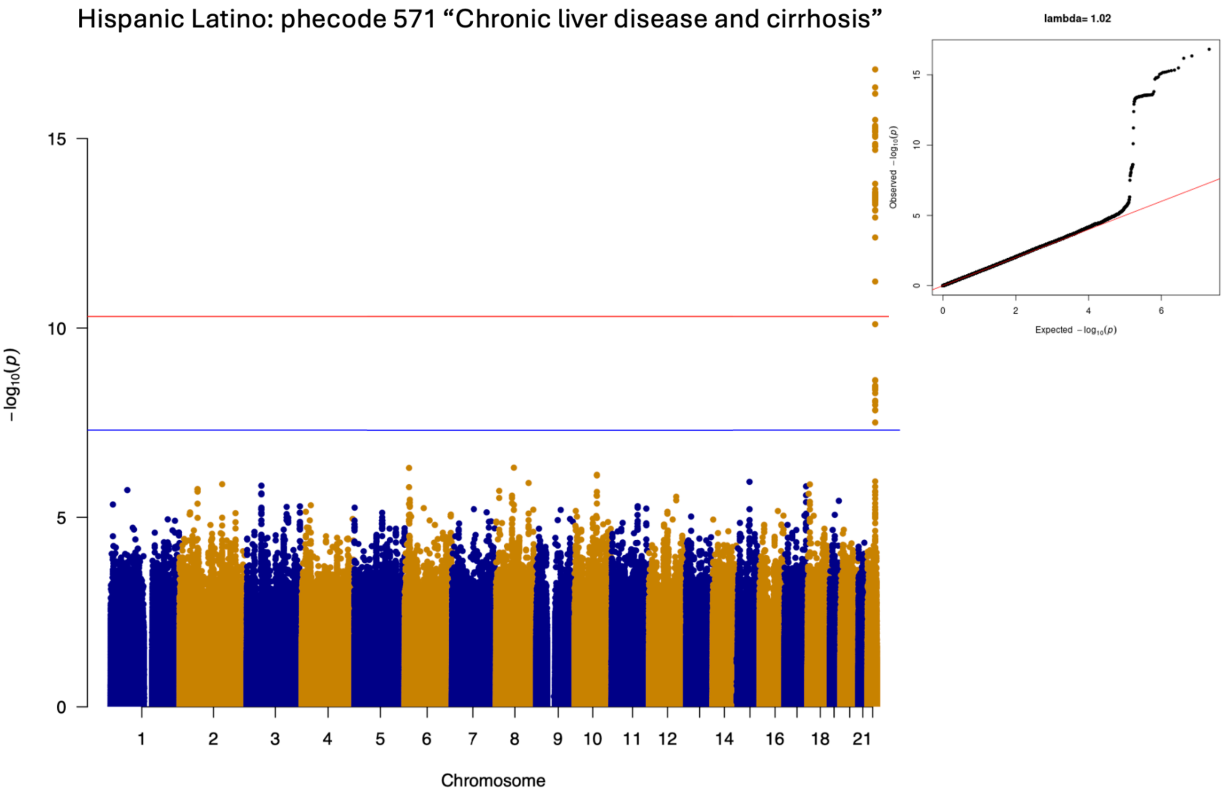

**Supplemental Figure 24:** Manhattan plot displaying GWAS association signal for phecode 571 "Chronic liver disease and cirrhosis" in Bio*Me* HL. Each point represents -log10 p-value at each tested variant colored by chromosome. The blue horizontal line indicates genome wide significance, while the red horizontal line indicates study wide significance. An inset shows a quantile-quantile (QQ) plot of observed versus expected p-value distributions, with the lambda inflation statistic reported above.

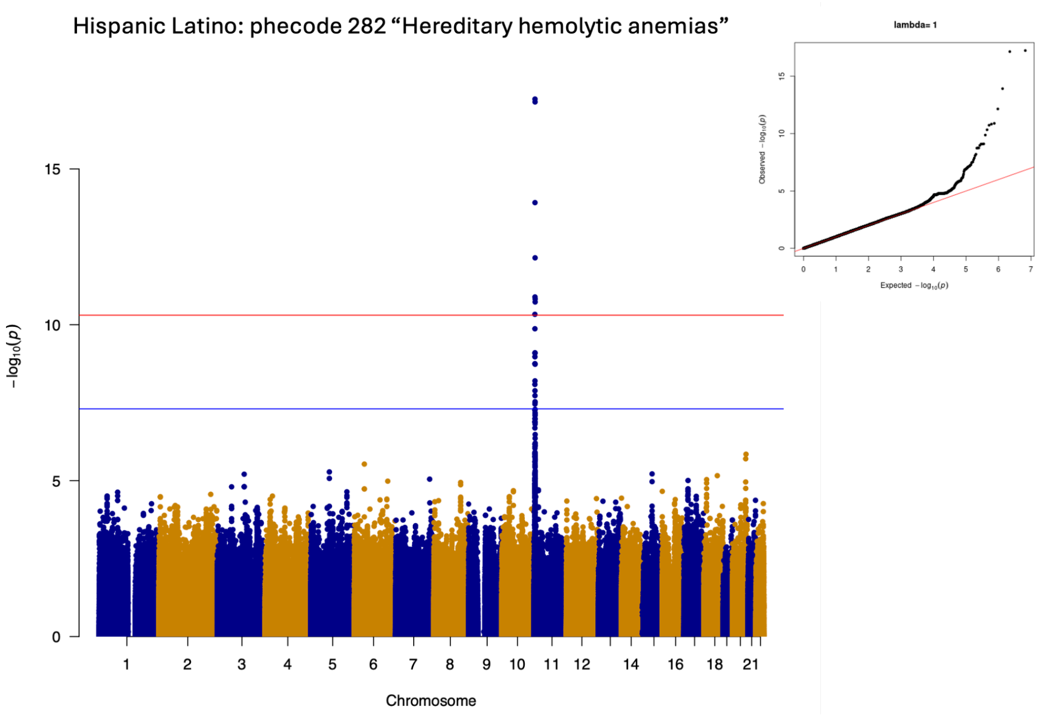

**Supplemental Figure 25**: Manhattan plot displaying GWAS association signal for phecode 427.2 "Heriditary hemolytic anemias" in Bio*Me* HL. Each point represents -log10 p-value at each tested variant colored by chromosome. The blue horizontal line indicates genome wide significance, while the red horizontal line indicates study wide significance. An inset shows a quantile-quantile (QQ) plot of observed versus expected p-value distributions, with the lambda inflation statistic reported above.

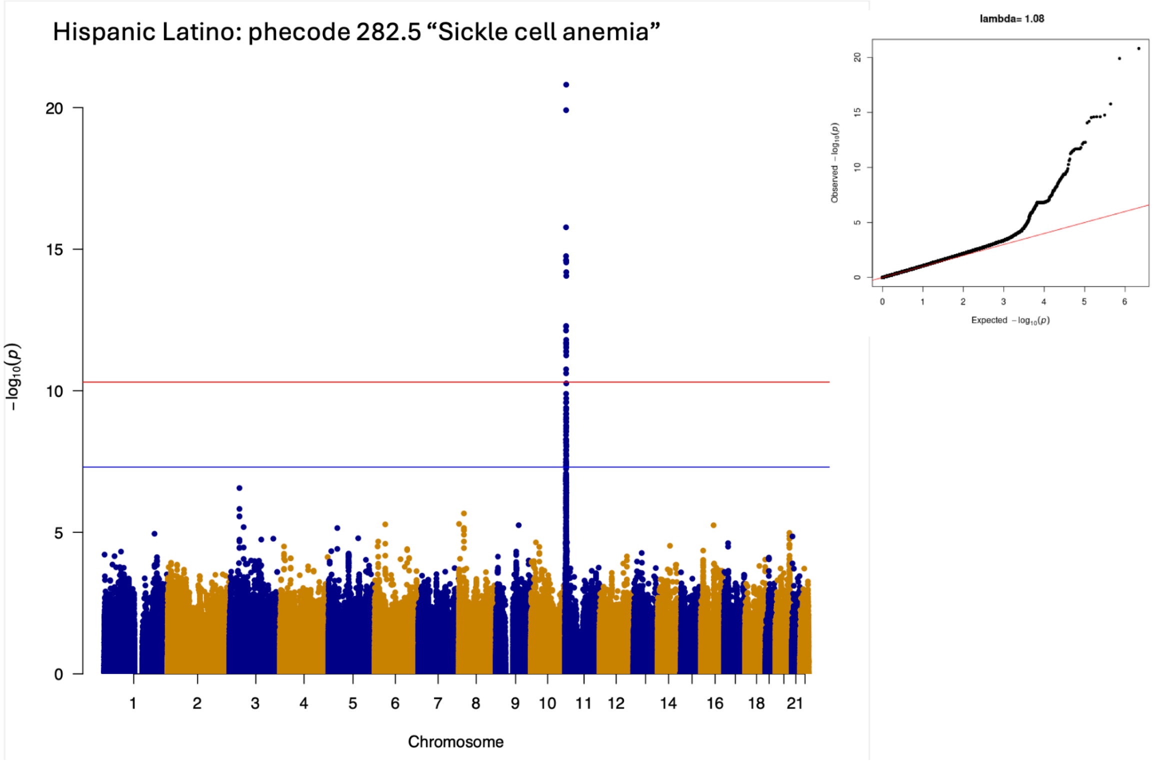

**Supplemental Figure 26:** Manhattan plot displaying GWAS association signal for phecode 282.5 "Sickle cell anemia" in Bio*Me* HL. Each point represents -log10 p-value at each tested variant colored by chromosome. The blue horizontal line indicates genome wide significance, while the red horizontal line indicates study wide significance. An inset shows a quantile-quantile (QQ) plot of observed versus expected p-value distributions, with the lambda inflation statistic reported above.

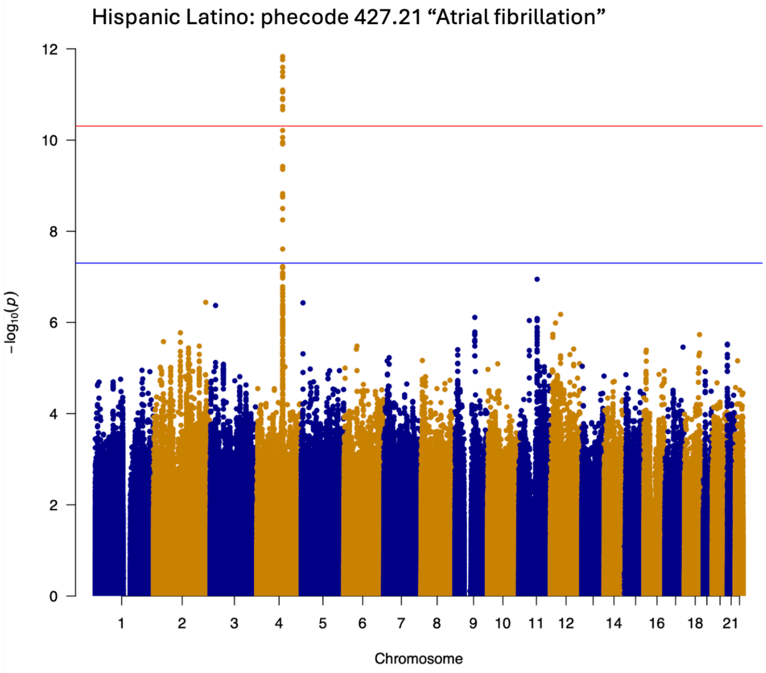

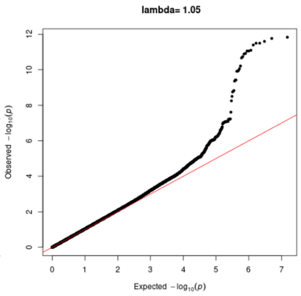

**Supplemental Figure 27**: Manhattan plot displaying GWAS association signal for phecode 427.21 "Atrial fibrillation" in Bio*Me* HL. Each point represents -log10 p-value at each tested variant colored by chromosome. The blue horizontal line indicates genome wide significance, while the red horizontal line indicates study wide significance. An inset shows a quantile-quantile (QQ) plot of observed versus expected p-value distributions, with the lambda inflation statistic reported above.

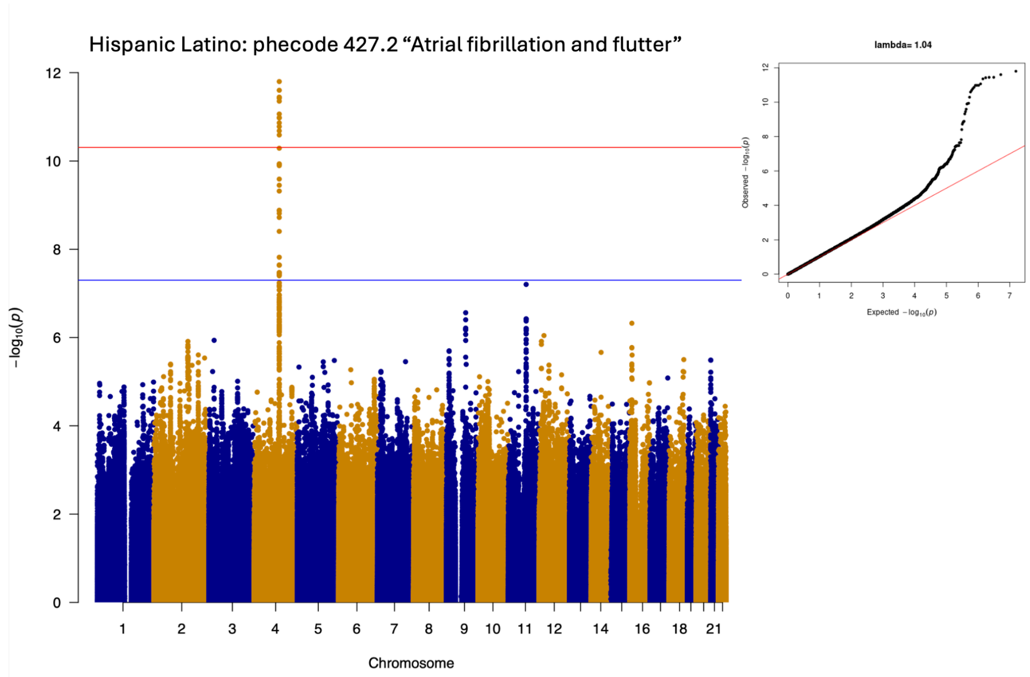

**Supplemental Figure 28**: Manhattan plot displaying GWAS association signal for phecode 427.2 "Atrial fibrillation and flutter" in Bio*Me* HL. Each point represents -log10 p-value at each tested variant colored by chromosome. The blue horizontal line indicates genome wide significance, while the red horizontal line indicates study wide significance. An inset shows a quantile-quantile (QQ) plot of observed versus expected p-value distributions, with the lambda inflation statistic reported above.

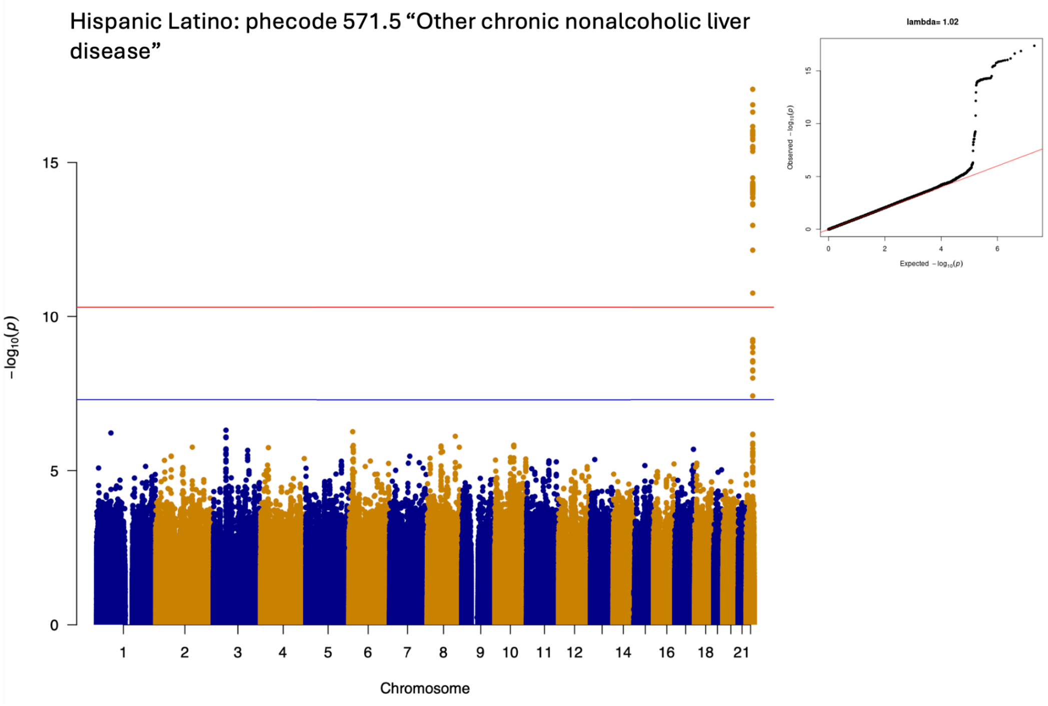

**Supplemental Figure 29**: Manhattan plot displaying GWAS association signal for phecode 571.5 "Other chronic nonalcoholic liver disease" in Bio*Me* HL. Each point represents -log10 p-value at each tested variant colored by chromosome. The blue horizontal line indicates genome wide significance, while the red horizontal line indicates study wide significance. An inset shows a quantile-quantile (QQ) plot of observed versus expected p-value distributions, with the lambda inflation statistic reported above.

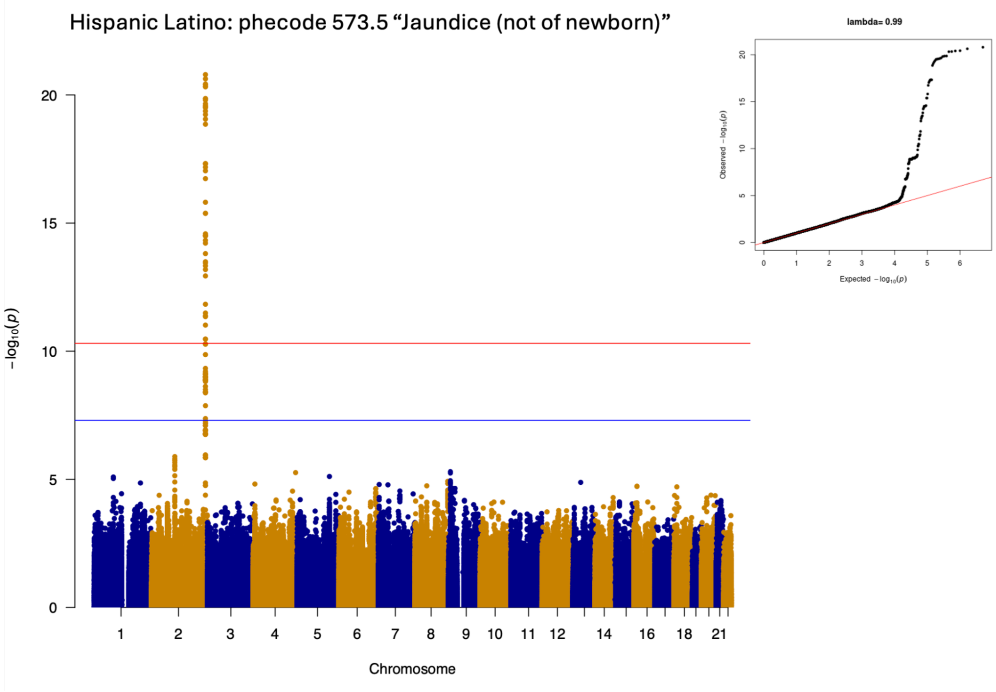

**Supplemental Figure 30**: Manhattan plot displaying GWAS association signal for phecode 573.5 "Jaundice (not of newborn)" in Bio*Me* HL. Each point represents -log10 p-value at each tested variant colored by chromosome. The blue horizontal line indicates genome wide significance, while the red horizontal line indicates study wide significance. An inset shows a quantile-quantile (QQ) plot of observed versus expected p-value distributions, with the lambda inflation statistic reported above.

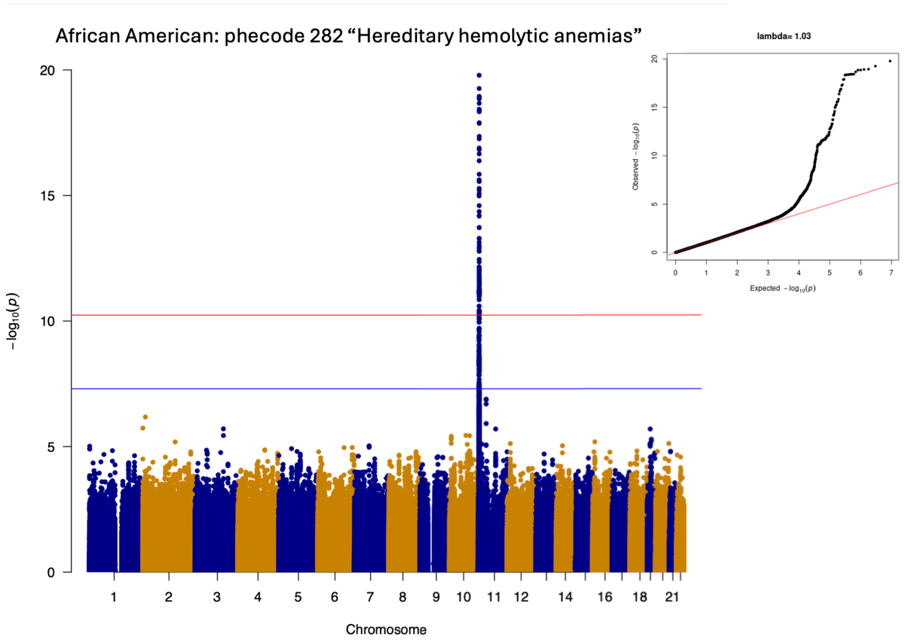

**Supplemental Figure 31**: Manhattan plot displaying GWAS association signal for phecode 282 "Hereditary hemolytic anaemias" in Bio*Me* AA. Each point represents -log10 p-value at each tested variant colored by chromosome. The blue horizontal line indicates genome wide significance, while the red horizontal line indicates study wide significance. An inset shows a quantile-quantile (QQ) plot of observed versus expected p-value distributions, with the lambda inflation statistic reported above.

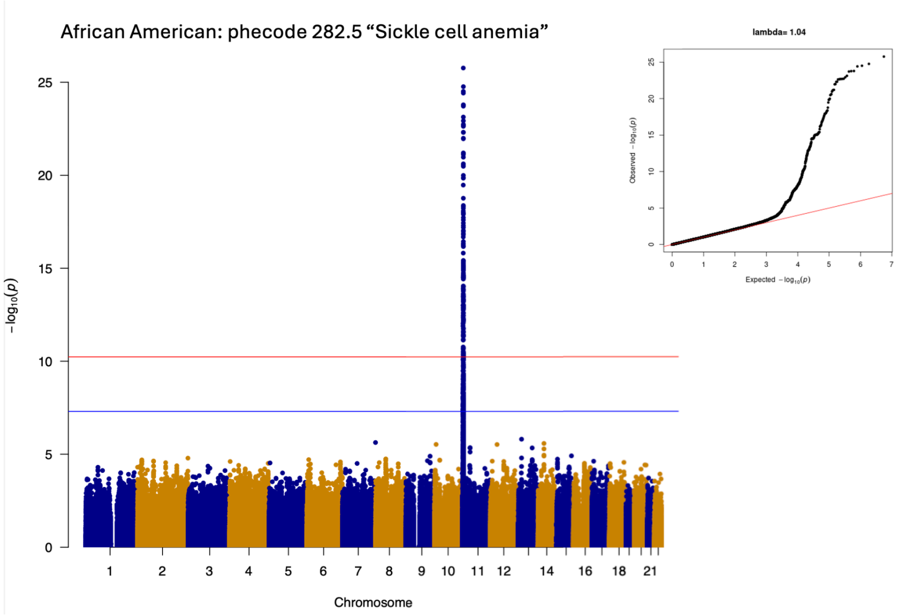

**Supplemental Figure 32**: Manhattan plot displaying GWAS association signal for phecode 282.5 "Sickle cell anemia" in BioMe AA. Each point represents -log10 p-value at each tested variant colored by chromosome. The blue horizontal line indicates genome wide significance, while the red horizontal line indicates study wide significance. An inset shows a quantile-quantile (QQ) plot of observed versus expected p-value distributions, with the lambda inflation statistic reported above.

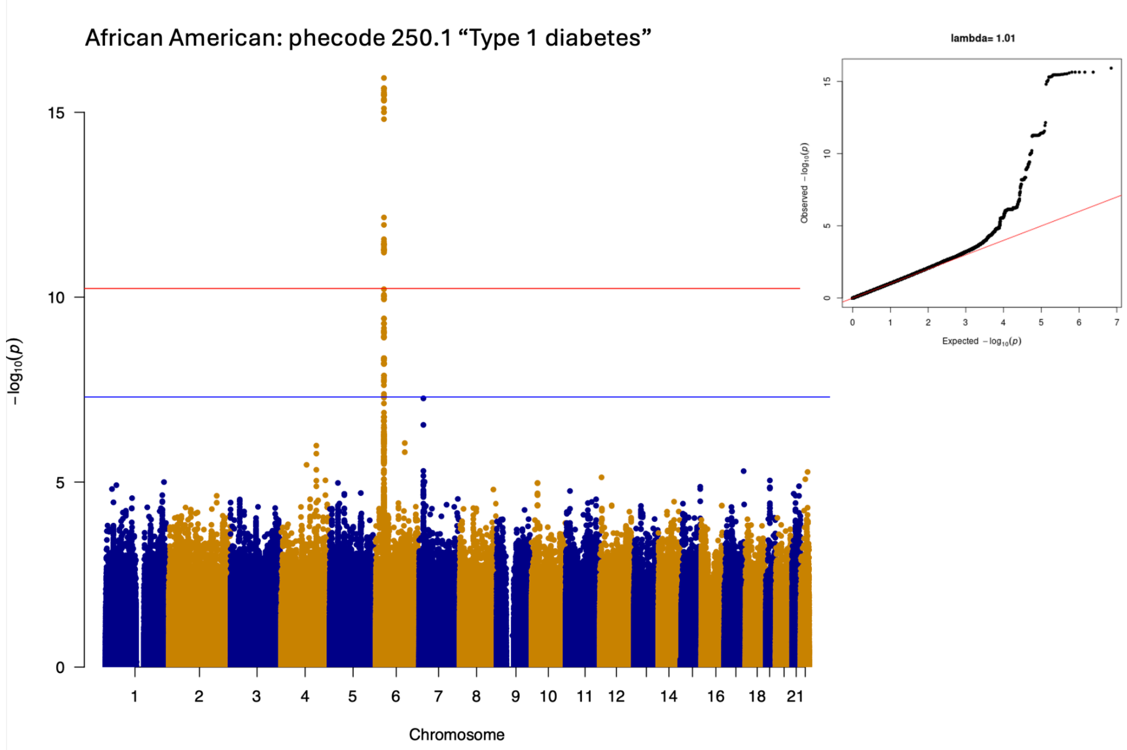

**Supplemental Figure 33**: Manhattan plot displaying GWAS association signal for phecode 250.1 "Type 1 Diabetes" in Bio*Me* AA. Each point represents -log10 p-value at each tested variant colored by chromosome. The blue horizontal line indicates genome wide significance, while the red horizontal line indicates study wide significance. An inset shows a quantile-quantile (QQ) plot of observed versus expected p-value distributions, with the lambda inflation statistic reported above.

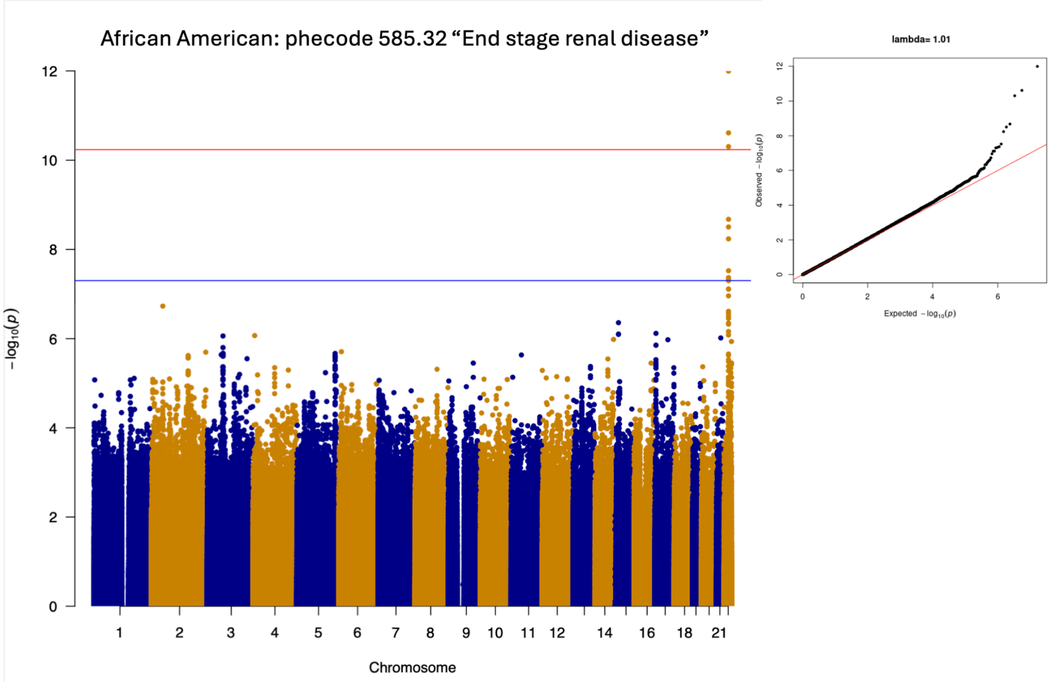

**Supplemental Figure 34**: Manhattan plot displaying GWAS association signal for phecode 585.32 " End Stage Renal Disease" in Bio*Me* African Americans. Each point represents -log10 p-value at each tested variant colored by chromosome. The blue horizontal line indicates genome wide significance, while the red horizontal line indicates study wide significance. An inset shows a quantile-quantile (QQ) plot of observed versus expected p-value distributions, with the lambda inflation statistic reported above.

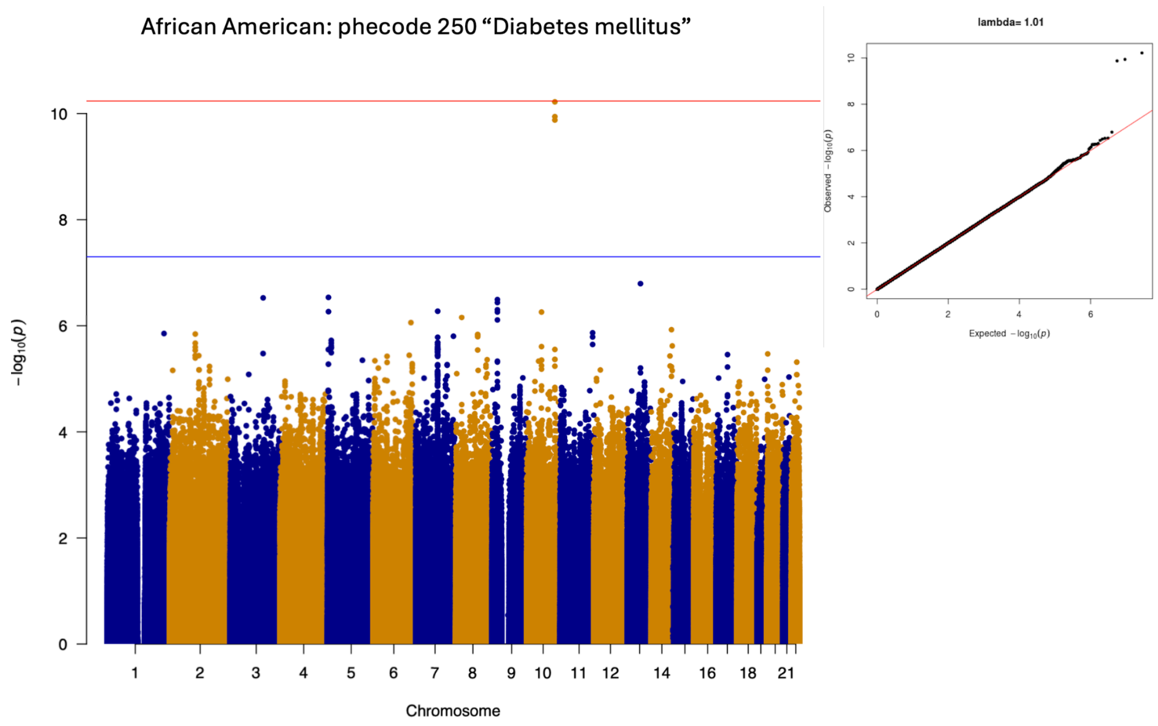

**Supplemental Figure 35**: Manhattan plot displaying GWAS association signal for phecode 250, " Diabetes Mellitus" in Bio*Me* AA. Each point represents -log10 p-value at each tested variant colored by chromosome. The blue horizontal line indicates genome wide significance, while the red horizontal line indicates study wide significance. An inset shows a quantile-quantile (QQ) plot of observed versus expected p-value distributions, with the lambda inflation statistic reported above.

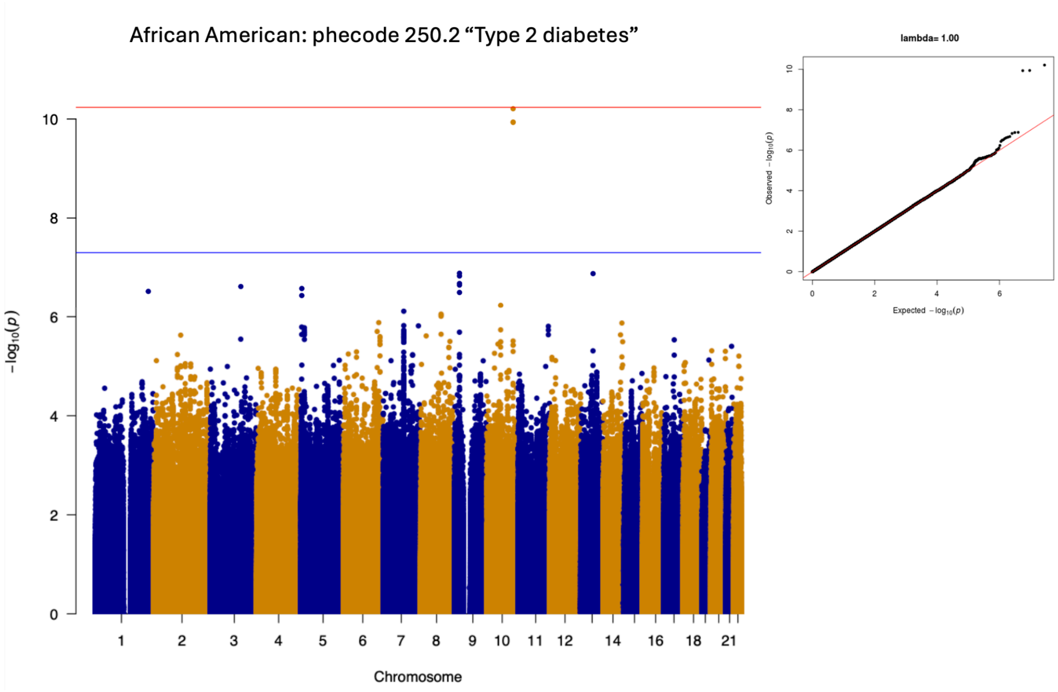

**Supplemental Figure 36**: Manhattan plot displaying GWAS association signal for phecode 250.2, "Type 2 diabetes" in Bio*Me* AA. Each point represents -log10 p-value at each tested variant colored by chromosome. The blue horizontal line indicates genome wide significance, while the red horizontal line indicates study wide significance. An inset shows a quantile-quantile (QQ) plot of observed versus expected p-value distributions, with the lambda inflation statistic reported above.

| Test Cohort | % power | RR | RR power | AER | Sample Size |
|---|---|---|---|---|---|
| Bio*Me* HL | 84.2% (16/19) | 57.9% (11/19) | 69% (11/16) | 0.64 | 14,878 |
| Bio*Me* AA | 31.8% (7/22) | 59.0% (12/22) | 43% (3/7) | 0.88 | 8877 |

**Supplemental Table 5:** Phenotype-genotype reference map (PGRM) replication summary statistics in Bio*Me*. RR = overall replication rate , RR power = powered replication rate, AER= actual:expected ratio, For further details on replication definitions, refer to (Bastarache et al. 2023)

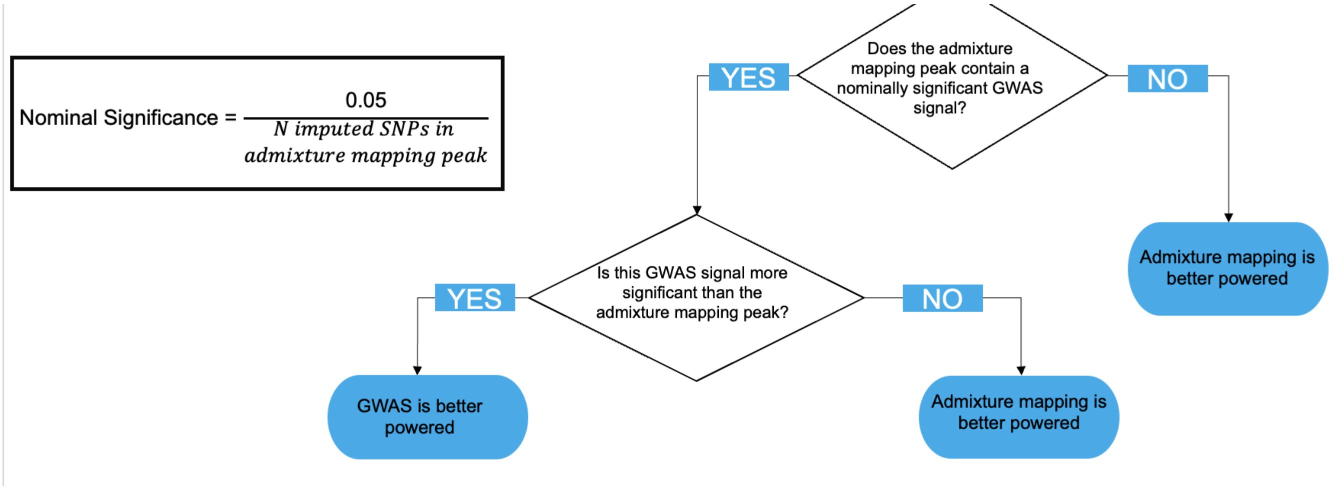

**Supplemental Figure 37:** Decision tree used to define GWAS association signal for variants falling within genome wide significant admixture mapping peaks, for main text Figure 2A.

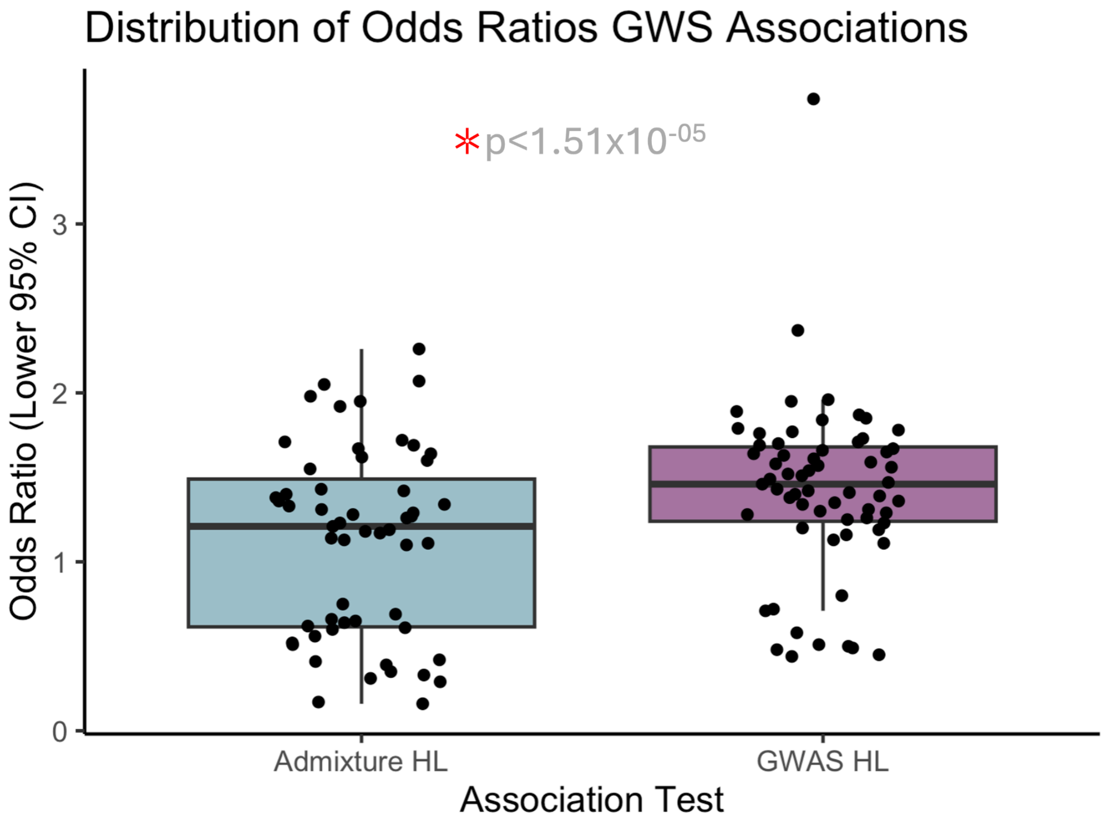

**Supplemental Figure 38:** Boxplot showing the distribution of the lower 95% confidence interval of odds ratios in admixture mapping vs GWAS analyses.
